# Use of Internally Validated Machine and Deep Learning Models to Predict Outcomes of Percutaneous Nephrolithotomy using data from the BAUS PCNL audit

**DOI:** 10.1101/2022.06.16.22276481

**Authors:** Robert M Geraghty, William Finch, Sarah Fowler, Seshadri Sriprasad, Daron Smith, Andrew Dickinson, Zara Gall, Bhaskar K Somani

## Abstract

**Background:** Machine (ML) and Deep learning (DL) are subsets of artificial intelligence that use data to build algorithms. These can be used to predict specific outcomes. To date there have been a few small studies on post-PCNL outcomes.

**Objective:** We aimed to build and internally validate ML/DL models for post-PCNL transfusion and infection using a comprehensive national database.

**Design:** Machine Learning study using prospective national database. Eight machine learning models for 11 outcomes using 43 predictors. Models were ‘complete-case’ analyses.

**Setting:** National database

**Participants:** Patients undergoing PCNL in the UK between 2014-2019.

**Outcome Measurements:** Diagnostic accuracy statistics including overall accuracy, area-under-the-curve (AUC), sensitivity and specificity.

**Results and Limitations:** 4412 patients were included, with 3088 in the training set and 1324 in the test set. The models predicted need for transfusion and post-operative infection with a very high degree of accuracy (99%) and high AUC (0.99-1.00). Unfortunately, the remainder of the outcomes did not achieve the same high levels. These two outcomes were therefore included in the provisional web-based application: https://endourology.shinyapps.io/PCNL_Prediction_tool/

**Conclusions:** This is the largest machine learning study on post-PCNL outcomes to date. These models can predict the need for post-PCNL transfusion and post-PCNL infection at an individual level with excellent accuracy. Further work will be done on model tuning and external validation.

**Patient Summary:** We used a national database of people having a major kidney stone operation (PCNL). Using this data, we built and tested 8 machine learning models for 11 different outcomes from the operation. Using this method, we can give individual predictions for the likely need for a blood transfusion and development of an infection. We have developed an app to allow surgeons to calculate an individual patient’s risk prior to surgery.

## 1. Introduction

Kidney stone disease is a highly prevalent and costly disease[1]. Large kidney stones are often treated by percutaneous nephrolithotomy (PCNL)[2]. In addition to the planned outcome of removing the stone (stone free status), PCNL has a number of known complications including need for transfusion, post-operative infection and visceral injury[3]. Several scoring systems have been built to attempt to predict outcomes for individual patients both in terms of stone free status and complications[4]. More recently, (supervised) machine learning (ML) techniques have been used to attempt to predict outcomes of PCNL[5–7]. Notably, Aminsharifi et al. compared a ML model (support vector machines) to the more traditional nomogram based systems (Guy’s Stone score and the CROES PCNL nomogram)[6], and demonstrated far superior accuracy of ML. To date, there are only four outcomes for which predictive models are described: stone free status, need for adjuvant treatment, need for stent insertion and need for blood transfusion.

We therefore aimed to utilize a large national database to develop ML and more modern deep learning (DL) models to predict a larger number of outcomes (n=11), with subsequent internal validation and model implementation in a web-based application for easily accessible individualized predictions.

## 2. Methods

### 2.1 Methodology reporting

We report this study using the TRIPOD checklist [8] (see supplementary material).

### 2.2 Patients and Dataset

We utilized data from the BAUS PCNL audit, the methods of data collection have previously been reported[9], but we briefly report them here: Through advertisement at national urologic meetings, all surgeons undertaking PCNL in the United Kingdom were invited to submit data to the registry using an online interface. An individual record that contained both a unique patient identifier and National Health Service(NHS) number was created for each PCNL procedure. Data was collected between 2014-2019.

### 2.3 Predictors and Outcomes

#### 43 predictors taken at operation

Age, BMI, Pre-operative haemoglobin (g/L), Charlson score (0-10), Age-related Charlson score (0-11) [10], number of tracts planned, number of tracts performed, sex, side of stones, previous UTI treatment, pre-operative antibiotic course, pre-operative urine culture, pre-operative urine culture result, primary pre-operative imaging, secondary pre-operative imaging, pre-operative dimercapto-succinic acid (DMSA) renogram, catheterization status, pre-operative estimated glomerular filtration rate (eGFR), prophylactic antibiotics on induction, grade of main operating surgeon, type of anaesthesia, interventional radiologist availability, secondary re-look nephroscopy, stone dimensions (cm), number of stones, index stone location, other stone location(s), Guy’s stone score[11], maximum Hounsfield units of index stone on CT KUB, pre-existing nephrostomy tube status, specialty and grade of practitioner performing puncture tract, puncture site, image guidance for renal puncture, patient position, anatomical placement of tract, size of amplatz sheath (Fr), type of dilators used, predicted difficulty, accessory procedures, post-operative nephrostomy, primary and secondary stone extraction techniques.

#### 11 outcomes

immediate clearance on fluoroscopy, visceral injury, survival, need for transfusion, post-operative infection, intra-operative complication, need for higher care (high dependency unit or intensive care unit), stone free at follow-up (first outpatient review using radiography,ultrasonography, or computed tomography according to local practice), need for adjuvant treatment, post-operative stay duration (0, 1, 2, ≥3 days) and Clavien-Dindo classification of complication [12].

### 2.4 Sample Size Calculation

Sample size was calculated for the least likely event i.e largest number of patients needed (vascular injury necessitating nephrectomy ∼0.1%). Sample size was calculated using a 0.1% population proportion (margin of error ±0.1%), to be n=3838. This would give a likely representative sample adequate for ML/DL.

### 2.5 Missing Data

Given the known issues around the inaccuracy of imputed data [13], and therefore the likely subsequent inaccuracy of models built on this data, patients with missing data were excluded. This study is therefore a ‘complete-case’ analysis.

### 2.6 Model Selection

We constructed seven different single-outcome classification models: logistic regression (traditional statistical technique), five classical machine learning (ML) models chosen to be representative of differing ML techniques (random forests, extreme gradient boosting [xgboost], Bayesian generalized linear model, partitioning and neural networks), along with more novel deep learning (DL) neural networks using ‘keras’ with ‘tensorflow’. Using DL neural networks, we also built a multiple-outcome classification model to predict all 11 outcomes.

### 2.7 Model Building

All models were built in R (version 4.1.2, Vienna, Austria)[14] using the ‘caret’[15], ‘keras’[16] and ‘tensorflow’[17] packages. Model calibration is performed automatically by ‘caret’ for the ML models. The DL neural networks were tested/calibrated with two and three layer nets, along with three different node layer sizes (48, 70 and 112). Full code of final models is available as supplementary material.

### 2.8 Internal Validation

Datasets were randomly split into training (70% of total) and test (30%) sets. The test set was used to internally validate the models for each outcome. We report total, training and test set demographics.

### 2.9 Statistical Analysis

Summary statistics are provided for example training and test sets. Each factor is compared between training and test sets to confirm randomisation as follows: categorical - chi^2^ tests, or Fisher’s exact test (n<5), normally distributed continuous - independent T-Tests and non-normally distributed continuous - Mann Whitney U tests.

We report diagnostic accuracy statistics following internal validation for each model: overall accuracy with 95% confidence interval, sensitivity, specificity, and area under the curve (AUC-ROC). Negative predictive values (NPV) and positive predictive values (PPV) are available as supplementary material. We present receiver operator curves (ROC) for all models. These were generated using the ‘MLeval’[18], ‘caret’[15], ‘pROC’ and ‘ggplot2’[19] packages.

### 2.10 Model Selection and Deployment

Models that were highly accurate, sensitive and specific with an AUC ≥0.8 were deployed online via the ‘shiny’ package within R [20]. Model explainers using the ‘lime’ (Local Interpretable Model-Agnostic Explainer) package are utilized to give explanations as to why the model is predicting a particular outcome [21]. Code is available as supplementary material.

## 3. Results

### 3.1 Demographics

In the total dataset (n=4418), the mean age was 56.5(±19.4), with 2074 women (46%), median BMI was 28.4 (IQR:25.0-33.0), median Charlson score was 1 (IQR:1-2), n=2133 (48%) had a previous UTI, n=1214 (48%) had some form of antibiotic cover prior to the procedure, n=3993 (90%) had a urine culture sent prior to the procedure. For further demographics of total, training and test sets please see supplementary table 1. There were no significant differences between training and tests sets on individual variable comparison, thus demonstrating randomization has worked.

In the total dataset the outcomes were as follows [see figure 1]: clearance on immediate post-operative imaging n=2144 (49%), visceral injury n=13 (0.3%), death n=7 (0.2%), post-operative transfusion n=196 (4%), post-operative infection n=966 (22%), intra-operative complication(s) n=140 (3%, n=360 with missing outcome), need for ITU/HDU admission n=201 (5%, n=218 with missing outcome), stone free at follow-up n=763 (70%, n=3327 with missing outcome) and need for adjuvant treatment n=185 (17%, n=3316 with missing outcome). Post-operative stay was subdivided into: daycase (n=52, 1%), 1 day (n=886, 20%), 2 days (n=998, 26%) and ≥3 days (n=2482, 56%). Clavien-Dindo classification of complications were as follows: I n=311 (7%), II n=343 (8%), IIIa n=93 (2%), IIIb n=63 (1%), IVa n=13 (0.3%), IVb n=0 and V n=7 (0.2%). Details of outcomes in training/test sets are detailed below.

**Figure 1.**
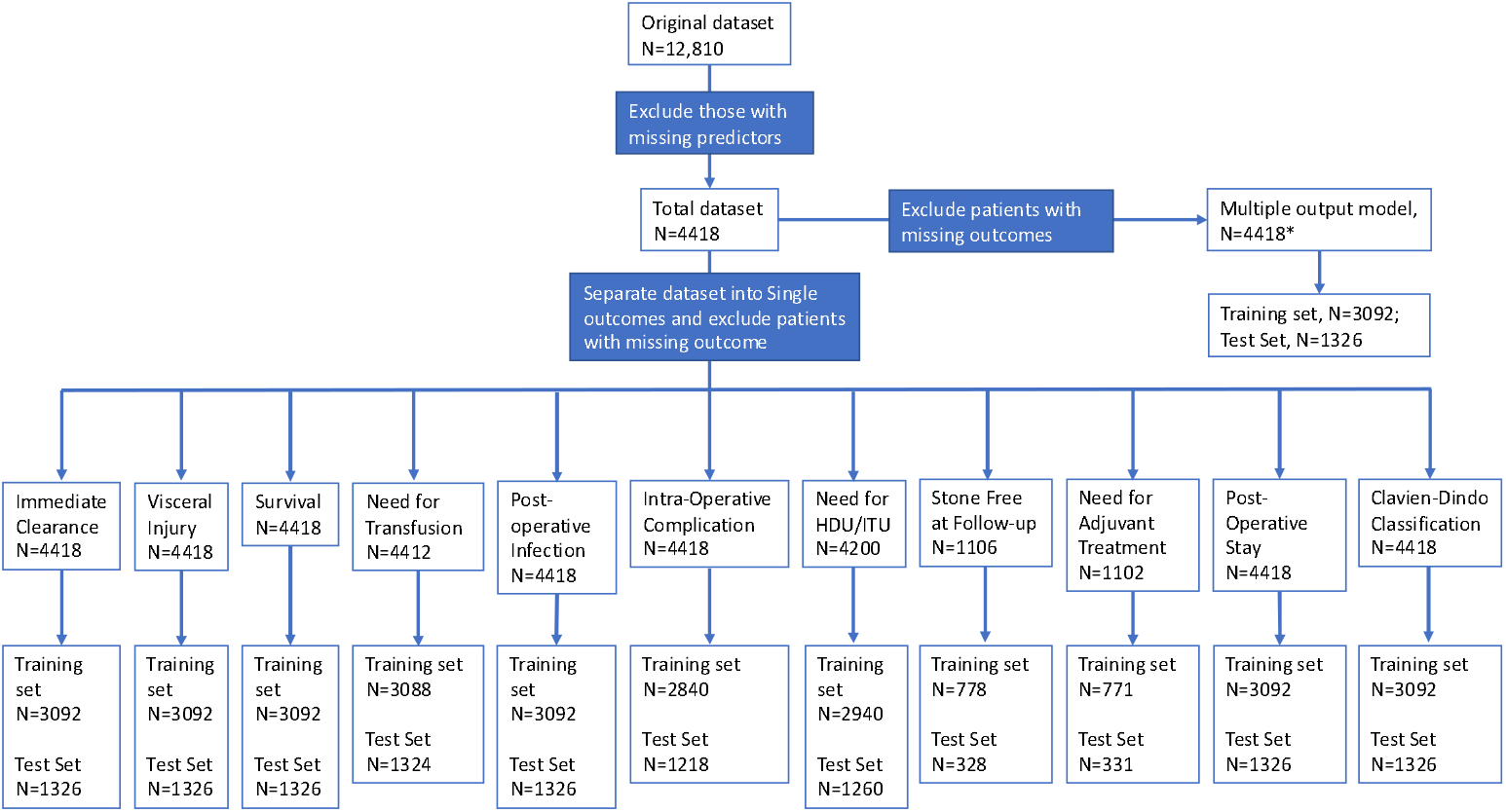
Flow Diagram of patient selection. The ‘N’ refers to total number of patients in dataset at particular stage of flow diagram, it does not relate to number with specific outcome. Outcomes split by 70% (training) and 30% (testing). *=Larger N for multiple output model despite small numbers of total patients with data on ‘Stone Free’ and ‘Need for Adjuvant Treatment’ as missing data were coded as a ‘0’ i.e ‘no’.

### 3.2 Models for Specific Outcomes

#### 3.2.1 Immediate clearance

The training group had n=3092 patients, n=2360 of which had immediate clearance on fluoroscopy. On internal validation (test set: n with outcome/total; n=1015/1326), the diagnostic accuracy statistics were: Random forests (RF) AUC=0.73, accuracy= 0.77 (95% CI:0.74-0.79), sensitivity=0.27, specificity=0.94; Partitioning AUC=0.63, accuracy=0.71 (95% CI:0.69-0.74), sensitivity=0.22, specificity=0.89; Extreme Gradient Boosting (XGBoost) AUC=0.75, accuracy=0.77 (95% CI:0.75-0.80), sensitivity=0.27, specificity=0.94; Logistic Regression (LR) AUC=0.63, accuracy=0.76 (95% CI:0.73-0.78), sensitivity=0.12, specificity=0.97; Classical Neural Network (NN) AUC=0.74, accuracy=0.75 (95% CI:0.73-0.78), sensitivity=0.23, specificity=0.94; Bayesian Generalised Linear Model (BGLM) AUC=0.75, accuracy=0.77 (95% CI:0.75-0.80), sensitivity=0.29, specificity=0.94; Deep Neural Network (DNN) Single-outcome model AUC=0.59, accuracy=0.77 (95% CI:0.75-0.79), sensitivity=0.57, specificity=0.79; DNN Multiple-outcome model AUC=0.53, accuracy=0.77 (95% CI:0.75-0.79), sensitivity=0.60, specificity=0.78 [see figure 2 and supplementary table **2**].

**Figure 2.**
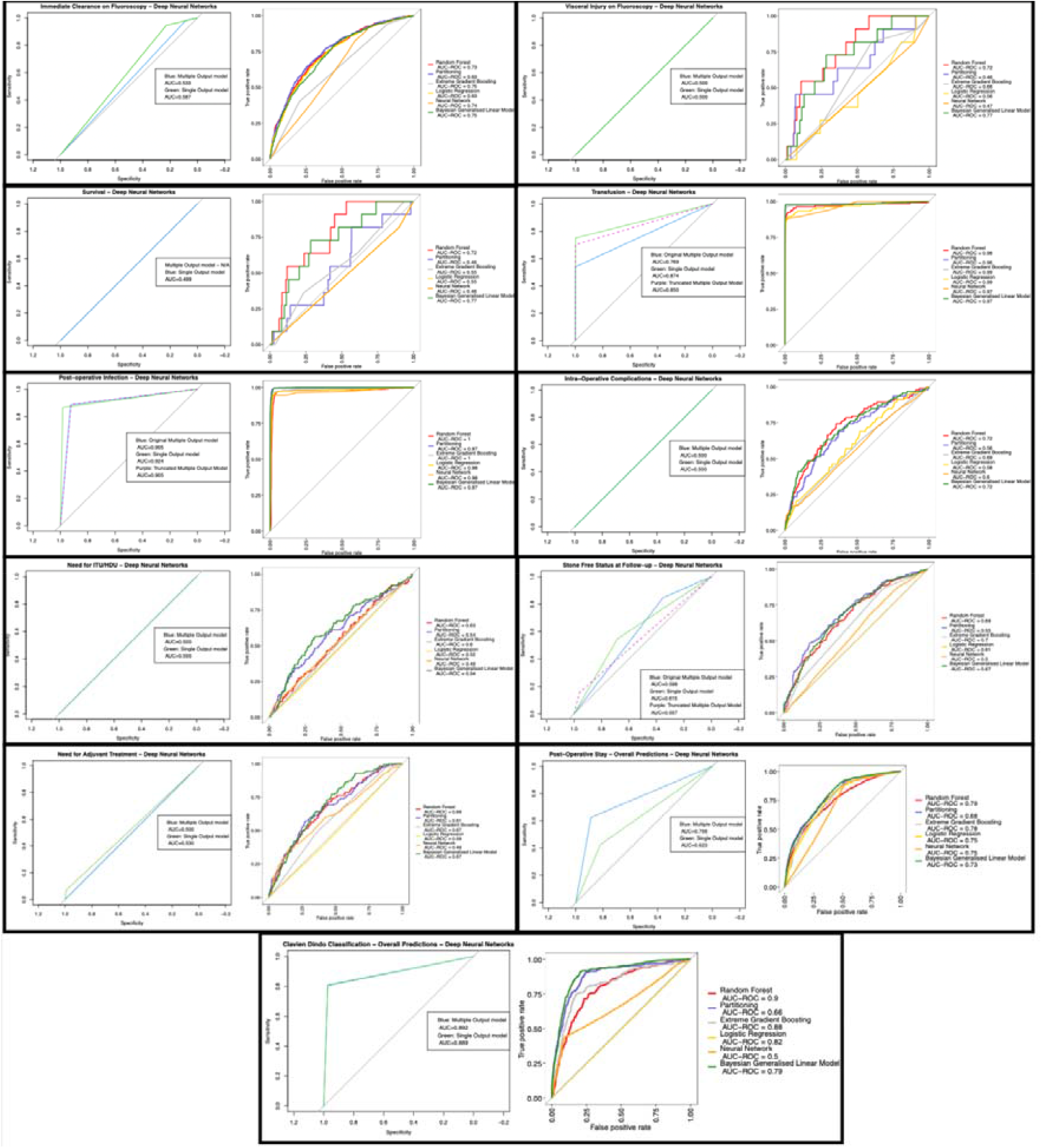
ROC curves for each model

#### 3.2.2 Visceral Injury

The training group (n=3092 patients) had n=11 with visceral injury. On internal validation (test set: n with outcome/total; n=5/1326), the diagnostic accuracy statistics were: RF AUC=0.72, accuracy=0.99 (95% CI:0.99=1.00), sensitivity=1.00, specificity=0.00; Partitioning AUC=0.46, accuracy=0.99 (95% CI:0.99=1.00), sensitivity=1.00, specificity=0.00; XGBoost AUC=0.66, accuracy=0.99 (95% CI:0.99=1.00), sensitivity=1.00, specificity=0.00; LR AUC=0.56, accuracy=0.99 (95% CI:0.99=1.00), sensitivity=1.00, specificity=0.00; NN AUC=0.47, accuracy=0.99 (95% CI:0.99=1.00), sensitivity=1.00, specificity=0.00; BGLM AUC=0.77, accuracy=0.99 (95% CI:0.99=1.00), sensitivity=1.00, specificity=0.00 ; DNN Single-outcome model AUC=0.50, accuracy=0.99 (95% CI:0.99-1.00), sensitivity=0.99, specificity=N/A; DNN Multiple-outcome model AUC=0.50, accuracy=0.99 (95% CI:0.99-1.00), sensitivity=0.99, specificity=N/A [see supplementary table **3**].

#### 3.2.3 Survival

The training group (n=3092 patients) had n=11 who died. On internal validation (test set: n with outcome/total; n=1/1326), the diagnostic accuracy statistics were: RF AUC=0.72, accuracy=0.99 (95% CI:0.99-1.00), sensitivity=1.00, specificity=0.00; Partitioning AUC=0.46, accuracy=0.99 (95% CI:0.99-1.00), sensitivity=1.00, specificity=0.00; XGBoost AUC=0.55, accuracy=0.99 (95% CI:0.99-1.00), sensitivity=1.00, specificity=0.00; LR AUC=0.55, accuracy=0.99 (95% CI:0.99-1.00), sensitivity=1.00, specificity=0.00; NN AUC=0.46, accuracy=0.99 (95% CI:0.99-1.00), sensitivity=1.00, specificity=0.00; BGLM AUC=0.77, accuracy=0.99 (95% CI:0.99-1.00), sensitivity=1.00, specificity=0.00; DNN Single-outcome model AUC=0.55, accuracy=0.99 (95% CI:0.99-1.00), sensitivity=1.00, specificity=0.00. Due to the poor predictive value as above, Survival was not included in the multi-output model [see supplementary table **4**].

#### 3.2.4 Need for Transfusion

The training group (n=3088 patients) had n=132 who were transfused. On internal validation (test set: n with outcome/total; n=63/1324), the diagnostic accuracy statistics were: RF AUC=0.98, accuracy=0.99 (95% CI:0.99-1.00), sensitivity=1.00, specificity=0.97; Partitioning AUC=0.96, accuracy=0.99 (95% CI:0.99-1.00), sensitivity=1.00, specificity=0.89; XGBoost AUC=0.99, accuracy=1.00 (95% CI:0.99-1.00), sensitivity=1.00, specificity=0.97; LR AUC=0.99, accuracy=1.00 (95% CI:0.99-1.00), sensitivity=1.00, specificity=0.92 ; NN AUC=0.97, accuracy=0.99 (95% CI:0.98-0.99), sensitivity=1.00, specificity=0.80 ; BGLM AUC=0.97, accuracy=0.99 (95% CI:0.99-1.00), sensitivity=1.00, specificity=0.84 ; DNN Single-outcome model AUC=0.87, accuracy=0.99 (95% CI:0.98-0.99), sensitivity=0.99, specificity=0.96 ; DNN Multiple-outcome model AUC=0.77, accuracy=0.98 (95% CI:0.97-0.99), sensitivity=0.98, specificity=0.96 [see table **1**].

**Table 1.**
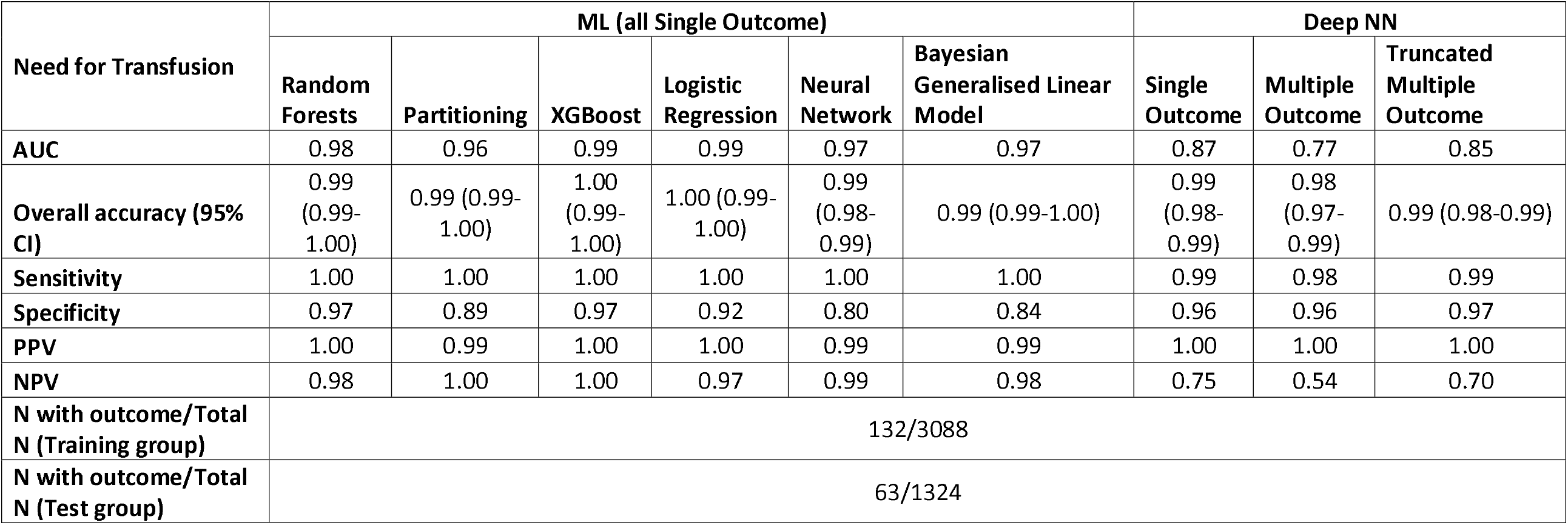
Diagnostic accuracy statistics for ‘Need for Transfusion’. PPV=positive predictive value, NPV=negative predictive value, XGBoost=Extreme Gradient Boosting, NN=Neural network. Truncated model included: Stone free status, Need for transfusion and post-operative infection

#### 3.2.5 Post-operative infection

The training group (n=3092 patients) had n=684 who developed a post-operative infection. On internal validation (test set: n with outcome/total; n=285/1326), the diagnostic accuracy statistics were: RF AUC=1.00, accuracy=0.98 (95% CI:0.97-0.99), sensitivity=0.99, specificity=0.96; Partitioning AUC=0.97, accuracy=0.97 (95% CI:0.96-0.98), sensitivity= 0.99, specificity=0.93; XGBoost AUC=1.00, accuracy=0.98 (95% CI:0.98-0.99), sensitivity=0.99, specificity=0.97 ; LR AUC=0.98, accuracy=0.98 (95% CI:0.97-0.98), sensitivity=0.98, specificity=0.95; NN AUC=0.98, accuracy=0.98 (95% CI:0.97-0.98), sensitivity=0.98, specificity=0.94; BGLM AUC=0.97, accuracy=0.95 (95% CI:0.94-0.97), sensitivity=0.98, specificity=0.86; DNN Single outcome model AUC=0.92, accuracy=0.96 (95% CI:0.95-0.97), sensitivity=0.96, specificity=0.93; DNN Multiple-outcome model AUC=0.90, accuracy=0.92 (95% CI:0.90-0.93), sensitivity=0.96, specificity=0.78 [see table **2**].

**Table 2.** Diagnostic accuracy statistics for Post-operative Infection. PPV=positive predictive value, NPV=negative predictive value, XGBoost=Extreme Gradient Boosting, NN=Neural network. Truncated model included: Stone free status, Need for transfusion and post-operative infection

| Post-Operative Infection | ML (all Single Outcome) |  |  |  |  |  | Deep NN |  |  |
| --- | --- | --- | --- | --- | --- | --- | --- | --- | --- |
|  | Random Forests | Partitioning | XGBoost | Logistic Regression | Neural Network | Bayesian Generalised Linear Model | Single Outcome | Multiple Outcome | Truncated Multiple Outcome |
| **AUC** | 1.00 | 0.97 | 1.00 | 0.98 | 0.98 | 0.97 | 0.92 | 0.90 | 0.90 |
| **Overall accuracy (95% CI)** | 0.98<br>(0.97-0.99) | 0.97 (0.96-0.98) | 0.98<br>(0.97-0.99) | 0.98 (0.97-0.98) | 0.98<br>(0.97-0.98) | 0.95 (0.94-0.97) | 0.96<br>(0.95-0.97) | 0.92<br>(0.90-0.93) | 0.91 (0.90-0.93) |
| **Sensitivity** | 0.99 | 0.99 | 0.99 | 0.98 | 0.98 | 0.98 | 0.96 | 0.96 | 0.96 |
| **Specificity** | 0.96 | 0.93 | 0.97 | 0.95 | 0.94 | 0.86 | 0.93 | 0.78 | 0.78 |
| **PPV** | 0.99 | 0.98 | 0.99 | 0.99 | 0.98 | 0.96 | 0.98 | 0.93 | 0.92 |
| **NPV** | 0.95 | 0.95 | 0.95 | 0.94 | 0.94 | 0.92 | 0.87 | 0.88 | 0.89 |
| **N with outcome/Total N (Training group)** | 684/3092 |  |  |  |  |  |  |  |  |
| **N with outcome/Total N (Test group)** | 285/1326 |  |  |  |  |  |  |  |  |

#### 3.2.6 Intra-operative complication

The training group (n=2840 patients) had n=93 who had an intra-operative complication. On internal validation (test set: n with outcome/total; n=47/1218), the diagnostic accuracy statistics were: RF AUC=0.72, accuracy=0.96 (95% CI:0.95-0.97), sensitivity=1.00, specificity=0.00; Partitioning AUC=0.56, accuracy=0.96 (95% CI:0.95-0.97), sensitivity=1.00, specificity=0.00; XGBoost AUC=0.69, accuracy=0.96 (95% CI:0.96-0.95-0.97), sensitivity=1.00, specificity=0.00; LR AUC=0.58, accuracy=0.96 (95% CI:0.95-0.97), sensitivity=1.00, specificity=0.04; NN AUC=0.60, accuracy=0.96 (95% CI:0.95-0.97), sensitivity=1.00, specificity=0.00; BGLM AUC=0.72, accuracy=0.96 (95% CI:0.95-0.97), sensitivity=1.00, specificity=0.02; DNN Single-outcome model AUC=0.50, accuracy=0.96 (95% CI:0.96-0.97), sensitivity=0.96, specificity=N/A; DNN Multiple-outcome model AUC=0.50, accuracy=0.97 (95% CI:0.95-0.97), sensitivity=0.97, specificity=N/A [see supplementary table **5**].

#### 3.2.7 Need for HDU/ITU

The training group (n=2940 patients) had n=128 who required higher care. On internal validation (test set: n with outcome/total; n=73/1260), the diagnostic accuracy statistics were: RF AUC=0.63, accuracy=0.94 (95% CI:0.93-0.95), sensitivity=1.00, specificity=0.00; Partitioning AUC=0.54, accuracy=0.94 (95% CI:0.93-0.95), sensitivity=1.00, specificity=0.00; XGBoost AUC=0.60, accuracy=0.94 (95% CI:0.93-0.95), sensitivity=1.00, specificity=0.00; LR AUC=0.52, accuracy=0.94 (95% CI:0.93-0.95), sensitivity=1.00, specificity=0.00; NN AUC=0.49, accuracy=0.94 (95% CI:0.93-0.95), sensitivity=1.00, specificity=0.00; BGLM AUC=0.54, accuracy=0.94 (95% CI:0.93-0.95), sensitivity=1.00, specificity=0.00; DNN Single-outcome model AUC=0.50, accuracy=0.94 (95% CI:0.93-0.95), sensitivity=0.94, specificity=N/A; DNN Multiple-outcome model AUC=0.50, accuracy=0.95 (95% CI:0.94-0.96), sensitivity=0.95, specificity=N/A [see supplementary table **6**].

#### 3.2.8 Stone Free at Follow-up

The training group (n=778 patients) had n=535 who were stone free at follow-up. On internal validation (test set: n with outcome/total; n=228/328), the diagnostic accuracy statistics were: RF AUC=0.69, accuracy=0.70 (95% CI:0.64-0.0.74), sensitivity=0.00, specificity=1.00; Partitioning AUC=0.55, accuracy=0.70 (95% CI:0.64-0.74), sensitivity=0.00, specificity=1.00; XGBoost AUC=0.70, accuracy=0.65 (95% CI:0.60-0.70), sensitivity=0.20, specificity=0.87; LR AUC=0.61, accuracy=0.62 (95% CI:0.56-0.67), sensitivity=0.30, specificity=0.78; NN AUC=0.50, accuracy=0.70 (95% CI:0.64-0.74), sensitivity=0.00, specificity=1.00; BGLM AUC=0.67, accuracy=0.69 (95% CI:0.63-0.74), sensitivity=0.30, specificity=0.88; DNN Single-outcome model AUC=0.62, accuracy=0.59 (95% CI:0.54-0.65), sensitivity=0.43, specificity=0.78; DNN Multiple-outcome model AUC=0.60, accuracy=0.44 (95% CI:0.41-0.46), sensitivity=0.92, specificity=0.21 [see supplementary table **7**].

#### 3.2.9 Need for Adjuvant Treatment

The training group (n=771 patients) had n=126 who needed adjuvant treatment. On internal validation (test set: n with outcome/total; n=59/331), the diagnostic accuracy statistics were: RF AUC=0.69, accuracy=0.82 (95% CI:0.77-0.86), sensitivity=0.99, specificity=0.02; Partitioning AUC=0.61, accuracy=0.83 (95% CI:0.78-0.87), sensitivity=0.98, specificity=0.12; XGBoost AUC=0.67, accuracy=0.82 (95% CI:0.78-0.86), sensitivity=1.00, specificity=0.02; LR AUC=0.59, accuracy=0.79 (95% CI:0.75-0.84), sensitivity=0.94, specificity=0.14; NN AUC=0.49, accuracy=0.82 (95% CI:0.78-0.86), sensitivity=1.00, specificity=0.00; BGLM AUC=0.67, accuracy=0.83 (95% CI:0.78-0.87), sensitivity=0.98, specificity=0.14; DNN Single-outcome model AUC=0.53, accuracy=0.83 (95% CI:0.78-0.87), sensitivity=0.83, specificity=0.67; DNN Multiple-outcome model AUC=0.50, accuracy=0.96 (95% CI:0.95-0.97), sensitivity=0.96, specificity=N/A [see supplementary table **8**].

#### 3.2.10 Post-operative Stay

On internal validation (test set, n=1326), the overall diagnostic accuracy statistics were: RF AUC=0.79, accuracy=0.66 (95% CI:0.63-0.69); Partitioning AUC=0.68, accuracy=0.66 (95% CI:0.63-0.68); XGBoost AUC=0.78, accuracy=0.68 (95% CI:0.65-0.70); LR AUC=0.75, accuracy=0.67 (95% CI:0.64-0.69); NN AUC=0.75, accuracy=0.68 (95% CI:0.65-0.70); BGLM AUC=0.73, accuracy=0.56 (95% CI:0.53-0.59); DNN Single-outcome model AUC=0.62, accuracy=0.72 (95% CI:0.71-0.73); DNN Multiple-outcome model AUC=0.76, accuracy=0.82 (95% CI:0.81-0.83).

Sensitivity, specificity, PPV and NPV for each length of stay (0,1,2 and ≥3 days) are detailed in supplementary table **9**.

#### 3.2.11 Clavien Dindo Classification

On internal validation (test set, n=1326), the diagnostic accuracy statistics were: RF AUC=0.90, accuracy=0.85 (95% CI:0.83-0.87); Partitioning AUC=0.66, accuracy=0.83 (95% CI:0.81-0.85); XGBoost AUC=0.88, accuracy=0.85 (95% CI:0.83-0.87); LR AUC=0.82, accuracy=0.86 (95% CI:0.84-0.88); NN AUC=0.50, accuracy=0.82 (95% CI:0.80-0.84); BGLM AUC=0.79, accuracy=0.82 (95% CI:0.80-0.84); DNN Single-outcome model AUC=0.89, accuracy=0.95 (95% CI:0.95-0.96); DNN Multiple-outcome model AUC=0.89, accuracy=0.95 (95% CI:0.95-0.96).

Sensitivity, specificity, PPV and NPV for each outcome (CD grade 0-V) are detailed in supplementary table **10**.

### 3.3 Model Selection and Deployment

Two RF models (transfusion and post-operative infection) selected for deployment in the online application. The application can be visualized at: https://endourology.shinyapps.io/PCNL_Prediction_tool/. Each model has a ‘Local Interpretable Model-Agnostic Explainer’ (‘lime’), this details and ranks by weight the variables the model is using to predict a particular outcome [see figure 3].

**Figure 3.**
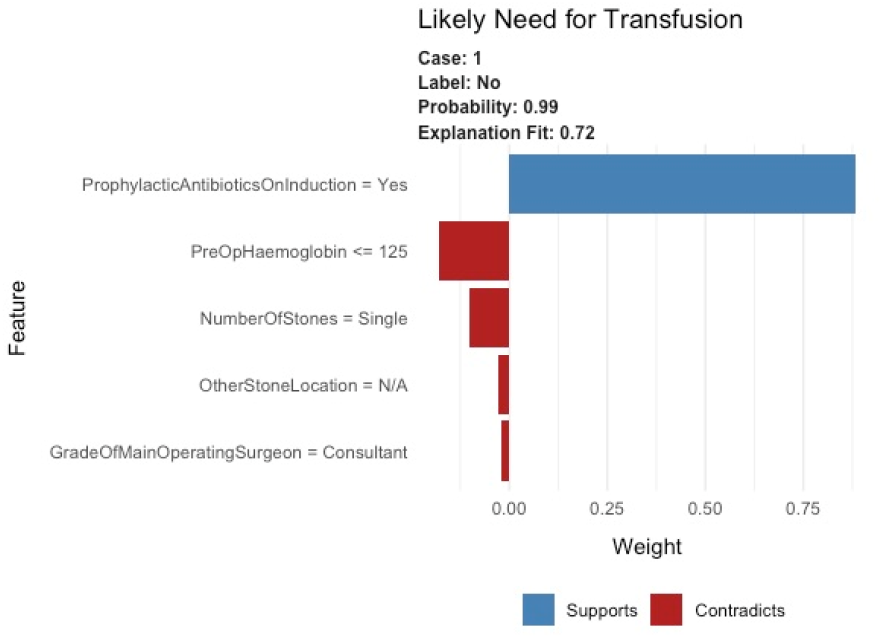
Example explanatory plot using a ‘lime’ explainer. Label = predicted outcome (in this case unlikely to need transfusion), Probability=probability according to the model, Explanation fit=how the particular patient’s characteristics compare to the model’s ideal characteristics for prediction of the outcome. Y-axis=Features ranked by weighting. X-Axis=weighting according to model.

## 4. Discussion

This is the largest machine learning study in PCNL to date, with the largest number of predicted outcomes (n=13), although only 2 of our outcomes (need for transfusion and post-operative infection) are well predicted in our internal validation dataset with high accuracy, sensitivity, specificity and AUC. These two have therefore been included in the online application for external validation. The post-operative stay and Clavien-Dindo (CD) complication classification also had high AUCs, but were not included as they do not accurately predict specific outcomes.

There are several limitations with this study, the main being the rarity of particular outcomes e.g visceral injury or death. This leads to poor prediction of these particular outcomes. There are methods of artificially increasing these outcomes (i.e duplicating entries with particular outcome), but this often to leads to the model overestimating the adverse outcome [22,23]. The addition of imaging data may increase the predictability of visceral injury. Previous studies have demonstrated the use of neural networks to identify renal tumours and their vasculature, which aids operative planning[24].

For most outcomes we demonstrate high accuracies and AUCs. These can be misleading. For example, the visceral injury models mostly report accuracies of 99%, with a maximum AUC of 0.77 (BGLM). This is because the models predict every participant as having ‘no’ visceral injury (sensitivity=1.00, specificity=0.00). With rare outcomes, visceral injury is seen in just 0.3% of patients, summary figures such as ‘accuracy’ and AUC should be discounted in favour of sensitivity and specificity.

Stone free status was poorly predicted. This may be due to loss of data, but more likely, due to differing methods of follow-up imaging. The gold standard of stone free status ascertainment is with CT[25]. In the UK, patients are often followed up with ultrasound or x-ray, which overestimate stone free status[26]. It is also unclear as to the ‘stone free’ definition. Historically, <4mm or <2mm fragments were deemed acceptable and included in the definition of ‘stone free’. However, more recently, this has been challenged. These residual fragments are likely to become clinically significant [27], and therefore the definition of ‘stone free’ has been redefined to ‘no fragments’. This ambiguity about stone free ascertainment and definition are the likely reasons behind poor prediction.

Future studies should externally validate our models for transfusion and post-operative infection. Studies aiming to build machine learning tools for post-PCNL outcome prediction may benefit from the inclusion of imaging data.

## 5. Conclusions

Machine/Deep learning can provide useful tools for prediction of particular outcomes (transfusion/infection) to high levels of diagnostic accuracy. However, in this study, some outcomes are poorly predicted, which suggests an incomplete dataset in some cases, and a very rare outcome in others. Future ML studies should utilize imaging data as well as clinical data.

## Supporting information

See statistical code

See statistical code

## Data Availability

All data produced in the present study are available upon application to the British Association of Urological Surgeons

## Funding

RG is supported by the National Institute for Health Research (NIHR) as an Academic Clinical Fellow.

## Notes

### Competing Interest Statement

The authors have declared no competing interest.

### Funding Statement

This study did not receive any funding

### Author Declarations

This study used data from a previously published national audit (https://doi.org/10.1016/j.eururo.2012.01.003). Access to this dataset was applied for and granted by the British Association of Urological Surgeons. As an audit it does not require Ethical approval as per Health Research Authority (UK).

