## Supplementary material for "Use of Internally Validated Machine and Deep Learning Models to Predict Outcomes of Percutaneous Nephrolithotomy using data from the BAUS PCNL audit": See statistical code

library(shiny)

library(lime)

library(caret)

library(mlbench)

library(gridExtra)

library(ggplot2)

library(MLeval)

transfusion_model<-readRDS("/PCNL_transfusion_rf.rds")

infection_model<-readRDS("/PCNL_Post_Infection_rf.rds")

post_op_stay_model<-readRDS("/PCNL_Stay_xgboost.rds")

clavien_dindo_model<-readRDS("/PCNL_Clavien_rf.rds")

transfusion_explainer<-readRDS("/transfusion_explainer.rds")

infection_explainer<-readRDS("/infection_explainer.rds")

stay_explainer<-readRDS("/stay_explainer.rds")

clavien_explainer<-readRDS("/clavien_explainer.rds")

PCNL_new3<-read.csv("/PCNL_new3.csv")

PCNL_new3$Age<-as.integer(PCNL_new3$Age)

PCNL_new3$Gender<-as.factor(PCNL_new3$Gender)

PCNL_new3$SideOfStones<-as.factor(PCNL_new3$SideOfStones)

PCNL_new3$BMI<-as.numeric(PCNL_new3$BMI)

PCNL_new3$CharlsonScore<-as.integer(PCNL_new3$CharlsonScore)

PCNL_new3$AgeRelatedCharlsonScore<-as.integer(PCNL_new3$AgeRelatedCharlsonScore)

PCNL_new3$PreviousUrinaryTractInfection<-as.factor(PCNL_new3$PreviousUrinaryTractInfection)

PCNL_new3$PreOperativeAntibioticCourse<-as.factor(PCNL_new3$PreOperativeAntibioticCourse)

PCNL_new3$PreOperativeMsu<-as.factor(PCNL_new3$PreOperativeMsu)

PCNL_new3$PreOperativeMsuResult<-as.factor(PCNL_new3$PreOperativeMsuResult)

PCNL_new3$Pre_op_radiology_primary<-as.factor(PCNL_new3$Pre_op_radiology_primary)

PCNL_new3$Pre_op_radiology_secondary<-as.factor(PCNL_new3$Pre_op_radiology_secondary)

PCNL_new3$Renogramdmsa<-as.factor(PCNL_new3$Renogramdmsa)

PCNL_new3$IsThePatientCatheterised<-as.factor(PCNL_new3$IsThePatientCatheterised)

PCNL_new3$PreOpHaemoglobin<-as.numeric(PCNL_new3$PreOpHaemoglobin)

PCNL_new3$PreOpEgfr<-as.factor(PCNL_new3$PreOpEgfr)

PCNL_new3$ProphylacticAntibioticsOnInduction<-as.factor(PCNL_new3$ProphylacticAntibioticsOnInduction)

PCNL_new3$GradeOfMainOperatingSurgeon<-as.factor(PCNL_new3$GradeOfMainOperatingSurgeon)

PCNL_new3$TypeOfAnaesthesia<-as.factor(PCNL_new3$TypeOfAnaesthesia)

PCNL_new3$InterventionalRadiologyBackupForSelectiveRenalArte<-as.factor(PCNL_new3$InterventionalRadiologyBackupForSelectiveRenalArte)

PCNL_new3$SecondaryReLookNephroscopy<-as.factor(PCNL_new3$SecondaryReLookNephroscopy)

PCNL_new3$StoneDimensions<-as.factor(PCNL_new3$StoneDimensions)

PCNL_new3$NumberOfStones<-as.factor(PCNL_new3$NumberOfStones)

PCNL_new3$IndexStoneLocation<-as.factor(PCNL_new3$IndexStoneLocation)

PCNL_new3$OtherStoneLocation<-as.factor(PCNL_new3$OtherStoneLocation)

PCNL_new3$StoneComplexity<-as.factor(PCNL_new3$StoneComplexity)

PCNL_new3$MaximumHounsfieldUnitsOfTheIndexStoneOnCtkub<-as.factor(PCNL_new3$MaximumHounsfieldUnitsOfTheIndexStoneOnCtkub)

PCNL_new3$PreExistingNephrostomyTube<-as.factor(PCNL_new3$PreExistingNephrostomyTube)

PCNL_new3$PunctureTractPerformedBy<-as.factor(PCNL_new3$PunctureTractPerformedBy)

PCNL_new3$GradeOfPerformer<-as.factor(PCNL_new3$GradeOfPerformer)

PCNL_new3$PunctureSite<-as.factor(PCNL_new3$PunctureSite)

PCNL_new3$ImageGuidanceForRenalPuncture<-as.factor(PCNL_new3$ImageGuidanceForRenalPuncture)

PCNL_new3$PatientPosition<-as.factor(PCNL_new3$PatientPosition)

PCNL_new3$NumberOfTractsPlanned<-as.factor(PCNL_new3$NumberOfTractsPlanned)

PCNL_new3$NumberOfTractsPlanned<-as.integer(PCNL_new3$NumberOfTractsPlanned)

PCNL_new3$NumberOfTractsPerformed<-as.integer(PCNL_new3$NumberOfTractsPerformed)

PCNL_new3$PlacementOfTract<-as.factor(PCNL_new3$PlacementOfTract)

PCNL_new3$SizeouterDiameterOfAmplatzSheath<-as.factor(PCNL_new3$SizeouterDiameterOfAmplatzSheath)

PCNL_new3$DilatorsUsed<-as.factor(PCNL_new3$DilatorsUsed)

PCNL_new3$PredictedDifficulty<-as.factor(PCNL_new3$PredictedDifficulty)

PCNL_new3$AccessoryProcedures<-as.factor(PCNL_new3$AccessoryProcedures)

PCNL_new3$NephrostomyDrainInSitu<-as.factor(PCNL_new3$NephrostomyDrainInSitu)

PCNL_new3$PrimaryStoneExtractionFragmentationTechnique<-as.factor(PCNL_new3$PrimaryStoneExtractionFragmentationTechnique)

PCNL_new3$SecondaryStoneExtractionFragmentationTechnique<-as.factor(PCNL_new3$SecondaryStoneExtractionFragmentationTechnique)

\# Define UI

ui <- fluidPage(tabPanel(

\# Application title

titlePanel("PCNL Outcome Prediction Tool"),

tabPanel("Disclaimer/Instructions",

p("This app has been built for external validation purposes only."),

p("Please use the tabs along to navigate the app. The prediction and explanatatory plots will automatically generate when new data is added"),

p("These predictive machine learning tools were built using data from the BAUS PCNL audit"),

p("Below are the ROC curves for each model, along with the area under the curve (AUC) for each model"),

p("Models were selected after extensive testing with 8 different machine learning techniques"),

p("See the link below to our publication explaining model selection"),

p("<link to publication>"),

plotOutput("roc_curves")

),

tabPanel(

"Input Parameters",

fluidRow(

column(3,

sliderInput(inputId = "Age", label = "Age", value = 25, min = 1, max = 100),

sliderInput(inputId = "BMI", label = "BMI", value = 25, min = 1, max = 60),

sliderInput(inputId = "PreOpHaemoglobin", label = "Pre-Operative Haemoglobin (g/L)", value = 130, min = 40, max = 200),

sliderInput(inputId = "CharlsonScore", label = "Charlson Score", value = 1, min = 0, max = 11),

sliderInput(inputId = "AgeRelatedCharlsonScore", label = "Age Related Charlson Score", value = 1, min = 0, max = 11),

sliderInput(inputId = "NumberOfTractsPlanned", label = "Number Of Tracts Planned", value = 1, min = 0, max = 10),

sliderInput(inputId = "NumberOfTractsPerformed", label = "Number Of Tracts Performed", value = 1, min = 0, max = 10),

selectInput(inputId = "Gender", label = "Gender", choices = levels(PCNL_new3$Gender)),

selectInput(inputId = "SideOfStones", label = "Side of Stone(s)", choices = levels(PCNL_new3$SideOfStones)),

selectInput(inputId = "PreviousUrinaryTractInfection", label = "Previous UTI treatment?", choices = levels(PCNL_new3$PreviousUrinaryTractInfection)),

selectInput(inputId = "PreOperativeAntibioticCourse", label = "Pre-Operative Antibiotic Course?", choices = levels(PCNL_new3$PreOperativeAntibioticCourse)),

selectInput(inputId = "PreOperativeMsu", label = "Pre-Operative MSU done?", choices = levels(PCNL_new3$PreOperativeMsu)),

selectInput(inputId = "PreOperativeMsuResult", label = "Pre-Operative MSU Result", choices = levels(PCNL_new3$PreOperativeMsuResult)),

offset = 1

),

column(3,

selectInput(inputId = "Pre_op_radiology_primary", label = "Primary Pre-Operative Imaging", choices = levels(PCNL_new3$Pre_op_radiology_primary)),

selectInput(inputId = "Pre_op_radiology_secondary", label = "Secondary Pre-Operative Imaging", choices = levels(PCNL_new3$Pre_op_radiology_secondary)),

selectInput(inputId = "Renogramdmsa", label = "Pre-Operative Renogram (DMSA)?", choices = levels(PCNL_new3$Renogramdmsa)),

selectInput(inputId = "IsThePatientCatheterised", label = "Is The Patient Catheterised?", choices = levels(PCNL_new3$IsThePatientCatheterised)),

selectInput(inputId = "PreOpEgfr", label = "Pre-Operative eGFR", choices = levels(PCNL_new3$PreOpEgfr)),

selectInput(inputId = "ProphylacticAntibioticsOnInduction", label = "Prophylactic Antibiotics On Induction Planned?", choices = levels(PCNL_new3$ProphylacticAntibioticsOnInduction)),

selectInput(inputId = "GradeOfMainOperatingSurgeon", label = "Grade Of Main Operating Surgeon", choices = levels(PCNL_new3$GradeOfMainOperatingSurgeon)),

selectInput(inputId = "TypeOfAnaesthesia", label = "Type Of Anaesthesia Planned", choices = levels(PCNL_new3$TypeOfAnaesthesia)),

selectInput(inputId = "InterventionalRadiologyBackupForSelectiveRenalArte", label = "Interventional Radiology Backup Available?", choices = levels(PCNL_new3$InterventionalRadiologyBackupForSelectiveRenalArte)),

selectInput(inputId = "SecondaryReLookNephroscopy", label = "Secondary Re-look Nephroscopy Planned?", choices = levels(PCNL_new3$SecondaryReLookNephroscopy)),

selectInput(inputId = "StoneDimensions", label = "Stone Dimensions (cm)", choices = levels(PCNL_new3$StoneDimensions)),

selectInput(inputId = "NumberOfStones", label = "Number Of Stones", choices = levels(PCNL_new3$NumberOfStones)),

selectInput(inputId = "IndexStoneLocation", label = "Index Stone Location", choices = levels(PCNL_new3$IndexStoneLocation)),

selectInput(inputId = "OtherStoneLocation", label = "Other Stone Location", choices = levels(PCNL_new3$OtherStoneLocation)),

selectInput(inputId = "StoneComplexity", label = "Guy's Stone Score", choices = levels(PCNL_new3$StoneComplexity)),

offset = 1

),

column(3,

selectInput(inputId = "MaximumHounsfieldUnitsOfTheIndexStoneOnCtkub", label = "Maximum Hounsfield Units Of Index Stone On CT KUB", choices = levels(PCNL_new3$MaximumHounsfieldUnitsOfTheIndexStoneOnCtkub)),

selectInput(inputId = "PreExistingNephrostomyTube", label = "Pre Existing Nephrostomy Tube", choices = levels(PCNL_new3$PreExistingNephrostomyTube)),

selectInput(inputId = "PunctureTractPerformedBy", label = "Puncture Tract Performed By?", choices = levels(PCNL_new3$PunctureTractPerformedBy)),

selectInput(inputId = "GradeOfPerformer", label = "Planned Grade Of Puncture Performer", choices = levels(PCNL_new3$GradeOfPerformer)),

selectInput(inputId = "PunctureSite", label = "Planned Puncture Site", choices = levels(PCNL_new3$PunctureSite)),

selectInput(inputId = "ImageGuidanceForRenalPuncture", label = "Planned Image Guidance For Renal Puncture", choices = levels(PCNL_new3$ImageGuidanceForRenalPuncture)),

selectInput(inputId = "PatientPosition", label = "Planned Patient Position", choices = levels(PCNL_new3$PatientPosition)),

selectInput(inputId = "PlacementOfTract", label = "Planned Placement Of Tract", choices = levels(PCNL_new3$PlacementOfTract)),

selectInput(inputId = "SizeouterDiameterOfAmplatzSheath", label = "Size/Outer Diameter Of Amplatz Sheath (Fr)", choices = levels(PCNL_new3$SizeouterDiameterOfAmplatzSheath)),

selectInput(inputId = "DilatorsUsed", label = "Planned Dilators", choices = levels(PCNL_new3$DilatorsUsed)),

selectInput(inputId = "PredictedDifficulty", label = "Predicted Difficulty", choices = levels(PCNL_new3$PredictedDifficulty)),

selectInput(inputId = "AccessoryProcedures", label = "Planned Accessory Procedures", choices = levels(PCNL_new3$AccessoryProcedures)),

selectInput(inputId = "NephrostomyDrainInSitu", label = "Planned Post-Operative Nephrostomy", choices = levels(PCNL_new3$NephrostomyDrainInSitu)),

selectInput(inputId = "PrimaryStoneExtractionFragmentationTechnique", label = "Primary Stone Extraction/Fragmentation Technique", choices = levels(PCNL_new3$PrimaryStoneExtractionFragmentationTechnique)),

selectInput(inputId = "SecondaryStoneExtractionFragmentationTechnique", label = "Secondary Stone Extraction/Fragmentation Technique", choices = levels(PCNL_new3$SecondaryStoneExtractionFragmentationTechnique)),

offset = 1

)

),

),

tabPanel("Prediction Table",

p("These predictions are generated using the inputted data"),

tableOutput("table")

),

tabPanel(

"Prediction Explanation Plots",

p("These graphs correspond to each of the outcome predictions"),

p("Each plot below corresponds to the prediction in the 'Prediction Table', the 'Label' on each graph has the same prediction "),

p("The 'Probability' should be read as the likelihood of the outcome being true according to the model (x100 for percentage)"),

p("The 'Explanation Fit' should be read as how closely the model fits to the inputted data for that particular outcome (x100 for percentage)"),

p("The 'Weight' on the x-axis corresponds to the weight allotted by the model to the particular feature"),

p("The 'Feature' allotted to the y-axis, corresponds to the inputted parameters. The features are ranked from most to least weight i.e impact on the predicted outcome"),

p("The 'number of features' for each graphs can be toggled here"),

sliderInput(inputId = "no_features", label = "Number of Features for Explanation Plot", value = 5, min = 1, max = 42),

plotOutput("combined_limeplot"))

))

server <- function(input, output) {

output$roc_curves<-renderPlot({

transfusion_model_roc<-evalm(transfusion_model)

infection_model_roc<-evalm(infection_model)

post_op_stay_model_roc<-evalm(post_op_stay_model)

clavien_dindo_model_roc<-evalm(clavien_dindo_model)

transfusion_model_roc1<-transfusion_model_roc$roc +ggtitle("Transfusion - Random Forests")

infection_model_roc1<-infection_model_roc$roc + ggtitle("Post-Operative Infection - Random Forests")

post_op_stay_model_roc1<-post_op_stay_model_roc$roc +ggtitle("Post-Operative Stay (days) - XGBoost")

clavien_dindo_model_roc1<-clavien_dindo_model_roc$roc +ggtitle("Clavien Dindo Classification - Random Forests")

grid.arrange(transfusion_model_roc1, infection_model_roc1, post_op_stay_model_roc1, clavien_dindo_model_roc1)

})

getData <- reactive({

data.frame(

Age = as.integer(input$Age),

BMI = as.integer(input$BMI),

PreOpHaemoglobin = as.integer(input$PreOpHaemoglobin),

CharlsonScore = as.integer(input$CharlsonScore),

AgeRelatedCharlsonScore = as.integer(input$AgeRelatedCharlsonScore),

NumberOfTractsPlanned = as.integer(input$NumberOfTractsPlanned),

NumberOfTractsPerformed=as.integer(input$NumberOfTractsPerformed),

Gender = as.factor(input$Gender),

SideOfStones= as.factor(input$SideOfStones),

PreviousUrinaryTractInfection= as.factor(input$PreviousUrinaryTractInfection),

PreOperativeAntibioticCourse= as.factor(input$PreOperativeAntibioticCourse),

PreOperativeMsu= as.factor(input$PreOperativeMsu),

PreOperativeMsuResult= as.factor(input$PreOperativeMsuResult),

Pre_op_radiology_primary= as.factor(input$Pre_op_radiology_primary),

Pre_op_radiology_secondary= as.factor(input$Pre_op_radiology_secondary),

Renogramdmsa= as.factor(input$Renogramdmsa),

IsThePatientCatheterised= as.factor(input$IsThePatientCatheterised),

PreOpEgfr= as.factor(input$PreOpEgfr),

ProphylacticAntibioticsOnInduction= as.factor(input$ProphylacticAntibioticsOnInduction),

GradeOfMainOperatingSurgeon= as.factor(input$GradeOfMainOperatingSurgeon),

TypeOfAnaesthesia= as.factor(input$TypeOfAnaesthesia),

InterventionalRadiologyBackupForSelectiveRenalArte= as.factor(input$InterventionalRadiologyBackupForSelectiveRenalArte),

SecondaryReLookNephroscopy= as.factor(input$SecondaryReLookNephroscopy),

StoneDimensions= as.factor(input$StoneDimensions),

NumberOfStones= as.factor(input$NumberOfStones),

IndexStoneLocation= as.factor(input$IndexStoneLocation),

OtherStoneLocation= as.factor(input$OtherStoneLocation),

StoneComplexity= as.factor(input$StoneComplexity),

MaximumHounsfieldUnitsOfTheIndexStoneOnCtkub= as.factor(input$MaximumHounsfieldUnitsOfTheIndexStoneOnCtkub),

PreExistingNephrostomyTube= as.factor(input$PreExistingNephrostomyTube),

PunctureTractPerformedBy= as.factor(input$PunctureTractPerformedBy),

GradeOfPerformer= as.factor(input$GradeOfPerformer),

PunctureSite= as.factor(input$PunctureSite),

ImageGuidanceForRenalPuncture= as.factor(input$ImageGuidanceForRenalPuncture),

PatientPosition= as.factor(input$PatientPosition),

PlacementOfTract= as.factor(input$PlacementOfTract),

SizeouterDiameterOfAmplatzSheath= as.factor(input$SizeouterDiameterOfAmplatzSheath),

DilatorsUsed= as.factor(input$DilatorsUsed),

PredictedDifficulty= as.factor(input$PredictedDifficulty),

AccessoryProcedures= as.factor(input$AccessoryProcedures),

NephrostomyDrainInSitu= as.factor(input$NephrostomyDrainInSitu),

PrimaryStoneExtractionFragmentationTechnique= as.factor(input$PrimaryStoneExtractionFragmentationTechnique),

SecondaryStoneExtractionFragmentationTechnique= as.factor(input$SecondaryStoneExtractionFragmentationTechnique)

)

}

)

no_features<-reactive({

no_features = as.integer(input$no_features)

})

output$table <- renderTable({

transfusion_prediction<-predict(transfusion_model, newdata=getData())

transfusion_prediction1<-as.data.frame((transfusion_prediction))

colnames(transfusion_prediction1)<-c("Likely Need for Transfusion")

infection_prediction<-predict(infection_model, newdata=getData())

infection_prediction1<-as.data.frame((infection_prediction))

colnames(infection_prediction1)<-c("Likely Post-Operative Infection")

post_op_stay_prediction<-predict(post_op_stay_model, newdata=getData())

post_op_stay_prediction1<-as.data.frame((post_op_stay_prediction))

colnames(post_op_stay_prediction1)<-c("Likely Post-Operative Stay (days)")

clavien_dindo_prediction<-predict(clavien_dindo_model, newdata=getData())

clavien_dindo_prediction1<-as.data.frame((clavien_dindo_prediction))

colnames(clavien_dindo_prediction1)<-c("Likely Clavien Dindo Classification of Expected Post-operative complication")

combined_table<-cbind(transfusion_prediction1, infection_prediction1, post_op_stay_prediction1, clavien_dindo_prediction1)

combined_table

}

)

output$combined_limeplot <- renderPlot({

transfusion_explanation <- lime::explain(getData(), transfusion_explainer, n_labels = 1, n_features = input$no_features)

transfusion_limeplot<-plot_features(transfusion_explanation) + ggtitle("Likely Need for Transfusion")

infection_explanation <- lime::explain(getData(), infection_explainer, n_labels = 1, n_features = input$no_features)

infection_limeplot<-plot_features(infection_explanation) + ggtitle("Likely Post-Operative Infection")

stay_explanation <- lime::explain(getData(), stay_explainer, n_labels = 1, n_features = input$no_features)

stay_limeplot<-plot_features(stay_explanation) + ggtitle("Likely Post-Operative Stay (days)")

clavien_explanation <- lime::explain(getData(), clavien_explainer, n_labels = 1, n_features = input$no_features)

clavien_dindo_limeplot<-plot_features(clavien_explanation) + ggtitle("Likely Clavien Dindo Classification \nof Expected Post-operative complication")

grid.arrange(transfusion_limeplot, infection_limeplot, stay_limeplot, clavien_dindo_limeplot)

})

}

\# Run the application

shinyApp(ui = ui, server = server)
