## Supplementary material for "Use of Internally Validated Machine and Deep Learning Models to Predict Outcomes of Percutaneous Nephrolithotomy using data from the BAUS PCNL audit": See statistical code

**R Geraghty**

Table of Contents

|  | Section | Page |
| --- | --- | --- |
| Set up + Import data | 1 | 3 |
| Adjust/Sort file for specific outcomes | 2 | 4-8 |
| Multiple Output DL model | 3 | 9-26 |
| Immediate Clearance on Fluoroscopy in Theatre | 4 | 27-39 |
| Visceral Injury | 5 | 40-49 |
| Survival | 6 | 50-59 |
| Transfusion | 7 | 60-68 |
| Post-operative Infection | 8 | 69-77 |
| Intra-operative Complications | 9 | 78-86 |
| Need for ITU/HDU | 10 | 87-95 |
| Stone Free at Follow-up | 11 | 96-104 |
| Need for Adjuvant treatment | 12 | 105-113 |
| Post-operative stay | 13 | 114-124 |
| Clavien Dindo Classification | 14 | 125-139 |
| Truncated Multiple Output DL model | 15 | 140-143 |
| ROC curves for DL models | 16 | 144-146 |

1. Set up + Import data

library(keras)

library(tensorflow)

library(lime)

library(dplyr)

library(tidyverse)

library(caret)

library(pROC)

library(arm)

library(MLeval)

PCNL_new2<-read.csv("PCNL_new2.csv", header=TRUE)

2. Adjust/Sort file for specific outcomes

PCNL_new2$Age<-as.integer(PCNL_new2$Age)

PCNL_new2$Gender<-as.factor(PCNL_new2$Gender)

PCNL_new2$Year<-as.integer(PCNL_new2$Year)

PCNL_new2$SideOfStones<-as.factor(PCNL_new2$SideOfStones)

PCNL_new2$BMI<-as.numeric(PCNL_new2$BMI)

PCNL_new2$CharlsonScore<-as.integer(PCNL_new2$CharlsonScore)

PCNL_new2$AgeRelatedCharlsonScore<-as.integer(PCNL_new2$AgeRelatedCharlsonScore)

PCNL_new2$PreviousUrinaryTractInfection<-as.factor(PCNL_new2$PreviousUrinaryTractInfection)

PCNL_new2$PreOperativeAntibioticCourse<-as.factor(PCNL_new2$PreOperativeAntibioticCourse)

PCNL_new2$PreOperativeMsu<-as.factor(PCNL_new2$PreOperativeMsu)

PCNL_new2$PreOperativeMsuResult<-as.factor(PCNL_new2$PreOperativeMsuResult)

PCNL_new2$Pre_op_radiology_primary<-as.factor(PCNL_new2$Pre_op_radiology_primary)

PCNL_new2$Pre_op_radiology_secondary<-as.factor(PCNL_new2$Pre_op_radiology_secondary)

PCNL_new2$Renogramdmsa<-as.factor(PCNL_new2$Renogramdmsa)

PCNL_new2$IsThePatientCatheterised<-as.factor(PCNL_new2$IsThePatientCatheterised)

PCNL_new2$PreOpHaemoglobin<-as.numeric(PCNL_new2$PreOpHaemoglobin)

PCNL_new2$PreOpEgfr<-as.factor(PCNL_new2$PreOpEgfr)

PCNL_new2$ProphylacticAntibioticsOnInduction<-as.factor(PCNL_new2$ProphylacticAntibioticsOnInduction)

PCNL_new2$GradeOfMainOperatingSurgeon<-as.factor(PCNL_new2$GradeOfMainOperatingSurgeon)

PCNL_new2$TypeOfAnaesthesia<-as.factor(PCNL_new2$TypeOfAnaesthesia)

PCNL_new2$InterventionalRadiologyBackupForSelectiveRenalArte<-as.factor(PCNL_new2$InterventionalRadiologyBackupForSelectiveRenalArte)

PCNL_new2$SecondaryReLookNephroscopy<-as.factor(PCNL_new2$SecondaryReLookNephroscopy)

PCNL_new2$StoneDimensions<-as.factor(PCNL_new2$StoneDimensions)

PCNL_new2$NumberOfStones<-as.factor(PCNL_new2$NumberOfStones)

PCNL_new2$IndexStoneLocation<-as.factor(PCNL_new2$IndexStoneLocation)

PCNL_new2$OtherStoneLocation<-as.factor(PCNL_new2$OtherStoneLocation)

PCNL_new2$StoneComplexity<-as.factor(PCNL_new2$StoneComplexity)

PCNL_new2$MaximumHounsfieldUnitsOfTheIndexStoneOnCtkub<-as.factor(PCNL_new2$MaximumHounsfieldUnitsOfTheIndexStoneOnCtkub)

PCNL_new2$PreExistingNephrostomyTube<-as.factor(PCNL_new2$PreExistingNephrostomyTube)

PCNL_new2$PunctureTractPerformedBy<-as.factor(PCNL_new2$PunctureTractPerformedBy)

PCNL_new2$GradeOfPerformer<-as.factor(PCNL_new$GradeOfPerformer)

PCNL_new2$PunctureSite<-as.factor(PCNL_new2$PunctureSite)

PCNL_new2$ImageGuidanceForRenalPuncture<-as.factor(PCNL_new2$ImageGuidanceForRenalPuncture)

PCNL_new2$PatientPosition<-as.factor(PCNL_new2$PatientPosition)

PCNL_new2$NumberOfTractsPlanned<-as.factor(PCNL_new2$NumberOfTractsPlanned)

PCNL_new2$NumberOfTractsPlanned<-as.integer(PCNL_new2$NumberOfTractsPlanned)

PCNL_new2$NumberOfTractsPerformed<-as.integer(PCNL_new2$NumberOfTractsPerformed)

PCNL_new2$PlacementOfTract<-as.factor(PCNL_new2$PlacementOfTract)

PCNL_new2$SizeouterDiameterOfAmplatzSheath<-as.factor(PCNL_new2$SizeouterDiameterOfAmplatzSheath)

PCNL_new2$DilatorsUsed<-as.factor(PCNL_new2$DilatorsUsed)

PCNL_new2$DifficultAccess<-as.factor(PCNL_new2$DifficultAccess)

PCNL_new2$PredictedDifficulty<-as.factor(PCNL_new2$PredictedDifficulty)

PCNL_new2$AccessoryProcedures<-as.factor(PCNL_new2$AccessoryProcedures)

PCNL_new2$UretericCatheterPostOperatively<-as.factor(PCNL_new2$UretericCatheterPostOperatively)

PCNL_new2$PrimaryStoneExtractionFragmentationTechnique<-as.factor(PCNL_new2$PrimaryStoneExtractionFragmentationTechnique)

PCNL_new2$SecondaryStoneExtractionFragmentationTechnique<-as.factor(PCNL_new2$SecondaryStoneExtractionFragmentationTechnique)

PCNL_new2$CompleteClearanceOnFluoroscopy<-as.factor(PCNL_new2$CompleteClearanceOnFluoroscopy)

PCNL_new2$ProcedureAbandoned<-as.factor(PCNL_new2$ProcedureAbandoned)

PCNL_new2$VisceralInjury<-as.factor(PCNL_new2$VisceralInjury)

PCNL_new2$NephrostomyDrainInSitu<-as.factor(PCNL_new2$NephrostomyDrainInSitu)

PCNL_new2$UretericCatheterPostOperatively<-as.factor(PCNL_new2$UretericCatheterPostOperatively)

PCNL_new2$StentInsertedPostOperatively<-as.factor(PCNL_new2$StentInsertedPostOperatively)

PCNL_new2$ClearanceOnPostOperativeRadiologicalImagingDuringA<-as.factor(PCNL_new2$ClearanceOnPostOperativeRadiologicalImagingDuringA)

PCNL_new2$PatientStatusdischarge<-as.factor(PCNL_new2$PatientStatusdischarge)

PCNL_new2$ClavienDindoGradeOfComplications<-as.factor(PCNL_new2$ClavienDindoGradeOfComplications)

PCNL_new2$BloodTransfusion<-as.factor(PCNL_new2$BloodTransfusion)

PCNL_new2$PostOperativeInfection<-as.factor(PCNL_new2$PostOperativeInfection)

PCNL_new2$PostOperativeStay<-as.integer(PCNL_new2$PostOperativeStay)

PCNL_new2$IntraopComplications<-as.factor(PCNL_new2$IntraopComplications)

PCNL_new2$ItuHduAdmission<-as.factor(PCNL_new2$ItuHduAdmission)

PCNL_new2$StoneFreeAtFollowUp<-as.factor(PCNL_new2$StoneFreeAtFollowUp)

PCNL_new2$AdjuvantTreatment<-as.factor(PCNL_new2$AdjuvantTreatment)

PCNL_new2$ Gender_coded<-as.integer(PCNL_new2$ Gender_coded)

PCNL_new2$Side_coded<- as.integer (PCNL_new2$Side_coded)

PCNL_new2$Previous_UTI_coded<-as.integer(PCNL_new2$Previous_UTI_coded)

PCNL_new2$Pre_Op_abx_coded<-as.integer(PCNL_new2$Pre_Op_abx_coded)

PCNL_new2$Pre_Op_MSU_coded<-as.integer(PCNL_new2$Pre_Op_MSU_coded)

PCNL_new2$Pre_Op_MSU_result_coded<-as.integer(PCNL_new2$Pre_Op_MSU_result_coded)

PCNL_new2$Pre_op_rad_prim_coded<-as.integer(PCNL_new2$Pre_op_rad_prim_coded)

PCNL_new2$Pre_op_radiology_secondary_coded<-as.integer(PCNL_new2$Pre_op_radiology_secondary_coded)

PCNL_new2$DMSA_coded<-as.integer(PCNL_new2$DMSA_coded)

PCNL_new2$Catheter_coded<-as.integer(PCNL_new2$Catheter_coded)

PCNL_new2$GFR_coded<-as.integer(PCNL_new2$GFR_coded)

PCNL_new2$Induction_Abx_coded<-as.integer(PCNL_new2$Induction_Abx_coded)

PCNL_new2$Surgeon_Grade_coded<-as.integer(PCNL_new2$Surgeon_Grade_coded)

PCNL_new2$Anaesthesia_coded<- as.integer(PCNL_new2$Anaesthesia_coded)

PCNL_new2$IR_Backup_coded<-as.integer(PCNL_new2$IR_Backup_coded)

PCNL_new2$Size_coded<-as.integer(PCNL_new2$Size_coded)

PCNL_new2$Number_stones_coded<-as.integer(PCNL_new2$Number_stones_coded)

PCNL_new2$Index_stone_coded<-as.integer(PCNL_new2$Index_stone_coded)

PCNL_new2$Other_location_coded<-as.integer(PCNL_new2$Other_location_coded)

PCNL_new2$GSS<-as.integer(PCNL_new2$GSS)

PCNL_new2$HU_coded<-as.integer(PCNL_new2$HU_coded)

PCNL_new2$Pre_existing_PCN_coded<-as.integer(PCNL_new2$Pre_existing_PCN_coded)

PCNL_new2$Tract_Performer_coded<-as.integer(PCNL_new2$Tract_Performer_coded)

PCNL_new2$Tract_performer_grade_coded<-as.integer(PCNL_new2$Tract_performer_grade_coded)

PCNL_new2$Puncture_site_coded<-as.integer(PCNL_new2$Puncture_site_coded)

PCNL_new2$Image_guidance_puncture_coded<-as.integer(PCNL_new2$Image_guidance_puncture_coded)

PCNL_new2$PatientPosition_coded<-as.integer(PCNL_new2$PatientPosition_coded)

PCNL_new2$Tract_placement_coded<-as.integer(PCNL_new2$Tract_placement_coded)

PCNL_new2$Amplatz_size_coded<-as.integer(PCNL_new2$Amplatz_size_coded)

PCNL_new2$Dilators_coded<-as.integer(PCNL_new2$Dilators_coded)

PCNL_new2$Predicted_difficulty_coded<-as.integer(PCNL_new2$Predicted_difficulty_coded)

PCNL_new2$Accessory_procedures_coded<-as.integer(PCNL_new2$Accessory_procedures_coded)

PCNL_new2$PCN_post_procedure_coded<-as.integer(PCNL_new2$PCN_post_procedure_coded)

PCNL_new2$Primary_technique_coded<-as.integer(PCNL_new2$Primary_technique_coded)

PCNL_new2$Secondary_technique_coded<-as.integer(PCNL_new2$Secondary_technique_coded)

PCNL_new2$Difficult_access_coded<-as.integer(PCNL_new2$Difficult_access_coded)

PCNL_new2$Post_op_stent_coded<-as.integer(PCNL_new2$Post_op_stent_coded)

PCNL_new2$Post_op_ureteric_coded<-as.integer(PCNL_new2$Post_op_ureteric_coded)

PCNL_new2$Fluoro_complete_clearance_coded<-as.integer(PCNL_new2$Fluoro_complete_clearance_coded)

PCNL_new2$Proc_abandoned_coded<-as.integer(PCNL_new2$Proc_abandoned_coded)

PCNL_new2$Visc_inj_coded<-as.integer(PCNL_new2$Visc_inj_coded)

PCNL_new2$Clearance_admission_imaging_coded<-as.integer(PCNL_new2$Clearance_admission_imaging_coded)

PCNL_new2$Alive_Dead<-as.integer(PCNL_new2$Alive_Dead)

PCNL_new2$CD_complication_coded<-as.integer(PCNL_new2$CD_complication_coded)

PCNL_new2$Transfusion_coded<-as.integer(PCNL_new2$Transfusion_coded)

PCNL_new2$Post_infection_coded<-as.integer(PCNL_new2$Post_infection_coded)

PCNL_new2$Intra_op_comp_coded<-as.integer(PCNL_new2$Intra_op_comp_coded)

PCNL_new2$ITU_HDU_admission_coded<-as.integer(PCNL_new2$ITU_HDU_admission_coded)

PCNL_new2$SF_FU_coded<-as.integer(PCNL_new2$SF_FU_coded)

PCNL_new2$Adjuvant_treatment_coded<-as.integer(PCNL_new2$Adjuvant_treatment_coded)

PCNL_new2$Stay_coded<-as.integer(PCNL_new2$Stay_coded)

PCNL_new2$Stay_words<-as.factor(PCNL_new2$Stay_words)

#Factors

PCNL_1_8<-PCNL_new2[,-9:-111]

PCNL_factors_coded<-PCNL_new2[,-44:-111]

PCNL_80<- PCNL_new2[,-80:-111]

PCNL_factors_non_coded <- PCNL_80[,-9:-44]

\# Non coded Outcome + Coded/Non coded factors:

Immediate_clearance<-PCNL_new2$CompleteClearanceOnFluoroscopy

PCNL_immediate_clearance_coded<-cbind(PCNL_factors_coded, Immediate_clearance)

PCNL_immediate_clearance_non_coded<-cbind(PCNL_factors_non_coded, Immediate_clearance)

Visc_injury<-PCNL_new2$VisceralInjury

PCNL_Visc_injury_coded<- cbind(PCNL_factors_coded, Visc_injury)

PCNL_Visc_injury_non_coded<- cbind(PCNL_factors_non_coded, Visc_injury)

Survival<-PCNL_new2$Alive_Dead

PCNL_survival_coded<-cbind(PCNL_factors_coded, Survival)

PCNL_survival_non_coded<- cbind(PCNL_factors_non_coded, Survival)

Clavien<-PCNL_new2$ClavienDindoGradeOfComplications

PCNL_Clavien_coded<- cbind(PCNL_factors_coded, Clavien)

PCNL_Clavien_non_coded<- cbind(PCNL_factors_non_coded, Clavien)

Transfusion<-PCNL_new2$BloodTransfusion

PCNL_transfusion_coded<- cbind(PCNL_factors_coded, Transfusion)

PCNL_transfusion_non_coded<- cbind(PCNL_factors_non_coded, Transfusion)

Post_infection<-PCNL_new2$PostOperativeInfection

PCNL_Post_Infection_coded<- cbind(PCNL_factors_coded, Post_infection)

PCNL_Post_Infection_non_coded<- cbind(PCNL_factors_non_coded, Post_infection)

Post_stay<-PCNL_new2$PostOperativeStay

PCNL_Post_Stay_coded<- cbind(PCNL_factors_coded, Post_stay)

PCNL_Post_Stay_non_coded<- cbind(PCNL_factors_non_coded, Post_stay)

Intra_complications<-PCNL_new2$IntraopComplications

PCNL_Intra_Complication_coded<- cbind(PCNL_factors_coded, Intra_complications)

PCNL_Intra_Complication_non_coded<- cbind(PCNL_factors_non_coded, Intra_complications)

PCNL_Intra_Complication_coded1<-na.omit(PCNL_Intra_Complication_coded)

PCNL_Intra_Complication_non_coded1<-na.omit(PCNL_Intra_Complication_non_coded)

ITU_HDU<-PCNL_new2$ItuHduAdmission

PCNL_ITU_HDU_coded<- cbind(PCNL_factors_coded, ITU_HDU)

PCNL_ITU_HDU_non_coded<- cbind(PCNL_factors_non_coded, ITU_HDU)

PCNL_ITU_HDU_coded1<-na.omit(PCNL_ITU_HDU_coded)

PCNL_ITU_HDU_non_coded1<-na.omit(PCNL_ITU_HDU_non_coded)

SF<-PCNL_new2$StoneFreeAtFollowUp

PCNL_SF_coded<- cbind(PCNL_factors_coded, SF)

PCNL_SF_non_coded<- cbind(PCNL_factors_non_coded, SF)

PCNL_SF_coded1<-na.omit(PCNL_SF_coded)

PCNL_SF_non_coded1<-na.omit(PCNL_SF_non_coded)

Adjuvant<-PCNL_new2$AdjuvantTreatment

PCNL_Adjuvant_coded<- cbind(PCNL_factors_coded, Adjuvant)

PCNL_Adjuvant_non_coded<- cbind(PCNL_factors_non_coded, Adjuvant)

#Outcomes – Coded:

Immediate_clearance_coded2<-PCNL_new2$Fluoro_complete_clearance_coded

PCNL_immediate_clearance_coded2<-cbind(PCNL_factors_coded, Immediate_clearance_coded2)

Visc_injury_coded<-PCNL_new2$Visc_inj_coded

PCNL_Visc_injury_coded2<- cbind(PCNL_factors_coded, Visc_injury_coded)

Survival<-PCNL_new2$Alive_Dead

PCNL_survival_coded2<- cbind(PCNL_factors_coded, Survival)

Clavien2<-PCNL_new2$CD_complication_coded

PCNL_Clavien_coded2<- cbind(PCNL_factors_coded, Clavien2)

Transfusion2<-PCNL_new2$Transfusion_coded

PCNL_transfusion_coded2<- cbind(PCNL_factors_coded, Transfusion2)

Post_infection2<-PCNL_new2$Post_infection_coded

PCNL_Post_Infection_coded2<- cbind(PCNL_factors_coded, Post_infection2)

Post_stay<-PCNL_new2$PostOperativeStay

PCNL_Post_Stay_coded2<- cbind(PCNL_factors_coded, Post_stay)

Intra_complications2<-PCNL_new2$Intra_op_comp_coded

PCNL_Intra_Complication_coded2<- cbind(PCNL_factors_coded, Intra_complications2)

PCNL_Intra_Complication_coded3<-na.omit(PCNL_Intra_Complication_coded2)

ITU_HDU2<-PCNL_new2$ITU_HDU_admission_coded

PCNL_ITU_HDU_coded2<- cbind(PCNL_factors_coded, ITU_HDU2)

PCNL_ITU_HDU_coded3<-na.omit(PCNL_ITU_HDU_coded2)

SF2<-PCNL_new2$SF_FU_coded

PCNL_SF_coded2<- cbind(PCNL_factors_coded, SF2)

PCNL_SF_coded3<-na.omit(PCNL_SF_coded2)

Adjuvant2<-PCNL_new2$Adjuvant_treatment_coded

PCNL_Adjuvant_coded2<- cbind(PCNL_factors_coded, Adjuvant2)

Stay1<-PCNL_new2$Stay_coded

PCNL_Stay_coded<-cbind(PCNL_factors_coded, Stay1)

Stay2<-as.factor(PCNL_new2$Stay_words)

PCNL_Stay_coded2<-cbind(PCNL_factors_coded, Stay2)

3. Multiple Output DL model

#Outcomes - combined for multiple output model

Immediate_clearance_coded3<-ifelse(Immediate_clearance=="Yes", 1,0)

Visc_injury_coded3<-ifelse(Visc_injury=="Yes", 1,0)

Survival_coded3<-ifelse(Survival=="Yes", 1,0)

Transfusion_coded3<-ifelse(Transfusion=="Yes", 1,0)

Post_infection_coded3<-ifelse(Post_infection=="Yes", 1, 0)

Intra_complications_coded3<-ifelse(Intra_complications=="Yes", 1,0)

ITU_HDU_coded3<-ifelse(ITU_HDU=="Yes",1,0)

SF_coded3<-ifelse(SF=="Yes",1,0)

Adjuvant_coded3<-ifelse(Adjuvant=="Yes",1,0)

PCNL_multiple_output_data<-na.omit(cbind(PCNL_factors_coded, Immediate_clearance_coded3, Visc_injury_coded3, Survival_coded3, Transfusion_coded3, Post_infection_coded3, Intra_complications_coded3, ITU_HDU_coded3, SF_coded3, Adjuvant_coded3))

PCNL_multiple_output_data1<-na.omit(cbind(PCNL_factors_coded, Immediate_clearance_coded3, Visc_injury_coded3, Survival_coded3, Transfusion_coded3, Post_infection_coded3, Intra_complications_coded3, ITU_HDU_coded3, SF_coded3, Adjuvant_coded3,Clavien2, Stay1))

PCNL_multiple_output_sample<-sample(1:nrow(PCNL_multiple_output_data1), size=nrow(PCNL_multiple_output_data1) *0.7)

PCNL_multiple_output_train<-PCNL_multiple_output_data1[PCNL_multiple_output_sample,]

PCNL_multiple_output_test<-PCNL_multiple_output_data1[-PCNL_multiple_output_sample,]

PCNL_multiple_output_train_predictors<-PCNL_multiple_output_train[,-44:-64]

PCNL_multiple_output_test_predictors<-PCNL_multiple_output_test[,-44:-64]

PCNL_multiple_output_train_outcomes<-PCNL_multiple_output_train[,-1:-43]

PCNL_multiple_output_test_outcomes<-PCNL_multiple_output_test[,-1:-43]

PCNL_multiple_output_train_predictors1<-as.matrix(PCNL_multiple_output_train_predictors)

PCNL_multiple_output_test_predictors1<-as.matrix(PCNL_multiple_output_test_predictors)

Immediate_clearance_coded4_train<-as.matrix(PCNL_multiple_output_train_outcomes$Immediate_clearance_coded3)

Visc_injury_coded4_train<-as.matrix(PCNL_multiple_output_train_outcomes$Visc_injury_coded3)

Survival_coded4_train<-as.matrix(PCNL_multiple_output_train_outcomes$Survival_coded3)

Transfusion_coded4_train<-as.matrix(PCNL_multiple_output_train_outcomes$Transfusion_coded3)

Post_infection_coded4_train<-as.matrix(PCNL_multiple_output_train_outcomes$Post_infection_coded3)

Intra_complications_coded4_train<-as.matrix(PCNL_multiple_output_train_outcomes$Intra_complications_coded3)

ITU_HDU_coded4_train<-as.matrix(PCNL_multiple_output_train_outcomes$ITU_HDU_coded3)

SF_coded4_train<-as.matrix(PCNL_multiple_output_train_outcomes$SF_coded3)

Adjuvant_coded4_train<-as.matrix(PCNL_multiple_output_train_outcomes$Adjuvant_coded3)

Post_stay4_train<-to_categorical(PCNL_multiple_output_train_outcomes$Stay1)

Clavien_coded4_train<-to_categorical(PCNL_multiple_output_train_outcomes$Clavien2)

Immediate_clearance_coded4_test<-as.matrix(PCNL_multiple_output_test_outcomes$Immediate_clearance_coded3)

Visc_injury_coded4_test<-as.matrix(PCNL_multiple_output_test_outcomes$Visc_injury_coded3)

Survival_coded4_test<-as.matrix(PCNL_multiple_output_test_outcomes$Survival_coded3)

Transfusion_coded4_test<-as.matrix(PCNL_multiple_output_test_outcomes$Transfusion_coded3)

Post_infection_coded4_test<-as.matrix(PCNL_multiple_output_test_outcomes$Post_infection_coded3)

Intra_complications_coded4_test<-as.matrix(PCNL_multiple_output_test_outcomes$Intra_complications_coded3)

ITU_HDU_coded4_test<-as.matrix(PCNL_multiple_output_test_outcomes$ITU_HDU_coded3)

SF_coded4_test<-as.matrix(PCNL_multiple_output_test_outcomes$SF_coded3)

Adjuvant_coded4_test<-as.matrix(PCNL_multiple_output_test_outcomes$Adjuvant_coded3)

Post_stay4_test<-to_categorical(PCNL_multiple_output_test_outcomes$Stay1)

Clavien_coded4_test<-to_categorical(PCNL_multiple_output_test_outcomes$Clavien2)

**#Build Model**

library(keras)

library(tensorflow)

library(caret)

library(pROC)

PCNL_base_model<-layer_input(shape=c(43)) %>%

layer_dense(units=128, activation = "relu") %>%

layer_dense(units=128, activation = "relu") %>%

layer_dense(units=43, activation = "relu")

Immediate_clearance_output<-PCNL_base_model %>%

layer_dense(units=1, activation="sigmoid")

Visc_injury_output<-PCNL_base_model %>%

layer_dense(units=1, activation="sigmoid")

Transfusion_output<-PCNL_base_model %>%

layer_dense(units=1, activation="sigmoid")

Post_infection_output<-PCNL_base_model %>%

layer_dense(units=1, activation="sigmoid")

Intra_complications_output<-PCNL_base_model %>%

layer_dense(units=1, activation="sigmoid")

ITU_HDU_output<-PCNL_base_model %>%

layer_dense(units=1, activation="sigmoid")

SF_output<-PCNL_base_model %>%

layer_dense(units=1, activation="sigmoid")

Adjuvant_output<-PCNL_base_model %>%

layer_dense(units=1, activation="sigmoid")

Post_stay_output<-PCNL_base_model %>%

layer_dense(units=4, activation="softmax")

Clavien_output<-PCNL_base_model %>%

layer_dense(units=8, activation="softmax")

PCNL_multi_model<-keras_model(PCNL_base_model, list(Immediate_clearance_output,Visc_injury_output, Transfusion_output, Post_infection_output, Intra_complications_output, ITU_HDU_output, SF_output, Adjuvant_output, Post_stay_output, Clavien_output))

PCNL_multi_model %>%

compile(optimizer = "rmsprop", loss = list("binary_crossentropy","binary_crossentropy", "binary_crossentropy","binary_crossentropy", "binary_crossentropy", "binary_crossentropy","binary_crossentropy","binary_crossentropy", "categorical_crossentropy", "categorical_crossentropy"), metrics='accuracy')

PCNL_multi_model %>%

fit(PCNL_multiple_output_train_predictors1, list(Immediate_clearance_coded4_train,Visc_injury_coded4_train,Transfusion_coded4_train,Post_infection_coded4_train,Intra_complications_coded4_train,ITU_HDU_coded4_train,SF_coded4_train, Adjuvant_coded4_train, Post_stay4_train, Clavien_coded4_train), validation_split=0.2, epoch=200, batch_size=32, verbose=1)

PCNL_multi_model_results<-PCNL_multi_model%>% evaluate(

PCNL_multiple_output_test_predictors1, list(Immediate_clearance_coded4_test,Visc_injury_coded4_test,Transfusion_coded4_test,Post_infection_coded4_test,Intra_complications_coded4_test,ITU_HDU_coded4_test,SF_coded4_test, Adjuvant_coded4_test, Post_stay4_test, Clavien_coded4_test), verbose=1

)

42/42 [==============================] - 0s 3ms/step - loss: 4.4350 - dense_7_loss: 0.5507 - dense_8_loss: 0.0676 - dense_9_loss: 0.0630 - dense_10_loss: 0.2618 - dense_11_loss: 0.2133 - dense_12_loss: 0.2323 - dense_13_loss: 0.8262 - dense_14_loss: 0.2037 - dense_15_loss: 1.1553 - dense_16_loss: 0.8612 - dense_7_accuracy: 0.7722 - dense_8_accuracy: 0.9947 - dense_9_accuracy: 0.9811 - dense_10_accuracy: 0.9155 - dense_11_accuracy: 0.9653 - dense_12_accuracy: 0.9495 - dense_13_accuracy: 0.4367 - dense_14_accuracy: 0.9585 - dense_15_accuracy: 0.6380 - dense_16_accuracy: 0.8122

**#Predictions**

PCNL_multi_model_predict<-PCNL_multi_model%>% predict(

PCNL_multiple_output_test_predictors1, batch_size=32, verbose=1)

PCNL_multi_model_predict1<-as.data.frame(PCNL_multi_model_predict)

PCNL_multi_model_predict_Immediate_clearance<-PCNL_multi_model_predict1[,1]

PCNL_multi_model_predict_Visc_injury_coded<-PCNL_multi_model_predict1[,2]

PCNL_multi_model_predict_Transfusion_coded<-PCNL_multi_model_predict1[,3]

PCNL_multi_model_predict_Post_infection_coded<-PCNL_multi_model_predict1[,4]

PCNL_multi_model_predict_Intra_complications_coded<-PCNL_multi_model_predict1[,5]

PCNL_multi_model_predict_ITU_HDU_coded<-PCNL_multi_model_predict1[,6]

PCNL_multi_model_predict_SF_coded<-PCNL_multi_model_predict1[,7]

PCNL_multi_model_predict_Adjuvant_coded<-PCNL_multi_model_predict1[,8]

PCNL_multi_model_predict_Post_stay<-PCNL_multi_model_predict1[,9:12]

PCNL_multi_model_predict_Clavien_coded<-PCNL_multi_model_predict1[,13:20]

Yes<-c(0,0)

PCNL_multi_model_predict_Immediate_clearance1<-round(PCNL_multi_model_predict_Immediate_clearance, digits = 0)

PCNL_multi_model_predict_Visc_injury_coded<-round(PCNL_multi_model_predict_Visc_injury_coded, digits = 0)

PCNL_multi_model_predict_Transfusion_coded1<-round(PCNL_multi_model_predict_Transfusion_coded, digits = 0)

PCNL_multi_model_predict_Post_infection_coded1<-round(PCNL_multi_model_predict_Post_infection_coded, digits = 0)

PCNL_multi_model_predict_Intra_complications_coded1<-round(PCNL_multi_model_predict_Intra_complications_coded, digits = 0)

PCNL_multi_model_predict_ITU_HDU_coded1<-round(PCNL_multi_model_predict_ITU_HDU_coded, digits = 0)

PCNL_multi_model_predict_SF_coded1<-round(PCNL_multi_model_predict_SF_coded, digits = 0)

PCNL_multi_model_predict_Adjuvant_coded1<-round(PCNL_multi_model_predict_Adjuvant_coded, digits = 0)

PCNL_multi_model_predict_Post_stay1<-round(PCNL_multi_model_predict_Post_stay, digits = 0)

PCNL_multi_model_predict_Clavien_coded1<-round(PCNL_multi_model_predict_Clavien_coded, digits = 0)

#Immediate clearance

Multi_Immediate_clearance_tb<-table(Immediate_clearance_coded4_test ,PCNL_multi_model_predict_Immediate_clearance1)

Multi_Immediate_clearance_roc<-roc(Immediate_clearance_coded4_test ,PCNL_multi_model_predict_Immediate_clearance1)

auc(Multi_Immediate_clearance_roc)

confusionMatrix(Multi_Immediate_clearance_tb)

```
Area under the curve: 0.5334
Confusion Matrix and Statistics

                               PCNL_multi_model_predict_Immediate_clearance1
Immediate_clearance_coded4_test   0   1
                              0  26 285
                              1  17 998

               Accuracy : 0.7722
                 95% CI : (0.7487, 0.7946)
    No Information Rate : 0.9676
    P-Value [Acc > NIR] : 1

                  Kappa : 0.0953

 Mcnemar's Test P-Value : <2e-16

            Sensitivity : 0.60465
            Specificity : 0.77786
         Pos Pred Value : 0.08360
         Neg Pred Value : 0.98325
             Prevalence : 0.03243
         Detection Rate : 0.01961
   Detection Prevalence : 0.23454
      Balanced Accuracy : 0.69126

       'Positive' Class : 0
```

#Visceral Injury

Multi_Visc_Injury_tb<-table(Visc_injury_coded4_test ,PCNL_multi_model_predict_Visc_injury_coded)

Multi_Visc_Injury_roc<-roc(Visc_injury_coded4_test ,PCNL_multi_model_predict_Visc_injury_coded)

auc(Multi_Visc_Injury_roc)

Multi_Visc_Injury_tb1<-cbind(Multi_Visc_Injury_tb,Yes)

colnames(Multi_Visc_Injury_tb1)<-c("0","1")

confusionMatrix(Multi_Visc_Injury_tb1)

```
Area under the curve: 0.5
Confusion Matrix and Statistics

     0    1
0 1319    0
1    7    0

               Accuracy : 0.9947
                 95% CI : (0.9892, 0.9979)
    No Information Rate : 1
    P-Value [Acc > NIR] : 1.00000

                  Kappa : 0

 Mcnemar's Test P-Value : 0.02334

            Sensitivity : 0.9947
            Specificity :     NA
         Pos Pred Value :     NA
         Neg Pred Value :     NA
             Prevalence : 1.0000
         Detection Rate : 0.9947
   Detection Prevalence : 0.9947
      Balanced Accuracy :     NA

       'Positive' Class : 0
```

#Transfusion

Multi_Transfusion_tb<-table(Transfusion_coded4_test ,PCNL_multi_model_predict_Transfusion_coded1)

Multi_Transfusion_roc<-roc(Transfusion_coded4_test ,PCNL_multi_model_predict_Transfusion_coded1)

auc(Multi_Transfusion_roc)

confusionMatrix(Multi_Transfusion_tb)

```
Area under the curve: 0.7688
Confusion Matrix and Statistics

                       PCNL_multi_model_predict_Transfusion_coded1
Transfusion_coded4_test    0    1
                      0 1273    1
                      1   24   28

               Accuracy : 0.9811
                 95% CI : (0.9723, 0.9878)
    No Information Rate : 0.9781
    P-Value [Acc > NIR] : 0.2611

                  Kappa : 0.6824

 Mcnemar's Test P-Value : 1.083e-05

            Sensitivity : 0.9815
            Specificity : 0.9655
         Pos Pred Value : 0.9992
         Neg Pred Value : 0.5385
             Prevalence : 0.9781
         Detection Rate : 0.9600
   Detection Prevalence : 0.9608
      Balanced Accuracy : 0.9735

       'Positive' Class : 0
```

#Post-operative Infection

Multi_Post_Infection_tb<-table(Post_infection_coded4_test ,PCNL_multi_model_predict_Post_infection_coded1)

Multi_Post_Infection_roc<-roc(Post_infection_coded4_test ,PCNL_multi_model_predict_Post_infection_coded1)

auc(Multi_Post_Infection_roc)

confusionMatrix(Multi_Post_Infection_tb)

```
Area under the curve: 0.9048
Confusion Matrix and Statistics

                          PCNL_multi_model_predict_Post_infection_coded1
Post_infection_coded4_test   0   1
                         0 938  76
                         1  36 276

               Accuracy : 0.9155
                 95% CI : (0.8993, 0.9299)
    No Information Rate : 0.7345
    P-Value [Acc > NIR] : < 2.2e-16

                  Kappa : 0.7753

 Mcnemar's Test P-Value : 0.0002286

            Sensitivity : 0.9630
            Specificity : 0.7841
         Pos Pred Value : 0.9250
         Neg Pred Value : 0.8846
             Prevalence : 0.7345
         Detection Rate : 0.7074
   Detection Prevalence : 0.7647
      Balanced Accuracy : 0.8736

       'Positive' Class : 0
```

#Intra-operative complications

Multi_Intra_Complications_tb<-table(Intra_complications_coded4_test ,PCNL_multi_model_predict_Intra_complications_coded1)

Multi_Intra_Complications_roc<-roc(Intra_complications_coded4_test ,PCNL_multi_model_predict_Intra_complications_coded1)

auc(Multi_Intra_Complications_roc)

Multi_Intra_Complications_tb1<-cbind(Multi_Intra_Complications_tb,Yes)

colnames(Multi_Intra_Complications_tb1)<-c("0","1")

confusionMatrix(Multi_Intra_Complications_tb1)

```
Area under the curve: 0.5
Confusion Matrix and Statistics

     0    1
0 1280    0
1   46    0

               Accuracy : 0.9653
                 95% CI : (0.954, 0.9745)
    No Information Rate : 1
    P-Value [Acc > NIR] : 1

                  Kappa : 0

 Mcnemar's Test P-Value : 3.247e-11

            Sensitivity : 0.9653
            Specificity :     NA
         Pos Pred Value :     NA
         Neg Pred Value :     NA
             Prevalence : 1.0000
         Detection Rate : 0.9653
   Detection Prevalence : 0.9653
      Balanced Accuracy :     NA

       'Positive' Class : 0
```

#Need for ITU/HDU

Multi_ITU_HDU_tb<-table(ITU_HDU_coded4_test ,PCNL_multi_model_predict_ITU_HDU_coded1)

Multi_ITU_HDU_roc<-roc(ITU_HDU_coded4_test ,PCNL_multi_model_predict_ITU_HDU_coded1)

auc(Multi_ITU_HDU_roc)

Multi_ITU_HDU_tb1<-cbind(Multi_ITU_HDU_tb,Yes)

colnames(Multi_ITU_HDU_tb1)<-c("0","1")

confusionMatrix(Multi_ITU_HDU_tb1)

```
Area under the curve: 0.5
Confusion Matrix and Statistics

     0    1
0 1259    0
1   67    0

               Accuracy : 0.9495
                 95% CI : (0.9363, 0.9606)
    No Information Rate : 1
    P-Value [Acc > NIR] : 1

                  Kappa : 0

 Mcnemar's Test P-Value : 7.433e-16

            Sensitivity : 0.9495
            Specificity :     NA
         Pos Pred Value :     NA
         Neg Pred Value :     NA
             Prevalence : 1.0000
         Detection Rate : 0.9495
   Detection Prevalence : 0.9495
      Balanced Accuracy :     NA

       'Positive' Class : 0
```

#Stone Free status at follow-up

Multi_SF_tb<-table(SF_coded4_test ,PCNL_multi_model_predict_SF_coded1)

Multi_SF_roc<-roc(SF_coded4_test ,PCNL_multi_model_predict_SF_coded1)

auc(Multi_SF_roc)

#Multi_SF_tb1<-cbind(Multi_SF_tb,Yes)

#colnames(Multi_SF_tb1)<-c("0","1")

confusionMatrix(Multi_SF_tb)

```
Area under the curve: 0.599
Confusion Matrix and Statistics

              PCNL_multi_model_predict_SF_coded1
SF_coded4_test   0   1
             0 387 712
             1  35 192

               Accuracy : 0.4367
                 95% CI : (0.4097, 0.4638)
    No Information Rate : 0.6817
    P-Value [Acc > NIR] : 1

                  Kappa : 0.0907

 Mcnemar's Test P-Value : <2e-16

            Sensitivity : 0.9171
            Specificity : 0.2124
         Pos Pred Value : 0.3521
         Neg Pred Value : 0.8458
             Prevalence : 0.3183
         Detection Rate : 0.2919
   Detection Prevalence : 0.8288
      Balanced Accuracy : 0.5647

       'Positive' Class : 0
```

#Need for Adjuvant treatment

Multi_Adjuvant_tb<-table(Adjuvant_coded4_test ,PCNL_multi_model_predict_Adjuvant_coded1)

Multi_Adjuvant_roc<-roc(Adjuvant_coded4_test ,PCNL_multi_model_predict_Adjuvant_coded1)

auc(Multi_Adjuvant_roc)

Multi_Adjuvant_tb1<-cbind(Multi_Adjuvant_tb,Yes)

colnames(Multi_Adjuvant_tb1)<-c("0","1")

confusionMatrix(Multi_Adjuvant_tb1)

```
Area under the curve: 0.5
Confusion Matrix and Statistics

     0    1
0 1271    0
1   55    0

               Accuracy : 0.9585
                 95% CI : (0.9464, 0.9686)
    No Information Rate : 1
    P-Value [Acc > NIR] : 1

                  Kappa : 0

 Mcnemar's Test P-Value : 3.305e-13

            Sensitivity : 0.9585
            Specificity :     NA
         Pos Pred Value :     NA
         Neg Pred Value :     NA
             Prevalence : 1.0000
         Detection Rate : 0.9585
   Detection Prevalence : 0.9585
      Balanced Accuracy :     NA

       'Positive' Class : 0
```

#Post-operative stay (coded)

PCNL_multi_model_predict_Post_stay2<-as.matrix(PCNL_multi_model_predict_Post_stay1)

Multi_Post_op_stay_tb<-table(Post_stay4_test, PCNL_multi_model_predict_Post_stay2)

Multi_Post_op_stay_roc<-roc(Post_stay4_test, PCNL_multi_model_predict_Post_stay2)

auc(Multi_Post_op_stay_roc)

confusionMatrix(Multi_Post_op_stay_tb)

```
Area under the curve: 0.7564
Confusion Matrix and Statistics

               PCNL_multi_model_predict_Post_stay2
Post_stay4_test    0    1
              0 3537  441
              1  499  827

               Accuracy : 0.8228
                 95% CI : (0.8122, 0.833)
    No Information Rate : 0.7609
    P-Value [Acc > NIR] : < 2e-16

                  Kappa : 0.5204

 Mcnemar's Test P-Value : 0.06301

            Sensitivity : 0.8764
            Specificity : 0.6522
         Pos Pred Value : 0.8891
         Neg Pred Value : 0.6237
             Prevalence : 0.7609
         Detection Rate : 0.6669
   Detection Prevalence : 0.7500
      Balanced Accuracy : 0.7643

       'Positive' Class : 0
```

PCNL_multi_model_predict_Post_stay3<-as.data.frame(PCNL_multi_model_predict_Post_stay1)

Post_stay4_test2<-as.data.frame(Post_stay4_test)

colnames(PCNL_multi_model_predict_Post_stay3)<-c("V1","V2","V3","V4")

#individual outcomes for post-op stay

Multi_Post_op_stay_roc1<-roc(Post_stay4_test2$V1, PCNL_multi_model_predict_Post_stay3$V1)

Multi_Post_op_stay_tb1<-table(Post_stay4_test2$V1, PCNL_multi_model_predict_Post_stay3$V1)

auc(Multi_Post_op_stay_roc1)

confusionMatrix(Multi_Post_op_stay_tb1)

```
Area under the curve: 0.5

Confusion Matrix and Statistics

     0    1
0 1305    0
1   21    0

               Accuracy : 0.9842
                 95% CI : (0.9759, 0.9902)
    No Information Rate : 1
    P-Value [Acc > NIR] : 1

                  Kappa : 0

 Mcnemar's Test P-Value : 1.275e-05

            Sensitivity : 0.9842
            Specificity :     NA
         Pos Pred Value :     NA
         Neg Pred Value :     NA
             Prevalence : 1.0000
         Detection Rate : 0.9842
   Detection Prevalence : 0.9842
      Balanced Accuracy :     NA

       'Positive' Class : 0
```

Multi_Post_op_stay_roc2<-roc(Post_stay4_test2$V2, PCNL_multi_model_predict_Post_stay3$V2)

Multi_Post_op_stay_tb2<-table(Post_stay4_test2$V2, PCNL_multi_model_predict_Post_stay3$V2)

auc(Multi_Post_op_stay_roc2)

confusionMatrix(Multi_Post_op_stay_tb2)

```
Area under the curve: 0.661

Confusion Matrix and Statistics


      0   1
  0 804 265
  1  44 213

               Accuracy : 0.767
                 95% CI : (0.7433, 0.7895)
    No Information Rate : 0.6395
    P-Value [Acc > NIR] : < 2.2e-16

                  Kappa : 0.4379

 Mcnemar's Test P-Value : < 2.2e-16

            Sensitivity : 0.9481
            Specificity : 0.4456
         Pos Pred Value : 0.7521
         Neg Pred Value : 0.8288
             Prevalence : 0.6395
         Detection Rate : 0.6063
   Detection Prevalence : 0.8062
      Balanced Accuracy : 0.6969

       'Positive' Class : 0
```

Multi_Post_op_stay_roc3<-roc(Post_stay4_test2$V3, PCNL_multi_model_predict_Post_stay3$V3)

Multi_Post_op_stay_tb3<-table(Post_stay4_test2$V3, PCNL_multi_model_predict_Post_stay3$V3)

auc(Multi_Post_op_stay_roc3)

confusionMatrix(Multi_Post_op_stay_tb3)

```
Area under the curve: 0.5

Confusion Matrix and Statistics


      0   1
  0 638 396
  1 158 134

               Accuracy : 0.5822
                 95% CI : (0.5551, 0.6089)
    No Information Rate : 0.6003
    P-Value [Acc > NIR] : 0.915

                  Kappa : 0.0587

 Mcnemar's Test P-Value : <2e-16

            Sensitivity : 0.8015
            Specificity : 0.2528
         Pos Pred Value : 0.6170
         Neg Pred Value : 0.4589
             Prevalence : 0.6003
         Detection Rate : 0.4811
   Detection Prevalence : 0.7798
      Balanced Accuracy : 0.5272

       'Positive' Class : 0
```

Multi_Post_op_stay_roc4<-roc(Post_stay4_test2$V4, PCNL_multi_model_predict_Post_stay3$V4)

Multi_Post_op_stay_tb4<-table(Post_stay4_test2$V4, PCNL_multi_model_predict_Post_stay3$V4)

auc(Multi_Post_op_stay_roc4)

confusionMatrix(Multi_Post_op_stay_tb4)

```
Area under the curve: 0.6343

Confusion Matrix and Statistics


      0   1
  0 562   8
  1 699  57

               Accuracy : 0.4668
                 95% CI : (0.4397, 0.4941)
    No Information Rate : 0.951
    P-Value [Acc > NIR] : 1

                  Kappa : 0.0534

 Mcnemar's Test P-Value : <2e-16

            Sensitivity : 0.4457
            Specificity : 0.8769
         Pos Pred Value : 0.9860
         Neg Pred Value : 0.0754
             Prevalence : 0.9510
         Detection Rate : 0.4238
   Detection Prevalence : 0.4299
      Balanced Accuracy : 0.6613

       'Positive' Class : 0
```

**#Clavien grade of complication**

PCNL_multi_model_predict_Clavien_coded2<-as.matrix(PCNL_multi_model_predict_Clavien_coded1)

Clavien_coded4_test2<-as.matrix(Clavien_coded4_test)

Multi_Clavien_tb<-table(Clavien_coded4_test2, PCNL_multi_model_predict_Clavien_coded2)

Multi_Clavien_roc<-roc(Clavien_coded4_test2, PCNL_multi_model_predict_Clavien_coded2)

auc(Multi_Clavien_roc)

confusionMatrix(Multi_Clavien_tb)

```
Area under the curve: 0.8916
Confusion Matrix and Statistics

                    PCNL_multi_model_predict_Clavien_coded2
Clavien_coded4_test2    0    1
                   0 9040  242
                   1  253 1073

               Accuracy : 0.9533
                 95% CI : (0.9492, 0.9573)
    No Information Rate : 0.876
    P-Value [Acc > NIR] : <2e-16

                  Kappa : 0.7859

 Mcnemar's Test P-Value : 0.6531

            Sensitivity : 0.9728
            Specificity : 0.8160
         Pos Pred Value : 0.9739
         Neg Pred Value : 0.8092
             Prevalence : 0.8760
         Detection Rate : 0.8522
   Detection Prevalence : 0.8750
      Balanced Accuracy : 0.8944

       'Positive' Class : 0
```

Clavien_coded4_test2<-as.data.frame(Clavien_coded4_test)

PCNL_multi_model_predict_Clavien_coded3<-as.data.frame(PCNL_multi_model_predict_Clavien_coded1)

colnames(PCNL_multi_model_predict_Clavien_coded3)<-c("V1","V2","V3","V4","V5","V6","V7","V8")

Multi_Clavien_roc1<-roc(Clavien_coded4_test2$V1, PCNL_multi_model_predict_Clavien_coded3$V1)

Multi_Clavien_tb1<-table(Clavien_coded4_test2$V1, PCNL_multi_model_predict_Clavien_coded3$V1)

auc(Multi_Clavien_roc1)

confusionMatrix(Multi_Clavien_tb1)

```
Area under the curve: 0.5163

Confusion Matrix and Statistics


       0    1
  0    3  245
  1    2 1076

               Accuracy : 0.8137
                 95% CI : (0.7917, 0.8343)
    No Information Rate : 0.9962
    P-Value [Acc > NIR] : 1

                  Kappa : 0.0164

 Mcnemar's Test P-Value : <2e-16

            Sensitivity : 0.600000
            Specificity : 0.814534
         Pos Pred Value : 0.012097
         Neg Pred Value : 0.998145
             Prevalence : 0.003771
         Detection Rate : 0.002262
   Detection Prevalence : 0.187029
      Balanced Accuracy : 0.707267

       'Positive' Class : 0
```

Multi_Clavien_roc2<-roc(Clavien_coded4_test2$V2, PCNL_multi_model_predict_Clavien_coded3$V2)

Multi_Clavien_tb2<-table(Clavien_coded4_test2$V2, PCNL_multi_model_predict_Clavien_coded3$V2)

auc(Multi_Clavien_roc2)

confusionMatrix(Multi_Clavien_tb2)

```
Area under the curve: 0.5

Confusion Matrix and Statistics

     0    1
0 1226    0
1  100    0

               Accuracy : 0.9246
                 95% CI : (0.909, 0.9382)
    No Information Rate : 1
    P-Value [Acc > NIR] : 1

                  Kappa : 0

 Mcnemar's Test P-Value : <2e-16

            Sensitivity : 0.9246
            Specificity :     NA
         Pos Pred Value :     NA
         Neg Pred Value :     NA
             Prevalence : 1.0000
         Detection Rate : 0.9246
   Detection Prevalence : 0.9246
      Balanced Accuracy :     NA

       'Positive' Class : 0
```

Multi_Clavien_roc3<-roc(Clavien_coded4_test2$V3, PCNL_multi_model_predict_Clavien_coded3$V3)

Multi_Clavien_tb3<-table(Clavien_coded4_test2$V3, PCNL_multi_model_predict_Clavien_coded3$V3)

auc(Multi_Clavien_roc3)

confusionMatrix(Multi_Clavien_tb3)

```
Area under the curve: 0.5

Confusion Matrix and Statistics

     0    1
0 1229    0
1   97    0

               Accuracy : 0.9268
                 95% CI : (0.9115, 0.9403)
    No Information Rate : 1
    P-Value [Acc > NIR] : 1

                  Kappa : 0

 Mcnemar's Test P-Value : <2e-16

            Sensitivity : 0.9268
            Specificity :     NA
         Pos Pred Value :     NA
         Neg Pred Value :     NA
             Prevalence : 1.0000
         Detection Rate : 0.9268
   Detection Prevalence : 0.9268
      Balanced Accuracy :     NA

       'Positive' Class : 0
```

Multi_Clavien_roc4<-roc(Clavien_coded4_test2$V4, PCNL_multi_model_predict_Clavien_coded3$V4)

Multi_Clavien_tb4<-table(Clavien_coded4_test2$V4, PCNL_multi_model_predict_Clavien_coded3$V4)

auc(Multi_Clavien_roc4)

confusionMatrix(Multi_Clavien_tb4)

```
Area under the curve: 0.4977

Confusion Matrix and Statistics


       0    1
  0 1299    1
  1   26    0

               Accuracy : 0.9796
                 95% CI : (0.9705, 0.9865)
    No Information Rate : 0.9992
    P-Value [Acc > NIR] : 1

                  Kappa : -0.0015

 Mcnemar's Test P-Value : 3.86e-06

            Sensitivity : 0.9804
            Specificity : 0.0000
         Pos Pred Value : 0.9992
         Neg Pred Value : 0.0000
             Prevalence : 0.9992
         Detection Rate : 0.9796
   Detection Prevalence : 0.9804
      Balanced Accuracy : 0.4902

       'Positive' Class : 0
```

Multi_Clavien_roc5<-roc(Clavien_coded4_test2$V5, PCNL_multi_model_predict_Clavien_coded3$V5)

Multi_Clavien_tb5<-table(Clavien_coded4_test2$V5, PCNL_multi_model_predict_Clavien_coded3$V5)

confusionMatrix(Multi_Clavien_tb5)

auc(Multi_Clavien_roc5)

```
Area under the curve: 0.5

Confusion Matrix and Statistics

     0    1
0 1306    0
1   20    0

               Accuracy : 0.9849
                 95% CI : (0.9768, 0.9908)
    No Information Rate : 1
    P-Value [Acc > NIR] : 1

                  Kappa : 0

 Mcnemar's Test P-Value : 2.152e-05

            Sensitivity : 0.9849
            Specificity :     NA
         Pos Pred Value :     NA
         Neg Pred Value :     NA
             Prevalence : 1.0000
         Detection Rate : 0.9849
   Detection Prevalence : 0.9849
      Balanced Accuracy :     NA

       'Positive' Class : 0
```

Multi_Clavien_roc6<-roc(Clavien_coded4_test2$V6, PCNL_multi_model_predict_Clavien_coded3$V6)

Multi_Clavien_tb6<-table(Clavien_coded4_test2$V6, PCNL_multi_model_predict_Clavien_coded3$V6)

auc(Multi_Clavien_roc6)

confusionMatrix(Multi_Clavien_tb6)

```
Area under the curve: 0.5

Confusion Matrix and Statistics

     0    1
0 1323    0
1    3    0

               Accuracy : 0.9977
                 95% CI : (0.9934, 0.9995)
    No Information Rate : 1
    P-Value [Acc > NIR] : 1.0000

                  Kappa : 0

 Mcnemar's Test P-Value : 0.2482

            Sensitivity : 0.9977
            Specificity :     NA
         Pos Pred Value :     NA
         Neg Pred Value :     NA
             Prevalence : 1.0000
         Detection Rate : 0.9977
   Detection Prevalence : 0.9977
      Balanced Accuracy :     NA

       'Positive' Class : 0
```

Multi_Clavien_roc7<-roc(Clavien_coded4_test2$V7, PCNL_multi_model_predict_Clavien_coded3$V7)

auc(Multi_Clavien_roc7)

confusionMatrix(Multi_Clavien_tb7)

```
Area under the curve: 0.5

Confusion Matrix and Statistics

     0    1
0 1325    0
1    1    0

               Accuracy : 0.9992
                 95% CI : (0.9958, 1)
    No Information Rate : 1
    P-Value [Acc > NIR] : 1

                  Kappa : 0

 Mcnemar's Test P-Value : 1

            Sensitivity : 0.9992
            Specificity :     NA
         Pos Pred Value :     NA
         Neg Pred Value :     NA
             Prevalence : 1.0000
         Detection Rate : 0.9992
   Detection Prevalence : 0.9992
      Balanced Accuracy :     NA

       'Positive' Class : 0
```

Multi_Clavien_roc8<-roc(Clavien_coded4_test2$V8, PCNL_multi_model_predict_Clavien_coded3$V8)

Multi_Clavien_tb8<-table(Clavien_coded4_test2$V8, PCNL_multi_model_predict_Clavien_coded3$V8)

auc(Multi_Clavien_roc8)

confusionMatrix(Multi_Clavien_tb8)

```
Area under the curve: 0.4992

Confusion Matrix and Statistics

     0    1
0 1325    0
1    1    0

               Accuracy : 0.9992
                 95% CI : (0.9958, 1)
    No Information Rate : 1
    P-Value [Acc > NIR] : 1

                  Kappa : 0

 Mcnemar's Test P-Value : 1

            Sensitivity : 0.9992
            Specificity :     NA
         Pos Pred Value :     NA
         Neg Pred Value :     NA
             Prevalence : 1.0000
         Detection Rate : 0.9992
   Detection Prevalence : 0.9992
      Balanced Accuracy :     NA

       'Positive' Class : 0
```

4. **Immediate Clearance on Fluoroscopy in Theatre**

#Create Training/Test sets

PCNL_immediate_clearance1<-na.omit(PCNL_immediate_clearance_coded)

PCNL_immediate_clearance2<-na.omit(PCNL_immediate_clearance_non_coded)

set.seed(1234)

PCNL_immediate_clearance_sample_coded<-sample(1:nrow(PCNL_immediate_clearance1), size=nrow(PCNL_immediate_clearance1) *0.7)

PCNL_immediate_clearance_train1<-PCNL_immediate_clearance1[PCNL_immediate_clearance_sample_coded,]

PCNL_immediate_clearance_test1<-PCNL_immediate_clearance1[-PCNL_immediate_clearance_sample_coded,]

PCNL_immediate_clearance_train_predictors1<-PCNL_immediate_clearance_train[,-44]

PCNL_immediate_clearance_test_predictors1<-PCNL_immediate_clearance_test[,-44]

PCNL_immediate_clearance_sample_non_coded<- sample(1:nrow(PCNL_immediate_clearance2), size=nrow(PCNL_immediate_clearance2) *0.7)

PCNL_immediate_clearance_train2<-PCNL_immediate_clearance2[PCNL_immediate_clearance_sample_non_coded,]

PCNL_immediate_clearance_test2<-PCNL_immediate_clearance2[-PCNL_immediate_clearance_sample_non_coded,]

PCNL_immediate_clearance_train_predictors2<-PCNL_immediate_clearance_train2[,-44]

PCNL_immediate_clearance_test_predictors2<-PCNL_immediate_clearance_test2[,-44]

#coded outcomes

PCNL_immediate_clearance3<-na.omit(PCNL_immediate_clearance_coded2)

set.seed(1234)

PCNL_immediate_clearance_sample_coded3<-sample(1:nrow(PCNL_immediate_clearance3), size=nrow(PCNL_immediate_clearance3) *0.7)

PCNL_immediate_clearance_train3<-PCNL_immediate_clearance3[PCNL_immediate_clearance_sample_coded3,]

PCNL_immediate_clearance_test3<-PCNL_immediate_clearance3[-PCNL_immediate_clearance_sample_coded3,]

PCNL_immediate_clearance_train_predictors3<-PCNL_immediate_clearance_train3[,-44]

PCNL_immediate_clearance_test_predictors3<-PCNL_immediate_clearance_test3[,-44]

PCNL_immediate_clearance_train_outcome3<- PCNL_immediate_clearance_train3[,-1:-43]

PCNL_immediate_clearance_test_outcome3<- PCNL_immediate_clearance_test3[,-1:-43]

PCNL_immediate_clearance_ctrl<-trainControl(method= "repeatedcv", number=10, classProbs=TRUE, savePredictions=TRUE)

#make matrices

PCNL_immediate_clearance_train_predictors4<-as.matrix(PCNL_immediate_clearance_train_predictors3)

PCNL_immediate_clearance_train_outcome4<-as.matrix(PCNL_immediate_clearance_train_outcome3)

PCNL_immediate_clearance_test_predictors4<-as.matrix(PCNL_immediate_clearance_test_predictors3)

PCNL_immediate_clearance_test_outcome4<-as.matrix(PCNL_immediate_clearance_test_outcome3)

**#Create Training/Test sets**

PCNL_immediate_clearance1<-na.omit(PCNL_immediate_clearance_coded)

PCNL_immediate_clearance2<-na.omit(PCNL_immediate_clearance_non_coded)

set.seed(1234)

PCNL_immediate_clearance_sample_coded<-sample(1:nrow(PCNL_immediate_clearance1), size=nrow(PCNL_immediate_clearance1) *0.7)

PCNL_immediate_clearance_train1<-PCNL_immediate_clearance1[PCNL_immediate_clearance_sample_coded,]

PCNL_immediate_clearance_test1<-PCNL_immediate_clearance1[-PCNL_immediate_clearance_sample_coded,]

PCNL_immediate_clearance_train_predictors1<-PCNL_immediate_clearance_train[,-44]

PCNL_immediate_clearance_test_predictors1<-PCNL_immediate_clearance_test[,-44]

PCNL_immediate_clearance_sample_non_coded<- sample(1:nrow(PCNL_immediate_clearance2), size=nrow(PCNL_immediate_clearance2) *0.7)

PCNL_immediate_clearance_train2<-PCNL_immediate_clearance2[PCNL_immediate_clearance_sample_non_coded,]

PCNL_immediate_clearance_test2<-PCNL_immediate_clearance2[-PCNL_immediate_clearance_sample_non_coded,]

PCNL_immediate_clearance_train_predictors2<-PCNL_immediate_clearance_train2[,-44]

PCNL_immediate_clearance_test_predictors2<-PCNL_immediate_clearance_test2[,-44]

**#coded outcomes**

PCNL_immediate_clearance3<-na.omit(PCNL_immediate_clearance_coded2)

set.seed(1234)

PCNL_immediate_clearance_sample_coded3<-sample(1:nrow(PCNL_immediate_clearance3), size=nrow(PCNL_immediate_clearance3) *0.7)

PCNL_immediate_clearance_train3<-PCNL_immediate_clearance3[PCNL_immediate_clearance_sample_coded3,]

PCNL_immediate_clearance_test3<-PCNL_immediate_clearance3[-PCNL_immediate_clearance_sample_coded3,]

PCNL_immediate_clearance_train_predictors3<-PCNL_immediate_clearance_train3[,-44]

PCNL_immediate_clearance_test_predictors3<-PCNL_immediate_clearance_test3[,-44]

PCNL_immediate_clearance_train_outcome3<- PCNL_immediate_clearance_train3[,-1:-43]

PCNL_immediate_clearance_test_outcome3<- PCNL_immediate_clearance_test3[,-1:-43]

**PCNL_immediate_clearance**_ctrl<-trainControl(method= "repeatedcv", number=10, classProbs=TRUE, savePredictions=TRUE)

**Random forests**

immediate_clearance_rf<-randomForest(PCNL_immediate_clearance_train_predictors2, PCNL_immediate_clearance_train2$Immediate_clearance)

immediate_clearance_rf_predict<-predict(immediate_clearance_rf, newdata=PCNL_immediate_clearance_test_predictors2)

immediate_clearance_rf_tb<-table(immediate_clearance_rf_predict, ref=PCNL_immediate_clearance_test2$Immediate_clearance)

confusionMatrix(immediate_clearance_rf_tb)

Confusion Matrix and Statistics

ref

PCNL_immediate_clearance_pred No Yes

No	67 47

Yes 244 965

Accuracy : 0.78

95% CI : (0.7567, 0.8021)

No Information Rate : 0.7649

P-Value [Acc > NIR] : 0.1024

Kappa : 0.2165

Mcnemar's Test P-Value : <2e-16

Sensitivity : 0.21543

Specificity : 0.95356

Pos Pred Value : 0.58772

Neg Pred Value : 0.79818

Prevalence : 0.23507

Detection Rate : 0.05064

Detection Prevalence : 0.08617

Balanced Accuracy : 0.58450

'Positive' Class : No

immediate_clearance_rf2<-train(Immediate_clearance~., data=PCNL_immediate_clearance_train1, method="cforest")

immediate_clearance_rf2_predict<-predict(immediate_clearance_rf2, newdata=PCNL_immediate_clearance_test_predictors1)

immediate_clearance_rf2_tb<-table(immediate_clearance_rf2_predict, ref=PCNL_immediate_clearance_test1$Immediate_clearance)

confusionMatrix(immediate_clearance_rf2_tb)

immediate_clearance_rf3<-train(Immediate_clearance~., data=PCNL_immediate_clearance_train1, method="rf", trControl=PCNL_immediate_clearance_ctrl)

immediate_clearance_rf3_predict<-predict(immediate_clearance_rf3, newdata=PCNL_immediate_clearance_test_predictors1)

immediate_clearance_rf3_tb<-table(immediate_clearance_rf3_predict, ref=PCNL_immediate_clearance_test1$Immediate_clearance)

confusionMatrix(immediate_clearance_rf3_tb)

res_immediate_clearance_rf3<-evalm(immediate_clearance_rf3)

***MLeval: Machine Learning Model Evaluation***

Input: caret train function object

Averaging probs.

Group 1 type: repeatedcv

Observations: 3092

Number of groups: 1

Observations per group: 3092

Positive: Yes

Negative: No

Group: Group 1

Positive: 2360

Negative: 732

***Performance Metrics***

Group 1 Optimal Informedness = 0.347068630175049

Group 1 AUC-ROC = 0.73

Confusion Matrix and Statistics

ref

immediate_clearance_rf3_predict No Yes

No	88 64

Yes 244 930

Accuracy : 0.7677

95% CI : (0.744, 0.7902)

No Information Rate : 0.7496

P-Value [Acc > NIR] : 0.06728

Kappa : 0.2449

Mcnemar's Test P-Value : < 2e-16

Sensitivity : 0.26506

Specificity : 0.93561

Pos Pred Value : 0.57895

Neg Pred Value : 0.79216

Prevalence : 0.25038

Detection Rate : 0.06637

Detection Prevalence : 0.11463

Balanced Accuracy : 0.60034

'Positive' Class : No

**Partitioning**

immediate_clearance_rpart<-rpart(Immediate_clearance ~ ., data=PCNL_immediate_clearance_train2, method="class")

immediate_clearance_rpart_predict<-predict(immediate_clearance_rpart, newdata=PCNL_immediate_clearance_test2, type="class")

immediate_clearance_rpart_tb<-table(pred=factor(immediate_clearance_rpart_predict), ref=PCNL_immediate_clearance_test2$Immediate_clearance)

confusionMatrix(immediate_clearance_rpart_tb)

Confusion Matrix and Statistics

ref

pred	No Yes

No	70 69

Yes 246 938

Accuracy : 0.7619

95% CI : (0.738, 0.7846)

No Information Rate : 0.7611

P-Value [Acc > NIR] : 0.4894

Kappa : 0.1894

Mcnemar's Test P-Value : <2e-16

Sensitivity : 0.22152

Specificity : 0.93148

Pos Pred Value : 0.50360

Neg Pred Value : 0.79223

Prevalence : 0.23885

Detection Rate : 0.05291

Detection Prevalence : 0.10506

Balanced Accuracy : 0.57650

'Positive' Class : No

immediate_clearance_rpart<-train(Immediate_clearance ~ ., data=PCNL_immediate_clearance_train1, method="rpart", trControl= **PCNL_immediate_clearance**_ctrl)

immediate_clearance_rpart_predict<-predict(immediate_clearance_rpart, newdata=PCNL_immediate_clearance_test1)

immediate_clearance_rpart_tb<-table(pred=factor(immediate_clearance_rpart_predict), ref=PCNL_immediate_clearance_test1$Immediate_clearance)

confusionMatrix(immediate_clearance_rpart_tb)

res_immediate_clearance_rpart<- evalm(immediate_clearance_rpart)

***MLeval: Machine Learning Model Evaluation***

Input: caret train function object

Averaging probs.

Group 1 type: repeatedcv

Observations: 3092

Number of groups: 1

Observations per group: 3092

Positive: Yes

Negative: No

Group: Group 1

Positive: 2360

Negative: 732

***Performance Metrics***

Group 1 Optimal Informedness = 0.235021302213578

Group 1 AUC-ROC = 0.63

Confusion Matrix and Statistics

ref

pred	No Yes

No	77 104

Yes 279 866

Accuracy : 0.7112

95% CI : (0.6859, 0.7354)

No Information Rate : 0.7315

P-Value [Acc > NIR] : 0.955

Kappa : 0.1292

Mcnemar's Test P-Value : <2e-16

Sensitivity : 0.21629

Specificity : 0.89278

Pos Pred Value : 0.42541

Neg Pred Value : 0.75633

Prevalence : 0.26848

Detection Rate : 0.05807

Detection Prevalence : 0.13650

Balanced Accuracy : 0.55454

'Positive' Class : No

**XGBoost**

#Convert data.frame to data.matrix

PCNL_immediate_clearance_train_matrix<-as.matrix.data.frame(PCNL_immediate_clearance_train2)

PCNL_immediate_clearance_test_matrix<-as.matrix.data.frame(PCNL_immediate_clearance_test2)

**#new 24/10/21**

immediate_clearance_logit<-train(Immediate_clearance_coded2~., data=PCNL_immediate_clearance_train3, method="LogitBoost", trControl= PCNL_immediate_clearance_ctrl)

immediate_clearance_logit_predict<-predict(immediate_clearance_logit, PCNL_immediate_clearance_test3)

immediate_clearance_logit_tb<-table(pred=factor(immediate_clearance_logit_predict), ref=PCNL_immediate_clearance_test3$Immediate_clearance2)

confusionMatrix(immediate_clearance_logit_tb)

res_immediate_clearance_logit<- evalm(data.frame(immediate_clearance_logit_predict, PCNL_immediate_clearance_test_outcome3))

***MLeval: Machine Learning Model Evaluation***

Input: caret train function object

Averaging probs.

Group 1 type: repeatedcv

Observations: 3092

Number of groups: 1

Observations per group: 3092

Positive: Yes

Negative: No

Group: Group 1

Positive: 2360

Negative: 732

***Performance Metrics***

Group 1 Optimal Informedness = 0.38935583958507

Group 1 AUC-ROC = 0.75

Confusion Matrix and Statistics

ref

pred	No Yes

No	89 57

Yes 243 937

Accuracy : 0.7738

95% CI : (0.7503, 0.796)

No Information Rate : 0.7496

P-Value [Acc > NIR] : 0.02204

Kappa : 0.2591

Mcnemar's Test P-Value : < 2e-16

Sensitivity : 0.26807

Specificity : 0.94266

Pos Pred Value : 0.60959

Neg Pred Value : 0.79407

Prevalence : 0.25038

Detection Rate : 0.06712

Detection Prevalence : 0.11011

Balanced Accuracy : 0.60536

'Positive' Class : No

**Logistic Regression**

immediate_clearance_logit<-train(Immediate_clearance~., data=PCNL_immediate_clearance_train1, method="LogitBoost", trControl= **PCNL_immediate_clearance**_ctrl)

immediate_clearance_logit_predict<-predict(immediate_clearance_logit, PCNL_immediate_clearance_test1)

immediate_clearance_logit_tb<-table(pred=factor(immediate_clearance_logit_predict), ref=PCNL_immediate_clearance_test1$Immediate_clearance)

confusionMatrix(immediate_clearance_logit_tb)

res_**immediate_clearance**_logit<- evalm(**immediate_clearance**_logit)

***MLeval: Machine Learning Model Evaluation***

Input: caret train function object

Averaging probs.

Group 1 type: repeatedcv

Observations: 3092

Number of groups: 1

Observations per group: 3092

Positive: Yes

Negative: No

Group: Group 1

Positive: 2360

Negative: 732

***Performance Metrics***

Group 1 Optimal Informedness = 0.197295545058813

Group 1 AUC-ROC = 0.63

Confusion Matrix and Statistics

ref

pred	No Yes

No	40 32

Yes 292 962

Accuracy : 0.7557

95% CI : (0.7316, 0.7786)

No Information Rate : 0.7496

P-Value [Acc > NIR] : 0.3187

Kappa : 0.1194

Mcnemar's Test P-Value : <2e-16

Sensitivity : 0.12048

Specificity : 0.96781

Pos Pred Value : 0.55556

Neg Pred Value : 0.76715

Prevalence : 0.25038

Detection Rate : 0.03017

Detection Prevalence : 0.05430

Balanced Accuracy : 0.54414

'Positive' Class : No

**NeuralNet**

**Recode entire datasheet to use integers not characters – 27/9/21**

**#need to convert factors into dummy integer variables using “model.matrix”**

immediate_clearance_nn_data<-model.matrix(Immediate_clearance ~ ., data=PCNL_immediate_clearance_train)

immediate_clearance_nn<-neuralnet(Immediate_clearance ~ Age + Gender_coded + Year + Side_coded + BMI + CharlsonScore + AgeRelatedCharlsonScore + Previous_UTI_coded + Pre_Op_abx_coded + Pre_Op_MSU_coded + Pre_Op_MSU_result_coded+ Pre_op_rad_prim_coded + Pre_op_radiology_secondary_coded + DMSA_coded + Catheter_coded + PreOpHaemoglobin + GFR_coded + Induction_Abx_coded + Surgeon_Grade_coded + Anaesthesia_coded + IR_Backup_coded + Size_coded + Number_stones_coded + Index_stone_coded + Other_location_coded + GSS + HU_coded + Pre_existing_PCN_coded + Tract_performer_coded + Tract_performer_grade_coded + Puncture_site_coded + Image_guidance_puncture_coded + PatientPosition_coded + NumberOfTractsPlanned + Trace_placement_coded + Amplatz_size_coded + Dilators_coded + Predicted_difficulty_coded + Accessory_procedures_coded + PCN_post_procedure_coded + Primary_technique_coded + Secondary_technique_coded, data=PCNL_immediate_clearance_train1, linear.output = FALSE)

##New 22/10/21

immediate_clearance_nn<-neuralnet(Immediate_clearance2 ~ ., data=PCNL_immediate_clearance_train3, linear.output = FALSE, err.fct= ‘ce’, likelihood=TRUE)

immediate_clearance_nn_predict<-predict(immediate_clearance_nn, newdata = PCNL_immediate_clearance_test3)

immediate_clearance_nn_predict<-compute(immediate_clearance_nn, PCNL_immediate_clearance_test3)

immediate_clearance_nn_tb<-data.frame(actual=PCNL_immediate_clearance_test3$Immediate_clearance2, prediction=immediate_clearance_nn_predict)

confusionMatrix(immediate_clearance_nn_tb)

**#New 24/10/21**

immediate_clearance_nn2<-train(Immediate_clearance~., data=PCNL_immediate_clearance_train1, method="nnet", trControl= **PCNL_immediate_clearance**_ctrl)

immediate_clearance_nn2_predict<-predict(immediate_clearance_nn2, PCNL_immediate_clearance_test1)

immediate_clearance_nn2_tb<-table(pred=factor(immediate_clearance_nn2_predict), ref=PCNL_immediate_clearance_test1$Immediate_clearance)

confusionMatrix(immediate_clearance_nn2_tb)

res_**immediate_clearance**_nn2<- evalm(**immediate_clearance**_nn2)

***MLeval: Machine Learning Model Evaluation***

Input: caret train function object

Averaging probs.

Group 1 type: repeatedcv

Observations: 3092

Number of groups: 1

Observations per group: 3092

Positive: Yes

Negative: No

Group: Group 1

Positive: 2360

Negative: 732

***Performance Metrics***

Group 1 Optimal Informedness = 0.359667037139946

Group 1 AUC-ROC = 0.74

Confusion Matrix and Statistics

ref

pred	No Yes

No	77 70

Yes 255 924

Accuracy : 0.7549

95% CI : (0.7308, 0.7778)

No Information Rate : 0.7496

P-Value [Acc > NIR] : 0.3418

Kappa : 0.1983

Mcnemar's Test P-Value : <2e-16

Sensitivity : 0.23193

Specificity : 0.92958

Pos Pred Value : 0.52381

Neg Pred Value : 0.78372

Prevalence : 0.25038

Detection Rate : 0.05807

Detection Prevalence : 0.11086

Balanced Accuracy : 0.58075

'Positive' Class : No

**Bayesian Generalised Linear Model**

Package: “arm”

immediate_clearance_bayes<-train(Immediate_clearance ~., data= PCNL_immediate_clearance_train1, method="bayesglm", trControl= **PCNL_immediate_clearance**_ctrl)

immediate_clearance_bayes_predict<-predict(immediate_clearance_bayes, PCNL_immediate_clearance_test1, type=)

immediate_clearance_bayes_tb<-table(pred=factor(immediate_clearance_bayes_predict), ref= PCNL_immediate_clearance_test1$Immediate_clearance)

confusionMatrix(immediate_clearance_bayes_tb)

res_**immediate_clearance**_bayes<- evalm(**immediate_clearance**_bayes)

***MLeval: Machine Learning Model Evaluation***

Input: caret train function object

Averaging probs.

Group 1 type: repeatedcv

Observations: 3092

Number of groups: 1

Observations per group: 3092

Positive: Yes

Negative: No

Group: Group 1

Positive: 2360

Negative: 732

***Performance Metrics***

Group 1 Optimal Informedness = 0.374986107252014

Group 1 AUC-ROC = 0.75

Confusion Matrix and Statistics

ref

pred	No Yes

No	96 64

Yes 236 930

Accuracy : 0.7738

95% CI : (0.7503, 0.796)

No Information Rate : 0.7496

P-Value [Acc > NIR] : 0.02204

Kappa : 0.2716

Mcnemar's Test P-Value : < 2e-16

Sensitivity : 0.2892

Specificity : 0.9356

Pos Pred Value : 0.6000

Neg Pred Value : 0.7976

Prevalence : 0.2504

Detection Rate : 0.0724

Detection Prevalence : 0.1207

Balanced Accuracy : 0.6124

'Positive' Class : No

**Generate ROC curves**

res_immediate_clearance_rf3<-evalm(data.frame(immediate_clearance_rf3_predict, PCNL_immediate_clearance_test1$Immediate_clearance))

res_immediate_clearance_rpart<-evalm(data.frame(immediate_clearance_rpart_predict, PCNL_immediate_clearance_test1$Immediate_clearance))

res_immediate_clearance_xgboost<-evalm(data.frame(immediate_clearance_xgboost_predict, PCNL_immediate_clearance_test1$Immediate_clearance))

res_immediate_clearance_logit<-evalm(data.frame(immediate_clearance_logit_predict, PCNL_immediate_clearance_test1$Immediate_clearance))

res_immediate_clearance_nn2<-evalm(data.frame(immediate_clearance_nn2_predict, PCNL_immediate_clearance_test1$Immediate_clearance))

res_immediate_clearance_bayes<-evalm(data.frame(immediate_clearance_bayes_predict, PCNL_immediate_clearance_test1$Immediate_clearance))

res_immediate_clearance_keras<-evalm(data.frame(immediate_clearance_keras_prediction1, PCNL_immediate_clearance_test_outcome3))

res_**immediate_clearance**_rf3$roc

res_**immediate_clearance**_rpart$roc

res_**immediate_clearance**_xgboost$roc

res_**immediate_clearance**_logit$roc

res_**immediate_clearance**_nn2$roc

res_**immediate_clearance**_bayes$roc

#Combine ROC curves

res_immediate_clearance<-evalm(list(immediate_clearance_rf3, **immediate_clearance**_rpart, **immediate_clearance**_xgboost, **immediate_clearance**_logit, **immediate_clearance**_nn2, **immediate_clearance**_bayes, immediate_clearance_keras), gnames=c('Random Forest', 'Partitioning', 'Extreme Gradient Boosting', 'Logistic Regression', 'Neural Network', 'Bayesian Generalised Linear Model', 'Convolutional Neural Network'))

#Deep Neural Net

immediate_clearance_keras<-keras_model_sequential()

immediate_clearance_keras %>% layer_dense(units = 32, input_shape = c(43)) %>% layer_activation('relu') %>% layer_dense(units = 1) %>% layer_activation('sigmoid')

immediate_clearance_keras %>% compile(loss='binary_crossentropy', optimizer='rmsprop', metrics='accuracy')

immediate_clearance_keras %>% fit(PCNL_immediate_clearance_train_predictors4, PCNL_immediate_clearance_train_outcome3, epochs=200, batch_size=5, validation_split=0.2, verbose=1)

immediate_clearance_keras_prediction<-immediate_clearance_keras%>%predict(PCNL_immediate_clearance_test_predictors4, batch_size=32, verbose=1)

immediate_clearance_keras_prediction1<-round(immediate_clearance_keras_prediction, digits=0)

immediate_clearance_keras_tb<- table(PCNL_immediate_clearance_test_outcome4, immediate_clearance_keras_prediction1)

immediate_clearance_keras_roc<- roc(PCNL_immediate_clearance_test_outcome4, immediate_clearance_keras_prediction1)

auc(immediate_clearance_keras_roc)

Area under the curve: 0.5867

plot(immediate_clearance_keras_roc)

confusionMatrix(immediate_clearance_keras_tb)

Confusion Matrix and Statistics

immediate_clearance_keras_prediction1

PCNL_immediate_clearance_test_outcome4	0	1

0 73 247

1 55 951

Accuracy : 0.7722

95% CI : (0.7487, 0.7946)

No Information Rate : 0.9035

P-Value [Acc > NIR] : 1

Kappa : 0.2181

Mcnemar's Test P-Value : <2e-16

Sensitivity : 0.57031

Specificity : 0.79382

Pos Pred Value : 0.22813

Neg Pred Value : 0.94533

Prevalence : 0.09653

Detection Rate : 0.05505

Detection Prevalence : 0.24133

Balanced Accuracy : 0.68207

'Positive' Class : 0

5. Visceral Injury

#Create training/test sets:

set.seed(1234)

PCNL_visc_injury1<-na.omit(PCNL_Visc_injury_coded)

PCNL_visc_injury2<-na.omit(PCNL_Visc_injury_non_coded)

set.seed(1234)

PCNL_visc_injury_sample_coded<-sample(1:nrow(PCNL_visc_injury1), size=nrow(PCNL_visc_injury1) *0.7)

PCNL_visc_injury_train1<-PCNL_visc_injury1[PCNL_visc_injury_sample_coded,]

PCNL_visc_injury_test1<-PCNL_visc_injury1[-PCNL_visc_injury_sample_coded,]

PCNL_visc_injury_train_predictors1<-PCNL_visc_injury_train1[,-44]

PCNL_visc_injury_test_predictors1<-PCNL_visc_injury_test1[,-44]

PCNL_visc_injury_sample_non_coded<- sample(1:nrow(PCNL_visc_injury2), size=nrow(PCNL_immediate_clearance2) *0.7)

PCNL_visc_injury_train2<-PCNL_visc_injury2[PCNL_visc_injury_sample_non_coded,]

PCNL_visc_injury_test2<-PCNL_visc_injury2[-PCNL_visc_injury_sample_non_coded,]

PCNL_visc_injury_train_predictors2<-PCNL_visc_injury_train2[,-44]

PCNL_visc_injury_test_predictors2<-PCNL_visc_injury_test2[,-44]

PCNL_visc_injury_ctrl<-trainControl(method= "repeatedcv", number=10, classProbs=TRUE, savePredictions=TRUE)

#coded outcomes

PCNL_visc_injury3<-na.omit(PCNL_Visc_injury_coded2)

set.seed(1234)

PCNL_visc_injury_sample_coded3<-sample(1:nrow(PCNL_visc_injury3), size=nrow(PCNL_visc_injury3) *0.7)

PCNL_visc_injury_train3<-PCNL_visc_injury3[PCNL_visc_injury_sample_coded3,]

PCNL_visc_injury_test3<- PCNL_visc_injury3 [-PCNL_visc_injury_sample_coded3,]

PCNL_visc_injury_train_predictors3<-PCNL_visc_injury_train3[,-44]

PCNL_visc_injury_test_predictors3<-PCNL_visc_injury_test3[,-44]

PCNL_visc_injury_train_outcome3<- PCNL_visc_injury_train3[,-1:-43]

PCNL_visc_injury_test_outcome3<- PCNL_visc_injury_test3[,-1:-43]

PCNL_visc_injury_ctrl<-trainControl(method= "repeatedcv", number=10, classProbs=TRUE, savePredictions=TRUE)

#make matrices

PCNL_visc_injury_train_predictors4<-as.matrix(PCNL_visc_injury_train_predictors3)

PCNL_visc_injury_train_outcome4<-as.matrix(PCNL_visc_injury_train_outcome3)

PCNL_visc_injury_test_predictors4<-as.matrix(PCNL_visc_injury_test_predictors3)

PCNL_visc_injury_test_outcome4<-as.matrix(PCNL_visc_injury_test_outcome3)

**#Create training/test sets:**

set.seed(1234)

PCNL_visc_injury1<-na.omit(PCNL_Visc_injury_coded)

PCNL_visc_injury2<-na.omit(PCNL_Visc_injury_non_coded)

set.seed(1234)

PCNL_visc_injury_sample_coded<-sample(1:nrow(PCNL_visc_injury1), size=nrow(PCNL_visc_injury1) *0.7)

PCNL_visc_injury_train1<-PCNL_visc_injury1[PCNL_visc_injury_sample_coded,]

PCNL_visc_injury_test1<-PCNL_visc_injury1[-PCNL_visc_injury_sample_coded,]

PCNL_visc_injury_train_predictors1<-PCNL_visc_injury_train1[,-44]

PCNL_visc_injury_test_predictors1<-PCNL_visc_injury_test1[,-44]

PCNL_visc_injury_sample_non_coded<- sample(1:nrow(PCNL_visc_injury2), size=nrow(PCNL_immediate_clearance2) *0.7)

PCNL_visc_injury_train2<-PCNL_visc_injury2[PCNL_visc_injury_sample_non_coded,]

PCNL_visc_injury_test2<-PCNL_visc_injury2[-PCNL_visc_injury_sample_non_coded,]

PCNL_visc_injury_train_predictors2<-PCNL_visc_injury_train2[,-44]

PCNL_visc_injury_test_predictors2<-PCNL_visc_injury_test2[,-44]

**PCNL_visc_injury**_ctrl<-trainControl(method= "repeatedcv", number=10, classProbs=TRUE, savePredictions=TRUE)

#coded outcomes

PCNL_visc_injury3<-na.omit(**PCNL_Visc_injury_coded2**)

set.seed(1234)

PCNL_visc_injury_sample_coded3<-sample(1:nrow(PCNL_visc_injury3), size=nrow(PCNL_visc_injury3) *0.7)

PCNL_visc_injury_train3<-PCNL_visc_injury3[PCNL_visc_injury_sample_coded3,]

PCNL_visc_injury_test3<- PCNL_visc_injury3 [-PCNL_visc_injury_sample_coded3,]

PCNL_visc_injury_train_predictors3<-PCNL_visc_injury_train3[,-44]

PCNL_visc_injury_test_predictors3<-PCNL_visc_injury_test3[,-44]

PCNL_visc_injury_train_outcome3<- PCNL_visc_injury_train3[,-1:-43]

PCNL_visc_injury_test_outcome3<- PCNL_visc_injury_test3[,-1:-43]

PCNL_visc_injury_ctrl<-trainControl(method= "repeatedcv", number=10, classProbs=TRUE, savePredictions=TRUE)

#make matrices

PCNL_visc_injury_train_predictors4<-as.matrix(PCNL_visc_injury_train_predictors3)

PCNL_visc_injury_train_outcome4<-as.matrix(PCNL_visc_injury_train_outcome3)

PCNL_visc_injury_test_predictors4<-as.matrix(PCNL_visc_injury_test_predictors3)

PCNL_visc_injury_test_outcome4<-as.matrix(PCNL_visc_injury_test_outcome3)

**#Random Forests**

**Visc_injury**_rf<- train(**Visc_injury** ~., data=**PCNL_visc_injury_train1**, method="cforest", trControl= **PCNL_visc_injury**_ctrl)

**Visc_injury**_rf_predict<-predict(**Visc_injury**_rf, newdata=**PCNL_visc_injury**_test_predictors1)

**Visc_injury**_rf_tb<-table(**Visc_injury**_rf_predict, ref=**PCNL_visc_injury**_test1$**Visc_injury**)

confusionMatrix(**Visc_injury**_rf_tb)

res_**Visc_injury**_rf<- evalm(**Visc_injury_rf**)

***MLeval: Machine Learning Model Evaluation***

Input: caret train function object

Averaging probs.

Group 1 type: repeatedcv

Observations: 3092

Number of groups: 1

Observations per group: 3092

Positive: Yes

Negative: No

Group: Group 1

Positive: 11

Negative: 3081

***Performance Metrics***

Group 1 Optimal Informedness = 0.444247735386976

Group 1 AUC-ROC = 0.72

Confusion Matrix and Statistics

ref

Visc_injury_rf_predict	No Yes

No 1324	2

Yes	0	0

Accuracy : 0.9985

95% CI : (0.9946, 0.9998)

No Information Rate : 0.9985

P-Value [Acc > NIR] : 0.6767

Kappa : 0

Mcnemar's Test P-Value : 0.4795

Sensitivity : 1.0000

Specificity : 0.0000

Pos Pred Value : 0.9985

Neg Pred Value : NaN

Prevalence : 0.9985

Detection Rate : 0.9985

Detection Prevalence : 1.0000

Balanced Accuracy : 0.5000

'Positive' Class : No

**#Partitioning**

**Visc_injury**_rpart<-train(**Visc_injury** ~ ., data= **PCNL_visc_injury_train1**, method="rpart", trControl= **PCNL_visc_injury**_ctrl)

**Visc_injury**_rpart_predict<-predict(**Visc_injury**_rpart, newdata= **PCNL_visc_injury**_test1)

**Visc_injury**_rpart_tb<-table(pred=factor(**Visc_injury**_rpart_predict), ref= **PCNL_visc_injury**_test1$**Visc_injury**)

confusionMatrix(**Visc_injury**_rpart_tb)

Error in !all.equal(nrow(data), ncol(data)) : invalid argument type

res_**Visc_injury**_rpart<- evalm(**Visc_injury**_rpart)

Confusion Matrix and Statistics

No Yes

No 1321	5

Yes	0	0

Accuracy : 0.9962

95% CI : (0.9912, 0.9988)

No Information Rate : 0.9962

P-Value [Acc > NIR] : 0.61596

Kappa : 0

Mcnemar's Test P-Value : 0.07364

Sensitivity : 1.0000

Specificity : 0.0000

Pos Pred Value : 0.9962

Neg Pred Value : NaN

Prevalence : 0.9962

Detection Rate : 0.9962

Detection Prevalence : 1.0000

Balanced Accuracy : 0.5000

'Positive' Class : No

***MLeval: Machine Learning Model Evaluation***

Input: caret train function object

Averaging probs.

Group 1 type: repeatedcv

Observations: 3092

Number of groups: 1

Observations per group: 3092

Positive: Yes

Negative: No

Group: Group 1

Positive: 11

Negative: 3081

***Performance Metrics***

Group 1 Optimal Informedness = 0

Group 1 AUC-ROC = 0.46

**#XGBoost**

**Visc_injury**_xgboost<-train(**Visc_injury** ~., data= **PCNL_visc_injury_train1**, method="xgbTree", trControl= **PCNL_visc_injury**_ctrl)

**Visc_injury**_xgboost_predict<-predict(**Visc_injury**_xgboost, **PCNL_visc_injury**_test1)

**Visc_injury**_xgboost_tb<-table(pred=factor(**Visc_injury**_xgboost_predict), ref= **PCNL_visc_injury**_test1$**Visc_injury**)

confusionMatrix(**Visc_injury**_xgboost_tb)

Error in !all.equal(nrow(data), ncol(data)) : invalid argument type

res_**Visc_injury**_xgboost<- evalm(**Visc_injury**_xgboost)

Confusion Matrix and Statistics

No Yes

No 1321	5

Yes	0	0

Accuracy : 0.9962

95% CI : (0.9912, 0.9988)

No Information Rate : 0.9962

P-Value [Acc > NIR] : 0.61596

Kappa : 0

Mcnemar's Test P-Value : 0.07364

Sensitivity : 1.0000

Specificity : 0.0000

Pos Pred Value : 0.9962

Neg Pred Value : NaN

Prevalence : 0.9962

Detection Rate : 0.9962

Detection Prevalence : 1.0000

Balanced Accuracy : 0.5000

'Positive' Class : No

***MLeval: Machine Learning Model Evaluation***

Input: caret train function object

Averaging probs.

Group 1 type: repeatedcv

Observations: 3092

Number of groups: 1

Observations per group: 3092

Positive: Yes

Negative: No

Group: Group 1

Positive: 11

Negative: 3081

***Performance Metrics***

Group 1 Optimal Informedness = 0.384762916408486

Group 1 AUC-ROC = 0.66

**#Logistic Regression**

**Visc_injury**_logit<-train(**Visc_injury** ~., data= **PCNL_visc_injury_train1**, method="LogitBoost", trControl= **PCNL_visc_injury**_ctrl)

**Visc_injury**_logit_predict<-predict(**Visc_injury**_logit, **PCNL_visc_injury**_test1)

**Visc_injury**_logit_tb<-table(pred=factor(**Visc_injury**_logit_predict), ref= **PCNL_visc_injury**_test1$**Visc_injury**)

confusionMatrix(**Visc_injury**_logit_tb)

Error in !all.equal(nrow(data), ncol(data)) : invalid argument type

res_**Visc_injury**_logit<- evalm(**Visc_injury**_logit)

Confusion Matrix and Statistics

No Yes

No 1321	5

Yes	0	0

Accuracy : 0.9962

95% CI : (0.9912, 0.9988)

No Information Rate : 0.9962

P-Value [Acc > NIR] : 0.61596

Kappa : 0

Mcnemar's Test P-Value : 0.07364

Sensitivity : 1.0000

Specificity : 0.0000

Pos Pred Value : 0.9962

Neg Pred Value : NaN

Prevalence : 0.9962

Detection Rate : 0.9962

Detection Prevalence : 1.0000

Balanced Accuracy : 0.5000

'Positive' Class : No

***MLeval: Machine Learning Model Evaluation***

Input: caret train function object

Averaging probs.

Group 1 type: repeatedcv

Observations: 3092

Number of groups: 1

Observations per group: 3092

Positive: Yes

Negative: No

Group: Group 1

Positive: 11

Negative: 3081

***Performance Metrics***

Group 1 Optimal Informedness = 0.218051990203889

Group 1 AUC-ROC = 0.56

**#Neural Net**

**Visc_injury**_nn<-train(**Visc_injury** ~., data= **PCNL_visc_injury_train1**, method="nnet", trControl= **PCNL_visc_injury**_ctrl)

**Visc_injury**_nn_predict<-predict(**Visc_injury**_nn, **PCNL_visc_injury**_test1)

**Visc_injury**_nn_tb<-table(pred=factor(**Visc_injury**_nn_predict), ref= **PCNL_visc_injury**_test1$**Visc_injury**)

confusionMatrix(**Visc_injury**_nn_tb)

Error in !all.equal(nrow(data), ncol(data)) : invalid argument type

res_**Visc_injury**_nn<- evalm(**Visc_injury**_nn)

Confusion Matrix and Statistics

No Yes

No 1321	5

Yes	0	0

Accuracy : 0.9962

95% CI : (0.9912, 0.9988)

No Information Rate : 0.9962

P-Value [Acc > NIR] : 0.61596

Kappa : 0

Mcnemar's Test P-Value : 0.07364

Sensitivity : 1.0000

Specificity : 0.0000

Pos Pred Value : 0.9962

Neg Pred Value : NaN

Prevalence : 0.9962

Detection Rate : 0.9962

Detection Prevalence : 1.0000

Balanced Accuracy : 0.5000

'Positive' Class : No

***MLeval: Machine Learning Model Evaluation***

Input: caret train function object

Averaging probs.

Group 1 type: repeatedcv

Observations: 3092

Number of groups: 1

Observations per group: 3092

Positive: Yes

Negative: No

Group: Group 1

Positive: 11

Negative: 3081

***Performance Metrics***

Group 1 Optimal Informedness = 0.0912041544952937

Group 1 AUC-ROC = 0.47

**#Bayesian Generalised Linear Model**

**Visc_injury**_bayes<-train(**Visc_injury** ~., data= **PCNL_visc_injury_train1**, method="bayesglm", trControl= **PCNL_visc_injury**_ctrl)

**Visc_injury**_bayes_predict<-predict(**Visc_injury**_bayes, **PCNL_visc_injury**_test1)

**Visc_injury**_bayes_tb<-table(pred=factor(**Visc_injury**_bayes_predict), ref= **PCNL_visc_injury**_test1$**Visc_injury**)

confusionMatrix(**Visc_injury**_bayes_tb)

res_**Visc_injury**_bayes<- evalm(**Visc_injury**_bayes)

Confusion Matrix and Statistics

No Yes

No 1321	5

Yes	0	0

Accuracy : 0.9962

95% CI : (0.9912, 0.9988)

No Information Rate : 0.9962

P-Value [Acc > NIR] : 0.61596

Kappa : 0

Mcnemar's Test P-Value : 0.07364

Sensitivity : 1.0000

Specificity : 0.0000

Pos Pred Value : 0.9962

Neg Pred Value : NaN

Prevalence : 0.9962

Detection Rate : 0.9962

Detection Prevalence : 1.0000

Balanced Accuracy : 0.5000

'Positive' Class : No

***MLeval: Machine Learning Model Evaluation***

Input: caret train function object

Averaging probs.

Group 1 type: repeatedcv

Observations: 3092

Number of groups: 1

Observations per group: 3092

Positive: Yes

Negative: No

Group: Group 1

Positive: 11

Negative: 3081

***Performance Metrics***

Group 1 Optimal Informedness = 0.437048183883627

Group 1 AUC-ROC = 0.77

**#Generate ROC curves**

res_**File_Name**_rf<-evalm(**File_Name**_rf)

res_**File_Name**_rpart<- evalm(**File_Name**_rpart)

res_**File_Name**_xgboost<- evalm(**File_Name**_xgboost)

res_**File_Name**_logit<- evalm(**File_Name**_logit)

res_**File_Name**_nn<- evalm(**File_Name**_nn)

res_**File_Name**_bayes<- evalm(**File_Name**_bayes)

res_**File_Name**_rf2$roc

res_**File_Name**_rpart$roc

res_**File_Name**_xgboost$roc

res_**File_Name**_logit$roc

res_**File_Name**_nn$roc

res_**File_Name**_bayes$roc

res_**Visc_injury**<-evalm(list(**Visc_injury**_rf, **Visc_injury**_rpart, **Visc_injury**_xgboost, **Visc_injury**_logit, **Visc_injury**_nn, **Visc_injury**_bayes), gnames=c('Random Forest', 'Partitioning', 'Extreme Gradient Boosting', 'Logistic Regression', 'Neural Network', 'Bayesian Generalised Linear Model'))

#Deep NN

Visc_injury_keras<-keras_model_sequential()

Visc_injury_keras %>% layer_dense(units = 32, input_shape = c(43)) %>% layer_activation('relu') %>% layer_dense(units = 1) %>% layer_activation('sigmoid')

Visc_injury_keras %>% compile(loss='binary_crossentropy', optimizer='rmsprop', metrics='accuracy')

Visc_injury_keras %>% fit(PCNL_visc_injury_train_predictors4, PCNL_visc_injury_train_outcome4, epochs=200, batch_size=5, validation_split=0.2, verbose=2)

Visc_injury_keras_results<- Visc_injury_keras %>% evaluate(PCNL_visc_injury_test_predictors4, PCNL_visc_injury_test_outcome4, verbose=1)

Visc_injury_keras_prediction<- Visc_injury_keras%>%predict(PCNL_visc_injury_test_predictors4, batch_size=32, verbose=1)

Visc_injury_keras_prediction1<-round(Visc_injury_keras_prediction, digits=0)

Visc_injury_keras_tb<- table(PCNL_visc_injury_test_outcome4, Visc_injury_keras_prediction1)

Visc_injury_keras_roc<- roc(PCNL_visc_injury_test_outcome4, Visc_injury_keras_prediction1)

auc(Visc_injury_keras_roc)

Area under the curve: 0.5

Visc_injury_keras_tb

Visc_injury_keras_prediction1

PCNL_visc_injury_test_outcome4	0

0 1321

1	5

Visc_injury_keras_tb1<-cbind(Visc_injury_keras_tb, Yes)

colnames(Visc_injury_keras_tb1)<-c("0","1")

confusionMatrix(Visc_injury_keras_tb1)

Confusion Matrix and Statistics

0	1

0 1321	0

1	5	0

Accuracy : 0.9962

95% CI : (0.9912, 0.9988)

No Information Rate : 1

P-Value [Acc > NIR] : 1.00000

Kappa : 0

Mcnemar's Test P-Value : 0.07364

Sensitivity : 0.9962

Specificity : NA

Pos Pred Value : NA

Neg Pred Value : NA

Prevalence : 1.0000

Detection Rate : 0.9962

Detection Prevalence : 0.9962

Balanced Accuracy : NA

'Positive' Class : 0

6. Survival

#Create training/test sets:

PCNL_survival_1<-na.omit(PCNL_survival_coded)

PCNL_survival_2<-na.omit(PCNL_survival_non_coded)

set.seed(1234)

PCNL_survival_sample_coded<-sample(1:nrow(PCNL_survival_1), size=nrow(PCNL_survival_1) *0.7)

PCNL_survival_train1<- PCNL_survival_1[PCNL_survival_sample_coded,]

PCNL_survival_test1<- PCNL_survival_1[-PCNL_survival_sample_coded,]

PCNL_survival_train_predictors1<- PCNL_survival_train1[,-44]

PCNL_survival_test_predictors1<- PCNL_survival_test1[,-44]

PCNL_survival_sample_non_coded<- sample(1:nrow(PCNL_survival_2), size=nrow(PCNL_survival_2) *0.7)

PCNL_survival_train2<- PCNL_survival_2[PCNL_survival_sample_non_coded,]

PCNL_survival_test2<- PCNL_survival_2[-PCNL_survival_sample_non_coded,]

PCNL_survival_train_predictors2<- PCNL_survival_train2[,-44]

PCNL_survival_test_predictors2<- PCNL_survival_test2[,-44]

PCNL_survival_ctrl<-trainControl(method= "repeatedcv", number=10, classProbs=TRUE, savePredictions=TRUE)

#Coded outcomes

#coded outcomes

PCNL_survival3<-na.omit(PCNL_survival_coded2)

set.seed(1234)

PCNL_survival_sample_coded3<-sample(1:nrow(PCNL_survival3), size=nrow(PCNL_survival3) *0.7)

PCNL_survival_train3<-PCNL_survival3[PCNL_survival_sample_coded3,]

PCNL_survival_test3<-PCNL_survival3[-PCNL_survival_sample_coded3,]

PCNL_survival_train_predictors3<-PCNL_survival_train3[,-44]

PCNL_survival_test_predictors3<-PCNL_survival_test3[,-44]

PCNL_survival_train_outcome3<- PCNL_survival_train3[,-1:-43]

PCNL_survival_test_outcome3<- PCNL_survival_test3[,-1:-43]

#make matrices

PCNL_survival_train_predictors4<-as.matrix(PCNL_survival_train_predictors3)

PCNL_survival_train_outcome4<-as.matrix(PCNL_survival_train_outcome3)

PCNL_survival_test_predictors4<-as.matrix(PCNL_survival_test_predictors3)

PCNL_survival_test_outcome4<-as.matrix(PCNL_survival_test_outcome3)

**#Create training/test sets:**

**PCNL_survival**_1<-na.omit(**PCNL_survival**_coded)

**PCNL_survival**_2<-na.omit(**PCNL_survival**_non_coded)

set.seed(1234)

**PCNL_survival**_sample_coded<-sample(1:nrow(**PCNL_survival**_1), size=nrow(**PCNL_survival**_1) *0.7)

**PCNL_survival**_train1<- **PCNL_survival**_1[**PCNL_survival**_sample_coded,]

**PCNL_survival**_test1<- **PCNL_survival**_1[-**PCNL_survival**_sample_coded,]

**PCNL_survival**_train_predictors1<- **PCNL_survival**_train1[,-44]

**PCNL_survival**_test_predictors1<- **PCNL_survival**_test1[,-44]

**PCNL_survival**_sample_non_coded<- sample(1:nrow(**PCNL_survival**_2), size=nrow(**PCNL_survival**_2) *0.7)

**PCNL_survival**_train2<- **PCNL_survival**_2[**PCNL_survival**_sample_non_coded,]

**PCNL_survival**_test2<- **PCNL_survival**_2[-**PCNL_survival**__sample_non_coded,]

**PCNL_survival**_train_predictors2<- **PCNL_survival**_train2[,-44]

**PCNL_survival**_test_predictors2<- **PCNL_survival**_test2[,-44]

**PCNL_survival**_ctrl<-trainControl(method= "repeatedcv", number=10, classProbs=TRUE, savePredictions=TRUE)

**#coded outcomes**

PCNL_survival3<-na.omit(PCNL_survival_coded2)

set.seed(1234)

PCNL_survival_sample_coded3<-sample(1:nrow(PCNL_survival3), size=nrow(PCNL_survival3) *0.7)

PCNL_survival_train3<-PCNL_survival3[PCNL_survival_sample_coded3,]

PCNL_survival_test3<-PCNL_survival3[-PCNL_survival_sample_coded3,]

PCNL_survival_train_predictors3<-PCNL_survival_train3[,-44]

PCNL_survival_test_predictors3<-PCNL_survival_test3[,-44]

PCNL_survival_train_outcome3<- PCNL_survival_train3[,-1:-43]

PCNL_survival_test_outcome3<- PCNL_survival_test3[,-1:-43]

#make matrices

PCNL_survival_train_predictors4<-as.matrix(PCNL_survival_train_predictors3)

PCNL_survival_train_outcome4<-as.matrix(PCNL_survival_train_outcome3)

PCNL_survival_test_predictors4<-as.matrix(PCNL_survival_test_predictors3)

PCNL_survival_test_outcome4<-as.matrix(PCNL_survival_test_outcome3)

**Random Forests**

**PCNL_survival**_rf<- train(**Survival** ~., data= **PCNL_survival_train1**, method="cforest", trControl= **PCNL_survival**_ctrl)

**PCNL_survival**_rf_predict<-predict(**PCNL_survival**_rf, newdata= **PCNL_survival**_test_predictors1)

**PCNL_survival**_rf_tb<-table(**PCNL_survival**_rf_predict, ref= **PCNL_survival**_test1$**Survival**)

confusionMatrix(**PCNL_survival**_rf_tb)

res_**PCNL_survival**_rf<-evalm(**PCNL_survival**_rf)

***MLeval: Machine Learning Model Evaluation***

Input: caret train function object

Averaging probs.

Group 1 type: repeatedcv

Observations: 3092

Number of groups: 1

Observations per group: 3092

Positive: Yes

Negative: No

Group: Group 1

Positive: 11

Negative: 3081

***Performance Metrics***

Group 1 Optimal Informedness = 0.444247735386976

Group 1 AUC-ROC = 0.72

Confusion Matrix and Statistics

ref

PCNL_survival_rf_predict	No Yes

No 1324	2

Yes	0	0

Accuracy : 0.9985

95% CI : (0.9946, 0.9998)

No Information Rate : 0.9985

P-Value [Acc > NIR] : 0.6767

Kappa : 0

Mcnemar's Test P-Value : 0.4795

Sensitivity : 1.0000

Specificity : 0.0000

Pos Pred Value : 0.9985

Neg Pred Value : NaN

Prevalence : 0.9985

Detection Rate : 0.9985

Detection Prevalence : 1.0000

Balanced Accuracy : 0.5000

'Positive' Class : No

**Partitioning**

**PCNL_survival**_rpart<-train(**Survival** ~ ., data= **PCNL_survival**_train1, method="rpart", trControl= **PCNL_survival**_ctrl)

**PCNL_survival**_rpart_predict<-predict(**PCNL_survival**_rpart, newdata= **PCNL_survival**_test1)

**PCNL_survival**_rpart_tb<-table(pred=factor(**PCNL_survival**_rpart_predict), ref= **PCNL_survival**_test1$**Survival**)

confusionMatrix(**PCNL_survival**_rpart_tb)

Error in !all.equal(nrow(data), ncol(data)) : invalid argument type

res_**PCNL_survival**_rpart<- evalm(**PCNL_survival**_rpart)

Confusion Matrix and Statistics

0	1

0 1323	3

1	0	0

Accuracy : 0.9977

95% CI : (0.9934, 0.9995)

No Information Rate : 0.9977

P-Value [Acc > NIR] : 0.6472

Kappa : 0

Mcnemar's Test P-Value : 0.2482

Sensitivity : 1.0000

Specificity : 0.0000

Pos Pred Value : 0.9977

Neg Pred Value : NaN

Prevalence : 0.9977

Detection Rate : 0.9977

Detection Prevalence : 1.0000

Balanced Accuracy : 0.5000

'Positive' Class : 0

***MLeval: Machine Learning Model Evaluation***

Input: caret train function object

Averaging probs.

Group 1 type: repeatedcv

Observations: 3092

Number of groups: 1

Observations per group: 3092

Positive: Yes

Negative: No

Group: Group 1

Positive: 11

Negative: 3081

***Performance Metrics***

Group 1 Optimal Informedness = 0

Group 1 AUC-ROC = 0.46

**XGBoost**

**PCNL_survival**_xgboost<-train(**Survival** ~., data= **PCNL_survival**_train1, method="xgbTree", trControl= **PCNL_survival**_ctrl)

**PCNL_survival**_xgboost_predict<-predict(**PCNL_survival**_xgboost, **PCNL_survival**_test1)

**PCNL_survival**_xgboost_tb<-table(pred=factor(**PCNL_survival**_xgboost_predict), ref= **PCNL_survival**_test1$**Survival**)

confusionMatrix(**PCNL_survival**_xgboost_tb)

Error in !all.equal(nrow(data), ncol(data)) : invalid argument type

res_**PCNL_survival**_xgboost<- evalm(**PCNL_survival**_xgboost)

**#Does not predict any as having visceral injury – therefore cannot generate confusion matrix**

Confusion Matrix and Statistics

No Yes

No 1324	2

Yes	0	0

Accuracy : 0.9985

95% CI : (0.9946, 0.9998)

No Information Rate : 0.9985

P-Value [Acc > NIR] : 0.6767

Kappa : 0

Mcnemar's Test P-Value : 0.4795

Sensitivity : 1.0000

Specificity : 0.0000

Pos Pred Value : 0.9985

Neg Pred Value : NaN

Prevalence : 0.9985

Detection Rate : 0.9985

Detection Prevalence : 1.0000

Balanced Accuracy : 0.5000

'Positive' Class : No

***MLeval: Machine Learning Model Evaluation***

Input: caret train function object

Averaging probs.

Group 1 type: repeatedcv

Observations: 3092

Number of groups: 1

Observations per group: 3092

Positive: Yes

Negative: No

Group: Group 1

Positive: 11

Negative: 3081

***Performance Metrics***

Group 1 Optimal Informedness = 0.250184414741377

Group 1 AUC-ROC = 0.55

**Logistic Regression**

**PCNL_survival**_logit<-train(**Survival** ~., data= **PCNL_survival**_train1, method="LogitBoost", trControl= **PCNL_survival**_ctrl)

**PCNL_survival**_logit_predict<-predict(**PCNL_survival**_logit, **PCNL_survival**_test1)

**PCNL_survival**_logit_tb<-table(pred=factor(**PCNL_survival**_logit_predict), ref= **PCNL_survival**_test1$**Survival**)

confusionMatrix(**PCNL_survival**_logit_tb)

Error in !all.equal(nrow(data), ncol(data)) : invalid argument type

res_**PCNL_survival**_logit<- evalm(**PCNL_survival**_logit)

Confusion Matrix and Statistics

No Yes

No 1324	2

Yes	0	0

Accuracy : 0.9985

95% CI : (0.9946, 0.9998)

No Information Rate : 0.9985

P-Value [Acc > NIR] : 0.6767

Kappa : 0

Mcnemar's Test P-Value : 0.4795

Sensitivity : 1.0000

Specificity : 0.0000

Pos Pred Value : 0.9985

Neg Pred Value : NaN

Prevalence : 0.9985

Detection Rate : 0.9985

Detection Prevalence : 1.0000

Balanced Accuracy : 0.5000

'Positive' Class : No

***MLeval: Machine Learning Model Evaluation***

Input: caret train function object

Averaging probs.

Group 1 type: repeatedcv

Observations: 3092

Number of groups: 1

Observations per group: 3092

Positive: Yes

Negative: No

Group: Group 1

Positive: 11

Negative: 3081

***Performance Metrics***

Group 1 Optimal Informedness = 0.12734944380514

Group 1 AUC-ROC = 0.55

**Neural Net**

**PCNL_survival**_nn<-train(**Survival** ~., data= **PCNL_survival**_train1, method="nnet", trControl= **PCNL_survival**_ctrl)

**PCNL_survival**_nn_predict<-predict(**PCNL_survival**_nn, **PCNL_survival**_test1)

**PCNL_survival**_nn_tb<-table(pred=factor(**PCNL_survival**_nn_predict), ref= **PCNL_survival**_test1$**Survival**)

confusionMatrix(**PCNL_survival**_nn_tb)

Error in !all.equal(nrow(data), ncol(data)) : invalid argument type

res_**PCNL_survival**_nn<- evalm(**PCNL_survival**_nn)

Confusion Matrix and Statistics

No Yes

No 1324	2

Yes	0	0

Accuracy : 0.9985

95% CI : (0.9946, 0.9998)

No Information Rate : 0.9985

P-Value [Acc > NIR] : 0.6767

Kappa : 0

Mcnemar's Test P-Value : 0.4795

Sensitivity : 1.0000

Specificity : 0.0000

Pos Pred Value : 0.9985

Neg Pred Value : NaN

Prevalence : 0.9985

Detection Rate : 0.9985

Detection Prevalence : 1.0000

Balanced Accuracy : 0.5000

'Positive' Class : No

***MLeval: Machine Learning Model Evaluation***

Input: caret train function object

Averaging probs.

Group 1 type: repeatedcv

Observations: 3092

Number of groups: 1

Observations per group: 3092

Positive: Yes

Negative: No

Group: Group 1

Positive: 11

Negative: 3081

***Performance Metrics***

Group 1 Optimal Informedness = 0

Group 1 AUC-ROC = 0.46

**Bayesian Generalised Linear Model**

**PCNL_survival**_bayes<-train(**Survival** ~., data= **PCNL_survival**_train1, method="bayesglm", trControl= **PCNL_survival**_ctrl)

**PCNL_survival**_bayes_predict<-predict(**PCNL_survival**_bayes, **PCNL_survival**_test1)

**PCNL_survival**_bayes_tb<-table(pred=factor(**PCNL_survival**_bayes_predict), ref= **PCNL_survival**_test1$**Survival**)

confusionMatrix(**PCNL_survival**_bayes_tb)

Error in !all.equal(nrow(data), ncol(data)) : invalid argument type

res_**PCNL_survival**_bayes<- evalm(**PCNL_survival**_bayes)

Confusion Matrix and Statistics

No Yes

No 1324	2

Yes	0	0

Accuracy : 0.9985

95% CI : (0.9946, 0.9998)

No Information Rate : 0.9985

P-Value [Acc > NIR] : 0.6767

Kappa : 0

Mcnemar's Test P-Value : 0.4795

Sensitivity : 1.0000

Specificity : 0.0000

Pos Pred Value : 0.9985

Neg Pred Value : NaN

Prevalence : 0.9985

Detection Rate : 0.9985

Detection Prevalence : 1.0000

Balanced Accuracy : 0.5000

'Positive' Class : No

***MLeval: Machine Learning Model Evaluation***

Input: caret train function object

Averaging probs.

Group 1 type: repeatedcv

Observations: 3092

Number of groups: 1

Observations per group: 3092

Positive: Yes

Negative: No

Group: Group 1

Positive: 11

Negative: 3081

***Performance Metrics***

Group 1 Optimal Informedness = 0.468029860434924

Group 1 AUC-ROC = 0.77

**Generate ROC curves**

res_**PCNL_survival** <-evalm(list(**PCNL_survival**_rf, **PCNL_survival**_rpart, **PCNL_survival**_xgboost, **PCNL_survival**_logit, **PCNL_survival**_nn, **PCNL_survival**_bayes), gnames=c('Random Forest', 'Partitioning', 'Extreme Gradient Boosting', 'Logistic Regression', 'Neural Network', 'Bayesian Generalised Linear Model'))

#Deep NN

Survival_keras<-keras_model_sequential()

Survival_keras %>% layer_dense(units = 32, input_shape = c(43)) %>% layer_activation('relu') %>% layer_dense(units = 1) %>% layer_activation('sigmoid')

Survival_keras %>% compile(loss='binary_crossentropy', optimizer='rmsprop', metrics='accuracy')

Survival_keras %>% fit(PCNL_survival_train_predictors4, PCNL_survival_train_outcome4, epochs=200, batch_size=5, validation_split=0.2, verbose=2)

Survival_keras_results<- Survival_keras %>% evaluate(PCNL_survival_test_predictors4, Survival_test_outcome4, verbose=1)

Survival_keras_prediction<- Survival_keras%>%predict(PCNL_survival_test_predictors4, batch_size=32, verbose=1)

Survival_keras_prediction1<-round(Survival_keras_prediction, digits=0)

Survival_keras_tb<- table(PCNL_survival_test_outcome4, Survival_keras_prediction1)

Survival_keras_roc<- roc(PCNL_survival_test_outcome4, Survival_keras_prediction1)

auc(Survival_keras_roc)

Area under the curve: 0.4989

confusionMatrix(Survival_keras_tb)

Confusion Matrix and Statistics

Survival_keras_prediction1

PCNL_survival_test_outcome4	0	1

0 1320	3

1	3	0

Accuracy : 0.9955

95% CI : (0.9902, 0.9983)

No Information Rate : 0.9977

P-Value [Acc > NIR] : 0.9667

Kappa : -0.0023

Mcnemar's Test P-Value : 1.0000

Sensitivity : 0.9977

Specificity : 0.0000

Pos Pred Value : 0.9977

Neg Pred Value : 0.0000

Prevalence : 0.9977

Detection Rate : 0.9955

Detection Prevalence : 0.9977

Balanced Accuracy : 0.4989

'Positive' Class : 0

7. Transfusion

**#Create training/test sets:**

**PCNL_transfusion**_1<-na.omit(**PCNL_transfusion**_coded)

**PCNL_transfusion**_2<-na.omit(**PCNL_transfusion**_non_coded)

set.seed(1234)

**PCNL_transfusion**_sample_coded<-sample(1:nrow(**PCNL_transfusion**_1), size=nrow(**PCNL_transfusion**_1) *0.7)

**PCNL_transfusion**_train1<- **PCNL_transfusion**_1[**PCNL_transfusion**_sample_coded,]

**PCNL_transfusion**_test1<- **PCNL_transfusion**_1[-**PCNL_transfusion**_sample_coded,]

**PCNL_transfusion**_train_predictors1<- **PCNL_transfusion**_train1[,-44]

**PCNL_transfusion**_test_predictors1<- **PCNL_transfusion**_test1[,-44]

**PCNL_transfusion**_sample_non_coded<- sample(1:nrow(**PCNL_transfusion**_2), size=nrow(**PCNL_transfusion**_2) *0.7)

**PCNL_transfusion**_train2<- **PCNL_transfusion**_2[**PCNL_transfusion**_sample_non_coded,]

**PCNL_transfusion**_test2<- **PCNL_transfusion**_2[-**PCNL_transfusion**_sample_non_coded,]

**PCNL_transfusion**_train_predictors2<- **PCNL_transfusion**_train2[,-44]

**PCNL_transfusion**_test_predictors2<- **PCNL_transfusion**_test2[,-44]

**PCNL_transfusion**_ctrl<-trainControl(method= "repeatedcv", number=10, classProbs=TRUE, savePredictions=TRUE)

**Random Forests**

**PCNL_transfusion**_rf<- train(**Transfusion** ~., data= **PCNL_transfusion**_train1, method="cforest", trControl= **PCNL_transfusion**_ctrl)

**PCNL_transfusion**_rf_predict<-predict(**PCNL_transfusion**_rf, newdata= **PCNL_transfusion**_test_predictors1)

**PCNL_transfusion**_rf_tb<-table(**PCNL_transfusion**_rf_predict, ref= **PCNL_transfusion**_test1$**Transfusion**)

confusionMatrix(**PCNL_transfusion**_rf_tb)

res_**PCNL_transfusion**_rf<-evalm(**PCNL_transfusion**_rf)

***MLeval: Machine Learning Model Evaluation***

Input: caret train function object

Averaging probs.

Group 1 type: repeatedcv

Observations: 3088

Number of groups: 1

Observations per group: 3088

Positive: Yes

Negative: No

Group: Group 1

Positive: 132

Negative: 2956

***Performance Metrics***

Group 1 Optimal Informedness = 0.974228072333621

Group 1 AUC-ROC = 0.98

Confusion Matrix and Statistics

ref

PCNL_transfusion_rf_predict	No Yes

No 1259	2

Yes	1	62

Accuracy : 0.9977

95% CI : (0.9934, 0.9995)

No Information Rate : 0.9517

P-Value [Acc > NIR] : <2e-16

Kappa : 0.9752

Mcnemar's Test P-Value : 1

Sensitivity : 0.9992

Specificity : 0.9688

Pos Pred Value : 0.9984

Neg Pred Value : 0.9841

Prevalence : 0.9517

Detection Rate : 0.9509

Detection Prevalence : 0.9524

Balanced Accuracy : 0.9840

'Positive' Class : No

**Partitioning**

**PCNL_transfusion**_rpart<-train(**Transfusion** ~ ., data= **PCNL_transfusion**_train1, method="rpart", trControl= **PCNL_transfusion**_ctrl)

**PCNL_transfusion**_rpart_predict<-predict(**PCNL_transfusion**_rpart, newdata= **PCNL_transfusion**_test1)

**PCNL_transfusion**_rpart_tb<-table(pred=factor(**PCNL_transfusion**_rpart_predict), ref= **PCNL_transfusion**_test1$**Transfusion**)

confusionMatrix(**PCNL_transfusion**_rpart_tb)

res_**PCNL_transfusion**_rpart<- evalm(**PCNL_transfusion**_rpart)

Confusion Matrix and Statistics

ref

pred	No Yes

No 1260	7

Yes	0	57

Accuracy : 0.9947

95% CI : (0.9891, 0.9979)

No Information Rate : 0.9517

P-Value [Acc > NIR] : < 2e-16

Kappa : 0.9394

Mcnemar's Test P-Value : 0.02334

Sensitivity : 1.0000

Specificity : 0.8906

Pos Pred Value : 0.9945

Neg Pred Value : 1.0000

Prevalence : 0.9517

Detection Rate : 0.9517

Detection Prevalence : 0.9569

Balanced Accuracy : 0.9453

'Positive' Class : No

***MLeval: Machine Learning Model Evaluation***

Input: caret train function object

Averaging probs.

Group 1 type: repeatedcv

Observations: 3088

Number of groups: 1

Observations per group: 3088

Positive: Yes

Negative: No

Group: Group 1

Positive: 132

Negative: 2956

***Performance Metrics***

Group 1 Optimal Informedness = 0.877096403821708

Group 1 AUC-ROC = 0.96

**XGBoost**

**PCNL_transfusion**_xgboost<-train(**Transfusion** ~., data= **PCNL_transfusion**_train1, method="xgbTree", trControl= **PCNL_transfusion**_ctrl)

**PCNL_transfusion**_xgboost_predict<-predict(**PCNL_transfusion**_xgboost, **PCNL_transfusion**_test1)

**PCNL_transfusion**_xgboost_tb<-table(pred=factor(**PCNL_transfusion**_xgboost_predict), ref= **PCNL_transfusion**_test1$**Transfusion**)

confusionMatrix(**PCNL_transfusion**_xgboost_tb)

res_**PCNL_transfusion**_xgboost<- evalm(**PCNL_transfusion**_xgboost)

***MLeval: Machine Learning Model Evaluation***

Input: caret train function object

Averaging probs.

Group 1 type: repeatedcv

Observations: 3088

Number of groups: 1

Observations per group: 3088

Positive: Yes

Negative: No

Group: Group 1

Positive: 132

Negative: 2956

***Performance Metrics***

Group 1 Optimal Informedness = 0.973889777340386

Group 1 AUC-ROC = 0.99

Confusion Matrix and Statistics

ref

pred	No Yes

No 1260	2

Yes	0	62

Accuracy : 0.9985

95% CI : (0.9946, 0.9998)

No Information Rate : 0.9517

P-Value [Acc > NIR] : <2e-16

Kappa : 0.9833

Mcnemar's Test P-Value : 0.4795

Sensitivity : 1.0000

Specificity : 0.9688

Pos Pred Value : 0.9984

Neg Pred Value : 1.0000

Prevalence : 0.9517

Detection Rate : 0.9517

Detection Prevalence : 0.9532

Balanced Accuracy : 0.9844

'Positive' Class : No

**Logistic Regression**

**PCNL_transfusion**_logit<-train(**Transfusion** ~., data= **PCNL_transfusion**_train1, method="LogitBoost", trControl= **PCNL_transfusion**_ctrl)

**PCNL_transfusion**_logit_predict<-predict(**PCNL_transfusion**_logit, **PCNL_transfusion**_test1)

**PCNL_transfusion**_logit_tb<-table(pred=factor(**PCNL_transfusion**_logit_predict), ref= **PCNL_transfusion**_test1$**Transfusion**)

confusionMatrix(**PCNL_transfusion**_logit_tb)

res_**PCNL_transfusion**_logit<- evalm(**PCNL_transfusion**_logit)

***MLeval: Machine Learning Model Evaluation***

Input: caret train function object

Averaging probs.

Group 1 type: repeatedcv

Observations: 3088

Number of groups: 1

Observations per group: 3088

Positive: Yes

Negative: No

Group: Group 1

Positive: 132

Negative: 2956

***Performance Metrics***

Group 1 Optimal Informedness = 0.949809324640177

Group 1 AUC-ROC = 0.99

Confusion Matrix and Statistics

ref

pred	No Yes

No 1258	5

Yes	2	59

Accuracy : 0.9947

95% CI : (0.9891, 0.9979)

No Information Rate : 0.9517

P-Value [Acc > NIR] : <2e-16

Kappa : 0.9412

Mcnemar's Test P-Value : 0.4497

Sensitivity : 0.9984

Specificity : 0.9219

Pos Pred Value : 0.9960

Neg Pred Value : 0.9672

Prevalence : 0.9517

Detection Rate : 0.9502

Detection Prevalence : 0.9539

Balanced Accuracy : 0.9601

'Positive' Class : No

**Neural Net**

**PCNL_transfusion**_nn<-train(**Transfusion** ~., data= **PCNL_transfusion**_train1, method="nnet", trControl= **PCNL_transfusion**_ctrl)

**PCNL_transfusion**_nn_predict<-predict(**PCNL_transfusion**_nn, **PCNL_transfusion**_test1)

**PCNL_transfusion**_nn_tb<-table(pred=factor(**PCNL_transfusion**_nn_predict), ref= **PCNL_transfusion**_test1$**Transfusion**)

confusionMatrix(**PCNL_transfusion**_nn_tb)

res_**PCNL_transfusion**_nn<- evalm(**PCNL_transfusion**_nn)

***MLeval: Machine Learning Model Evaluation***

Input: caret train function object

Averaging probs.

Group 1 type: repeatedcv

Observations: 3088

Number of groups: 1

Observations per group: 3088

Positive: Yes

Negative: No

Group: Group 1

Positive: 132

Negative: 2956

***Performance Metrics***

Group 1 Optimal Informedness = 0.872288514372412

Group 1 AUC-ROC = 0.97

Confusion Matrix and Statistics

ref

pred	No Yes

No 1259	13

Yes	1	51

Accuracy : 0.9894

95% CI : (0.9823, 0.9942)

No Information Rate : 0.9517

P-Value [Acc > NIR] : 1.693e-14

Kappa : 0.8738

Mcnemar's Test P-Value : 0.003283

Sensitivity : 0.9992

Specificity : 0.7969

Pos Pred Value : 0.9898

Neg Pred Value : 0.9808

Prevalence : 0.9517

Detection Rate : 0.9509

Detection Prevalence : 0.9607

Balanced Accuracy : 0.8980

'Positive' Class : No

**Bayesian Generalised Linear Model**

**PCNL_transfusion**_bayes<-train(**Transfusion** ~., data= **PCNL_transfusion**_train1, method="bayesglm", trControl= **PCNL_transfusion**_ctrl)

**PCNL_transfusion**_bayes_predict<-predict(**PCNL_transfusion**_bayes, **PCNL_transfusion**_test1)

**PCNL_transfusion**_bayes_tb<-table(pred=factor(**PCNL_transfusion**_bayes_predict), ref= **PCNL_transfusion**_test1$**Transfusion**)

confusionMatrix(**PCNL_transfusion**_bayes_tb)

res_**PCNL_transfusion**_bayes<- evalm(**PCNL_transfusion**_bayes)

***MLeval: Machine Learning Model Evaluation***

Input: caret train function object

Averaging probs.

Group 1 type: repeatedcv

Observations: 3088

Number of groups: 1

Observations per group: 3088

Positive: Yes

Negative: No

Group: Group 1

Positive: 132

Negative: 2956

***Performance Metrics***

Group 1 Optimal Informedness = 0.906312789601017

Group 1 AUC-ROC = 0.97

Confusion Matrix and Statistics

ref

pred	No Yes

No 1259	10

Yes	1	54

Accuracy : 0.9917

95% CI : (0.9852, 0.9958)

No Information Rate : 0.9517

P-Value [Acc > NIR] : < 2e-16

Kappa : 0.9032

Mcnemar's Test P-Value : 0.01586

Sensitivity : 0.9992

Specificity : 0.8438

Pos Pred Value : 0.9921

Neg Pred Value : 0.9818

Prevalence : 0.9517

Detection Rate : 0.9509

Detection Prevalence : 0.9585

Balanced Accuracy : 0.9215

'Positive' Class : No

**Generate ROC curves**

res_**PCNL_transfusion**<-evalm(list(**PCNL_transfusion**_rf, **PCNL_transfusion**_rpart, **PCNL_transfusion**_xgboost, **PCNL_transfusion**_logit, **PCNL_transfusion**_nn, **PCNL_transfusion**_bayes), gnames=c('Random Forest', 'Partitioning', 'Extreme Gradient Boosting', 'Logistic Regression', 'Neural Network', 'Bayesian Generalised Linear Model'))

#Deep Neural Net

PCNL_transfusion_train_predictors3<-as.matrix(PCNL_transfusion_train_predictors1)

PCNL_transfusion_train_outcome<-ifelse(PCNL_transfusion_train1$Transfusion=="Yes", 1,0)

PCNL_transfusion_test_predictors3<-as.matrix(PCNL_transfusion_test_predictors1)

PCNL_transfusion_test_outcome<-ifelse(PCNL_transfusion_test1$Transfusion=="Yes", 1,0)

**#Deep NN**

library(keras)

library(tensorflow)

library(pROC)

Transfusion_keras<-keras_model_sequential()

Transfusion_keras %>%

layer_dense(units = 32, input_shape = c(43)) %>%

layer_activation('relu') %>%

layer_dense(units = 1) %>%

layer_activation('sigmoid')

Transfusion_keras %>% compile(loss='binary_crossentropy', optimizer='rmsprop', metrics='accuracy')

Transfusion_keras %>% fit(PCNL_transfusion_train_predictors3, PCNL_transfusion_train_outcome, epochs=200, batch_size=5, validation_split=0.2, verbose=2)

Transfusion_keras_results<-Transfusion_keras %>% evaluate(PCNL_transfusion_test_predictors3, PCNL_transfusion_test_outcome, verbose=1)

Transfusion_keras_prediction<- Transfusion_keras%>%predict(PCNL_transfusion_test_predictors3, batch_size=32, verbose=1)

Transfusion_keras_prediction1<-round(Transfusion_keras_prediction, digits=0)

Transfusion_keras_tb<- table(PCNL_transfusion_test_outcome, Transfusion_keras_prediction1)

Transfusion_keras_roc<- roc(PCNL_transfusion_test_outcome, Transfusion_keras_prediction1)

auc(Transfusion_keras_roc)

Area under the curve: 0.8742

confusionMatrix(Transfusion_keras_tb)

Confusion Matrix and Statistics

Transfusion_keras_prediction1

PCNL_transfusion_test_outcome	0	1

0 1258	2

1	16	48

Accuracy : 0.9864

95% CI : (0.9786, 0.9919)

No Information Rate : 0.9622

P-Value [Acc > NIR] : 1.179e-07

Kappa : 0.8351

Mcnemar's Test P-Value : 0.002183

Sensitivity : 0.9874

Specificity : 0.9600

Pos Pred Value : 0.9984

Neg Pred Value : 0.7500

Prevalence : 0.9622

Detection Rate : 0.9502

Detection Prevalence : 0.9517

Balanced Accuracy : 0.9737

'Positive' Class : 0

8. **Post-operative Infection**

**#Create training/test sets:**

**PCNL_Post_Infection**_1<-na.omit(**PCNL_Post_Infection**_coded)

**PCNL_Post_Infection**_2<-na.omit(**PCNL_Post_Infection**_non_coded)

set.seed(1234)

**PCNL_Post_Infection**_sample_coded<-sample(1:nrow(**PCNL_Post_Infection**_1), size=nrow(**PCNL_Post_Infection**_1) *0.7)

**PCNL_Post_Infection**_train1<- **PCNL_Post_Infection**_1[**PCNL_Post_Infection**_sample_coded,]

**PCNL_Post_Infection**_test1<- **PCNL_Post_Infection**_1[-**PCNL_Post_Infection**_sample_coded,]

**PCNL_Post_Infection**_train_predictors1<- **PCNL_Post_Infection**_train1[,-44]

**PCNL_Post_Infection**_test_predictors1<- **PCNL_Post_Infection**_test1[,-44]

**PCNL_Post_Infection**_sample_non_coded<- sample(1:nrow(**PCNL_Post_Infection**_2), size=nrow(**PCNL_Post_Infection**_2) *0.7)

**PCNL_Post_Infection**_train2<- **PCNL_Post_Infection**_2[**PCNL_Post_Infection**_sample_non_coded,]

**PCNL_Post_Infection**_test2<- **PCNL_Post_Infection**_2[-**PCNL_Post_Infection**_sample_non_coded,]

**PCNL_Post_Infection**_train_predictors2<- **PCNL_Post_Infection**_train2[,-44]

**PCNL_Post_Infection**_test_predictors2<- **PCNL_Post_Infection**_test2[,-44]

**PCNL_Post_Infection**_ctrl<-trainControl(method= "repeatedcv", number=10, classProbs=TRUE, savePredictions=TRUE)

**Random Forests**

**PCNL_Post_Infection**_rf<- train(**Post_infection** ~., data= **PCNL_Post_Infection**_train1, method="cforest", trControl= **PCNL_Post_Infection**_ctrl)

**PCNL_Post_Infection**_rf_predict<-predict(**PCNL_Post_Infection**_rf, newdata= **PCNL_Post_Infection**_test_predictors1)

**PCNL_Post_Infection**_rf_tb<-table(**PCNL_Post_Infection**_rf_predict, ref= **PCNL_Post_Infection**_test1$**Post_infection**)

confusionMatrix(**PCNL_Post_Infection**_rf_tb)

res_**PCNL_Post_Infection**_rf<-evalm(**PCNL_Post_Infection**_rf)

***MLeval: Machine Learning Model Evaluation***

Input: caret train function object

Averaging probs.

Group 1 type: repeatedcv

Observations: 3092

Number of groups: 1

Observations per group: 3092

Positive: Yes

Negative: No

Group: Group 1

Positive: 684

Negative: 2408

***Performance Metrics***

Group 1 Optimal Informedness = 0.969633385790057

Group 1 AUC-ROC = 1

Confusion Matrix and Statistics

ref

PCNL_Post_Infection_rf_predict	No Yes

No 1029	12

Yes	15 270

Accuracy : 0.9796

95% CI : (0.9705, 0.9865)

No Information Rate : 0.7873

P-Value [Acc > NIR] : <2e-16

Kappa : 0.9394

Mcnemar's Test P-Value : 0.7003

Sensitivity : 0.9856

Specificity : 0.9574

Pos Pred Value : 0.9885

Neg Pred Value : 0.9474

Prevalence : 0.7873

Detection Rate : 0.7760

Detection Prevalence : 0.7851

Balanced Accuracy : 0.9715

'Positive' Class : No

**Partitioning**

**PCNL_Post_Infection**_rpart<-train(**Post_infection** ~ ., data= **PCNL_Post_Infection**_train1, method="rpart", trControl= **PCNL_Post_Infection**_ctrl)

**PCNL_Post_Infection**_rpart_predict<-predict(**PCNL_Post_Infection**_rpart, newdata= **PCNL_Post_Infection**_test1)

**PCNL_Post_Infection**_rpart_tb<-table(pred=factor(**PCNL_Post_Infection**_rpart_predict), ref= **PCNL_Post_Infection**_test1$**Post_infection**)

confusionMatrix(**PCNL_Post_Infection**_rpart_tb)

res_**PCNL_Post_Infection**_rpart<- evalm(**PCNL_Post_Infection**_rpart)

***MLeval: Machine Learning Model Evaluation***

Input: caret train function object

Averaging probs.

Group 1 type: repeatedcv

Observations: 3092

Number of groups: 1

Observations per group: 3092

Positive: Yes

Negative: No

Group: Group 1

Positive: 684

Negative: 2408

***Performance Metrics***

Group 1 Optimal Informedness = 0.934278525771794

Group 1 AUC-ROC = 0.97

Confusion Matrix and Statistics

ref

pred	No Yes

No 1029	21

Yes	15 261

Accuracy : 0.9729

95% CI : (0.9626, 0.9809)

No Information Rate : 0.7873

P-Value [Acc > NIR] : <2e-16

Kappa : 0.9183

Mcnemar's Test P-Value : 0.4047

Sensitivity : 0.9856

Specificity : 0.9255

Pos Pred Value : 0.9800

Neg Pred Value : 0.9457

Prevalence : 0.7873

Detection Rate : 0.7760

Detection Prevalence : 0.7919

Balanced Accuracy : 0.9556

'Positive' Class : No

**XGBoost**

**PCNL_Post_Infection**_xgboost<-train(**Post_infection** ~., data= **PCNL_Post_Infection**_train1, method="xgbTree", trControl= **PCNL_Post_Infection**_ctrl)

**PCNL_Post_Infection**_xgboost_predict<-predict(**PCNL_Post_Infection**_xgboost, **PCNL_Post_Infection**_test1)

**PCNL_Post_Infection**_xgboost_tb<-table(pred=factor(**PCNL_Post_Infection**_xgboost_predict), ref= **PCNL_Post_Infection**_test1$**Post_infection**)

confusionMatrix(**PCNL_Post_Infection**_xgboost_tb)

res_**PCNL_Post_Infection**_xgboost<- evalm(**PCNL_Post_Infection**_xgboost)

Confusion Matrix and Statistics

ref

pred	No Yes

No 1031	8

Yes	13 274

Accuracy : 0.9842

95% CI : (0.9759, 0.9902)

No Information Rate : 0.7873

P-Value [Acc > NIR] : <2e-16

Kappa : 0.953

Mcnemar's Test P-Value : 0.3827

Sensitivity : 0.9875

Specificity : 0.9716

Pos Pred Value : 0.9923

Neg Pred Value : 0.9547

Prevalence : 0.7873

Detection Rate : 0.7775

Detection Prevalence : 0.7836

Balanced Accuracy : 0.9796

'Positive' Class : No

***MLeval: Machine Learning Model Evaluation***

Input: caret train function object

Averaging probs.

Group 1 type: repeatedcv

Observations: 3092

Number of groups: 1

Observations per group: 3092

Positive: Yes

Negative: No

Group: Group 1

Positive: 684

Negative: 2408

***Performance Metrics***

Group 1 Optimal Informedness = 0.975032056886402

Group 1 AUC-ROC = 1

**Logistic Regression**

**PCNL_Post_Infection**_logit<-train(**Post_infection** ~., data= **PCNL_Post_Infection**_train1, method="LogitBoost", trControl= **PCNL_Post_Infection**_ctrl)

**PCNL_Post_Infection**_logit_predict<-predict(**PCNL_Post_Infection**_logit, **PCNL_Post_Infection**_test1)

**PCNL_Post_Infection**_logit_tb<-table(pred=factor(**PCNL_Post_Infection**_logit_predict), ref= **PCNL_Post_Infection**_test1$**Post_infection**)

confusionMatrix(**PCNL_Post_Infection**_logit_tb)

res_**PCNL_Post_Infection**_logit<- evalm(**PCNL_Post_Infection**_logit)

Confusion Matrix and Statistics

ref

pred	No Yes

No 1027	15

Yes	17 267

Accuracy : 0.9759

95% CI : (0.9661, 0.9834)

No Information Rate : 0.7873

P-Value [Acc > NIR] : <2e-16

Kappa : 0.9281

Mcnemar's Test P-Value : 0.8597

Sensitivity : 0.9837

Specificity : 0.9468

Pos Pred Value : 0.9856

Neg Pred Value : 0.9401

Prevalence : 0.7873

Detection Rate : 0.7745

Detection Prevalence : 0.7858

Balanced Accuracy : 0.9653

'Positive' Class : No

***MLeval: Machine Learning Model Evaluation***

Input: caret train function object

Averaging probs.

Group 1 type: repeatedcv

Observations: 3092

Number of groups: 1

Observations per group: 3092

Positive: Yes

Negative: No

Group: Group 1

Positive: 684

Negative: 2408

***Performance Metrics***

Group 1 Optimal Informedness = 0.943067455460356

Group 1 AUC-ROC = 0.98

**Neural Net**

**PCNL_Post_Infection**_nn<-train(**Post_infection** ~., data= **PCNL_Post_Infection**_train1, method="nnet", trControl= **PCNL_Post_Infection**_ctrl)

**PCNL_Post_Infection**_nn_predict<-predict(**PCNL_Post_Infection**_nn, **PCNL_Post_Infection**_test1)

**PCNL_Post_Infection**_nn_tb<-table(pred=factor(**PCNL_Post_Infection**_nn_predict), ref= **PCNL_Post_Infection**_test1$**Post_infection**)

confusionMatrix(**PCNL_Post_Infection**_nn_tb)

res_**PCNL_Post_Infection**_nn<- evalm(**PCNL_Post_Infection**_nn)

Confusion Matrix and Statistics

ref

pred	No Yes

No 1027	16

Yes	17 266

Accuracy : 0.9751

95% CI : (0.9652, 0.9828)

No Information Rate : 0.7873

P-Value [Acc > NIR] : <2e-16

Kappa : 0.9258

Mcnemar's Test P-Value : 1

Sensitivity : 0.9837

Specificity : 0.9433

Pos Pred Value : 0.9847

Neg Pred Value : 0.9399

Prevalence : 0.7873

Detection Rate : 0.7745

Detection Prevalence : 0.7866

Balanced Accuracy : 0.9635

'Positive' Class : No

***MLeval: Machine Learning Model Evaluation***

Input: caret train function object

Averaging probs.

Group 1 type: repeatedcv

Observations: 3092

Number of groups: 1

Observations per group: 3092

Positive: Yes

Negative: No

Group: Group 1

Positive: 684

Negative: 2408

***Performance Metrics***

Group 1 Optimal Informedness = 0.928947854131453

Group 1 AUC-ROC = 0.98

**Bayesian Generalised Linear Model**

**PCNL_Post_Infection**_bayes<-train(**Post_infection** ~., data= **PCNL_Post_Infection**_train1, method="bayesglm", trControl= **PCNL_Post_Infection**_ctrl)

**PCNL_Post_Infection**_bayes_predict<-predict(**PCNL_Post_Infection**_bayes, **PCNL_Post_Infection**_test1)

**PCNL_Post_Infection**_bayes_tb<-table(pred=factor(**PCNL_Post_Infection**_bayes_predict), ref= **PCNL_Post_Infection**_test1$**Post_infection**)

confusionMatrix(**PCNL_Post_Infection**_bayes_tb)

res_**PCNL_Post_Infection**_bayes<- evalm(**PCNL_Post_Infection**_bayes)

Confusion Matrix and Statistics

ref

pred	No Yes

No 1024	40

Yes	20 242

Accuracy : 0.9548

95% CI : (0.9421, 0.9653)

No Information Rate : 0.7873

P-Value [Acc > NIR] : < 2e-16

Kappa : 0.8613

Mcnemar's Test P-Value : 0.01417

Sensitivity : 0.9808

Specificity : 0.8582

Pos Pred Value : 0.9624

Neg Pred Value : 0.9237

Prevalence : 0.7873

Detection Rate : 0.7722

Detection Prevalence : 0.8024

Balanced Accuracy : 0.9195

'Positive' Class : No

***MLeval: Machine Learning Model Evaluation***

Input: caret train function object

Averaging probs.

Group 1 type: repeatedcv

Observations: 3092

Number of groups: 1

Observations per group: 3092

Positive: Yes

Negative: No

Group: Group 1

Positive: 684

Negative: 2408

***Performance Metrics***

Group 1 Optimal Informedness = 0.924447747275165

Group 1 AUC-ROC = 0.97

**Generate ROC curves**

res_**File_Name**_rf<-evalm(**File_Name**_rf)

res_**File_Name**_rpart<- evalm(**File_Name**_rpart)

res_**File_Name**_xgboost<- evalm(**File_Name**_xgboost)

res_**File_Name**_logit<- evalm(**File_Name**_logit)

res_**File_Name**_nn<- evalm(**File_Name**_nn)

res_**File_Name**_bayes<- evalm(**File_Name**_bayes)

res_**File_Name**_rf2$roc

res_**File_Name**_rpart$roc

res_**File_Name**_xgboost$roc

res_**File_Name**_logit$roc

res_**File_Name**_nn$roc

res_**File_Name**_bayes$roc

res_**PCNL_Post_Infection**<-evalm(list(**PCNL_Post_Infection**_rf, **PCNL_Post_Infection**_rpart, **PCNL_Post_Infection**_xgboost, **PCNL_Post_Infection**_logit, **PCNL_Post_Infection**_nn, **PCNL_Post_Infection**_bayes), gnames=c('Random Forest', 'Partitioning', 'Extreme Gradient Boosting', 'Logistic Regression', 'Neural Network', 'Bayesian Generalised Linear Model'))

**#Deep Neural Net**

PCNL_Post_Infection_train_predictors3<-as.matrix(PCNL_Post_Infection_train_predictors1)

PCNL_Post_Infection_train_outcome<-ifelse(PCNL_Post_Infection_train1$Post_infection=="Yes", 1,0)

PCNL_Post_Infection_test_predictors3<-as.matrix(PCNL_Post_Infection_test_predictors1)

PCNL_Post_Infection_test_outcome<-ifelse(PCNL_Post_Infection_test1$Post_infection=="Yes", 1,0)

library(keras)

library(tensorflow)

library(pROC)

Post_Infection_keras<-keras_model_sequential()

Post_Infection_keras %>%

layer_dense(units = 32, input_shape = c(43)) %>%

layer_activation('relu') %>%

layer_dense(units = 1) %>%

layer_activation('sigmoid')

Post_Infection_keras %>% compile(loss='binary_crossentropy', optimizer='rmsprop', metrics='accuracy')

Post_Infection_keras %>% fit(PCNL_Post_Infection_train_predictors3, PCNL_Post_Infection_train_outcome, epochs=200, batch_size=5, validation_split=0.2, verbose=2)

Post_Infection_keras_results<-Post_Infection_keras %>% evaluate(PCNL_Post_Infection_test_predictors3, PCNL_Post_Infection_test_outcome, verbose=1)

Post_Infection_keras_prediction<- Post_Infection_keras%>%predict(PCNL_Post_Infection_test_predictors3, batch_size=32, verbose=1)

Post_Infection_keras_prediction1<-round(Post_Infection_keras_prediction, digits=0)

Post_Infection_keras_tb<- table(PCNL_Post_Infection_test_outcome, Post_Infection_keras_prediction1)

Post_Infection_keras_roc<- roc(PCNL_Post_Infection_test_outcome, Post_Infection_keras_prediction1)

auc(Post_Infection_keras_roc)

confusionMatrix(Post_Infection_keras_tb)

plot(Post_Infection_keras_roc)

```
Area under the curve: 0.924
Confusion Matrix and Statistics

                                Post_Infection_keras_prediction1
PCNL_Post_Infection_test_outcome    0    1
                               0 1026   18
                               1   38  244

               Accuracy : 0.9578
                 95% CI : (0.9455, 0.9679)
    No Information Rate : 0.8024
    P-Value [Acc > NIR] : < 2e-16

                  Kappa : 0.8705

 Mcnemar's Test P-Value : 0.01112

            Sensitivity : 0.9643
            Specificity : 0.9313
         Pos Pred Value : 0.9828
         Neg Pred Value : 0.8652
             Prevalence : 0.8024
         Detection Rate : 0.7738
   Detection Prevalence : 0.7873
      Balanced Accuracy : 0.9478

       'Positive' Class : 0
```

9. **Intra-operative Complications**

**#Create training/test sets:**

**PCNL_Intra_Complication**_1<-na.omit(**PCNL_Intra_Complication**_coded)

**PCNL_Intra_Complication**_2<-na.omit(**PCNL_Intra_Complication**_non_coded)

set.seed(1234)

**PCNL_Intra_Complication**_sample_coded<-sample(1:nrow(**PCNL_Intra_Complication**_1), size=nrow(**PCNL_Intra_Complication**_1) *0.7)

**PCNL_Intra_Complication**_train1<- **PCNL_Intra_Complication**_1[**PCNL_Intra_Complication**_sample_coded,]

**PCNL_Intra_Complication**_test1<- **PCNL_Intra_Complication**_1[-**PCNL_Intra_Complication**_sample_coded,]

**PCNL_Intra_Complication**_train_predictors1<- **PCNL_Intra_Complication**_train1[,-44]

**PCNL_Intra_Complication**_test_predictors1<- **PCNL_Intra_Complication**_test1[,-44]

**PCNL_Intra_Complication**_sample_non_coded<- sample(1:nrow(**PCNL_Intra_Complication**_2), size=nrow(**PCNL_Intra_Complication**_2) *0.7)

**PCNL_Intra_Complication**_train2<- **PCNL_Intra_Complication**_2[**PCNL_Intra_Complication**_sample_non_coded,]

**PCNL_Intra_Complication**_test2<- **PCNL_Intra_Complication**_2[-**PCNL_Intra_Complication**_sample_non_coded,]

**PCNL_Intra_Complication**_train_predictors2<- **PCNL_Intra_Complication**_train2[,-44]

**PCNL_Intra_Complication**_test_predictors2<- **PCNL_Intra_Complication**_test2[,-44]

**PCNL_Intra_Complication**_ctrl<-trainControl(method= "repeatedcv", number=10, classProbs=TRUE, savePredictions=TRUE)

**Random Forests**

**PCNL_Intra_Complication**_rf<- train(**Intra_complications** ~., data= **PCNL_Intra_Complication**_train1, method="cforest", trControl= **PCNL_Intra_Complication**_ctrl)

**PCNL_Intra_Complication**_rf_predict<-predict(**PCNL_Intra_Complication**_rf, newdata= **PCNL_Intra_Complication**_test_predictors1)

**PCNL_Intra_Complication**_rf_tb<-table(**PCNL_Intra_Complication**_rf_predict, ref= **PCNL_Intra_Complication**_test1$**Intra_complications**)

confusionMatrix(**PCNL_Intra_Complication**_rf_tb)

res_**PCNL_Intra_Complication**_rf<-evalm(**PCNL_Intra_Complication**_rf)

Confusion Matrix and Statistics

ref

PCNL_Intra_Complication_rf_predict	No Yes

No 1171	47

Yes	0	0

Accuracy : 0.9614

95% CI : (0.949, 0.9715)

No Information Rate : 0.9614

P-Value [Acc > NIR] : 0.5387

Kappa : 0

Mcnemar's Test P-Value : 1.949e-11

Sensitivity : 1.0000

Specificity : 0.0000

Pos Pred Value : 0.9614

Neg Pred Value : NaN

Prevalence : 0.9614

Detection Rate : 0.9614

Detection Prevalence : 1.0000

Balanced Accuracy : 0.5000

'Positive' Class : No

***MLeval: Machine Learning Model Evaluation***

Input: caret train function object

Averaging probs.

Group 1 type: repeatedcv

Observations: 2840

Number of groups: 1

Observations per group: 2840

Positive: Yes

Negative: No

Group: Group 1

Positive: 93

Negative: 2747

***Performance Metrics***

Group 1 Optimal Informedness = 0.343545842776675

Group 1 AUC-ROC = 0.72

**Partitioning**

**PCNL_Intra_Complication**_rpart<-train(**Intra_complications** ~ ., data= **PCNL_Intra_Complication**_train1, method="rpart", trControl= **PCNL_Intra_Complication**_ctrl)

**PCNL_Intra_Complication**_rpart_predict<-predict(**PCNL_Intra_Complication**_rpart, newdata= **PCNL_Intra_Complication**_test1)

**PCNL_Intra_Complication**_rpart_tb<-table(pred=factor(**PCNL_Intra_Complication**_rpart_predict), ref= **PCNL_Intra_Complication**_test1$**Intra_complications**)

confusionMatrix(**PCNL_Intra_Complication**_rpart_tb)

res_**PCNL_Intra_Complication**_rpart<- evalm(**PCNL_Intra_Complication**_rpart)

Confusion Matrix and Statistics

No Yes

No 1171	47

Yes	0	0

Accuracy : 0.9614

95% CI : (0.949, 0.9715)

No Information Rate : 0.9614

P-Value [Acc > NIR] : 0.5387

Kappa : 0

Mcnemar's Test P-Value : 1.949e-11

Sensitivity : 1.0000

Specificity : 0.0000

Pos Pred Value : 0.9614

Neg Pred Value : NaN

Prevalence : 0.9614

Detection Rate : 0.9614

Detection Prevalence : 1.0000

Balanced Accuracy : 0.5000

'Positive' Class : No

***MLeval: Machine Learning Model Evaluation***

Input: caret train function object

Averaging probs.

Group 1 type: repeatedcv

Observations: 2840

Number of groups: 1

Observations per group: 2840

Positive: Yes

Negative: No

Group: Group 1

Positive: 93

Negative: 2747

***Performance Metrics***

Group 1 Optimal Informedness = 0.114525719161862

Group 1 AUC-ROC = 0.56

**XGBoost**

**PCNL_Intra_Complication**_xgboost<-train(**Intra_complications** ~., data= **PCNL_Intra_Complication**_train1, method="xgbTree", trControl= **PCNL_Intra_Complication**_ctrl)

**PCNL_Intra_Complication**_xgboost_predict<-predict(**PCNL_Intra_Complication**_xgboost, **PCNL_Intra_Complication**_test1)

**PCNL_Intra_Complication**_xgboost_tb<-table(pred=factor(**PCNL_Intra_Complication**_xgboost_predict), ref= **PCNL_Intra_Complication**_test1$**Intra_complications**)

confusionMatrix(**PCNL_Intra_Complication**_xgboost_tb)

res_**PCNL_Intra_Complication**_xgboost<- evalm(**PCNL_Intra_Complication**_xgboost)

Confusion Matrix and Statistics

No Yes

No 1171	47

Yes	0	0

Accuracy : 0.9614

95% CI : (0.949, 0.9715)

No Information Rate : 0.9614

P-Value [Acc > NIR] : 0.5387

Kappa : 0

Mcnemar's Test P-Value : 1.949e-11

Sensitivity : 1.0000

Specificity : 0.0000

Pos Pred Value : 0.9614

Neg Pred Value : NaN

Prevalence : 0.9614

Detection Rate : 0.9614

Detection Prevalence : 1.0000

Balanced Accuracy : 0.5000

'Positive' Class : No

***MLeval: Machine Learning Model Evaluation***

Input: caret train function object

Averaging probs.

Group 1 type: repeatedcv

Observations: 2840

Number of groups: 1

Observations per group: 2840

Positive: Yes

Negative: No

Group: Group 1

Positive: 93

Negative: 2747

***Performance Metrics***

Group 1 Optimal Informedness = 0.315401748143625

Group 1 AUC-ROC = 0.69

**Logistic Regression**

**PCNL_Intra_Complication**_logit<-train(**Intra_complications** ~., data= **PCNL_Intra_Complication**_train1, method="LogitBoost", trControl= **PCNL_Intra_Complication**_ctrl)

**PCNL_Intra_Complication**_logit_predict<-predict(**PCNL_Intra_Complication**_logit, **PCNL_Intra_Complication**_test1)

**PCNL_Intra_Complication**_logit_tb<-table(pred=factor(**PCNL_Intra_Complication**_logit_predict), ref= **PCNL_Intra_Complication**_test1$**Intra_complications**)

confusionMatrix(**PCNL_Intra_Complication**_logit_tb)

res_**PCNL_Intra_Complication**_logit<- evalm(**PCNL_Intra_Complication**_logit)

Confusion Matrix and Statistics

ref

pred	No Yes

No 1169	45

Yes	2	2

Accuracy : 0.9614

95% CI : (0.949, 0.9715)

No Information Rate : 0.9614

P-Value [Acc > NIR] : 0.5387

Kappa : 0.0728

Mcnemar's Test P-Value : 8.993e-10

Sensitivity : 0.99829

Specificity : 0.04255

Pos Pred Value : 0.96293

Neg Pred Value : 0.50000

Prevalence : 0.96141

Detection Rate : 0.95977

Detection Prevalence : 0.99672

Balanced Accuracy : 0.52042

'Positive' Class : No

***MLeval: Machine Learning Model Evaluation***

Input: caret train function object

Averaging probs.

Group 1 type: repeatedcv

Observations: 2840

Number of groups: 1

Observations per group: 2840

Positive: Yes

Negative: No

Group: Group 1

Positive: 93

Negative: 2747

***Performance Metrics***

Group 1 Optimal Informedness = 0.102919705172016

Group 1 AUC-ROC = 0.58

**Neural Net**

**PCNL_Intra_Complication**_nn<-train(**Intra_complications** ~., data= **PCNL_Intra_Complication**_train1, method="nnet", trControl= **PCNL_Intra_Complication**_ctrl)

**PCNL_Intra_Complication**_nn_predict<-predict(**PCNL_Intra_Complication**_nn, **PCNL_Intra_Complication**_test1)

**PCNL_Intra_Complication**_nn_tb<-table(pred=factor(**PCNL_Intra_Complication**_nn_predict), ref= **PCNL_Intra_Complication**_test1$**Intra_complications**)

confusionMatrix(**PCNL_Intra_Complication**_nn_tb)

res_**PCNL_Intra_Complication**_nn<- evalm(**PCNL_Intra_Complication**_nn)

Confusion Matrix and Statistics

No Yes

No 1171	47

Yes	0	0

Accuracy : 0.9614

95% CI : (0.949, 0.9715)

No Information Rate : 0.9614

P-Value [Acc > NIR] : 0.5387

Kappa : 0

Mcnemar's Test P-Value : 1.949e-11

Sensitivity : 1.0000

Specificity : 0.0000

Pos Pred Value : 0.9614

Neg Pred Value : NaN

Prevalence : 0.9614

Detection Rate : 0.9614

Detection Prevalence : 1.0000

Balanced Accuracy : 0.5000

'Positive' Class : No

***MLeval: Machine Learning Model Evaluation***

Input: caret train function object

Averaging probs.

Group 1 type: repeatedcv

Observations: 2840

Number of groups: 1

Observations per group: 2840

Positive: Yes

Negative: No

Group: Group 1

Positive: 93

Negative: 2747

***Performance Metrics***

Group 1 Optimal Informedness = 0.169788351711153

Group 1 AUC-ROC = 0.6

**Bayesian Generalised Linear Model**

**PCNL_Intra_Complication**_bayes<-train(**Intra_complications** ~., data= **PCNL_Intra_Complication**_train1, method="bayesglm", trControl= **PCNL_Intra_Complication**_ctrl)

**PCNL_Intra_Complication**_bayes_predict<-predict(**PCNL_Intra_Complication**_bayes, **PCNL_Intra_Complication**_test1)

**PCNL_Intra_Complication**_bayes_tb<-table(pred=factor(**PCNL_Intra_Complication**_bayes_predict), ref= **PCNL_Intra_Complication**_test1$**Intra_complications**)

confusionMatrix(**PCNL_Intra_Complication**_bayes_tb)

res_**PCNL_Intra_Complication**_bayes<- evalm(**PCNL_Intra_Complication**_bayes)

Confusion Matrix and Statistics

ref

pred	No Yes

No 1171	46

Yes	0	1

Accuracy : 0.9622

95% CI : (0.9499, 0.9722)

No Information Rate : 0.9614

P-Value [Acc > NIR] : 0.4795

Kappa : 0.0401

Mcnemar's Test P-Value : 3.247e-11

Sensitivity : 1.00000

Specificity : 0.02128

Pos Pred Value : 0.96220

Neg Pred Value : 1.00000

Prevalence : 0.96141

Detection Rate : 0.96141

Detection Prevalence : 0.99918

Balanced Accuracy : 0.51064

'Positive' Class : No

***MLeval: Machine Learning Model Evaluation***

Input: caret train function object

Averaging probs.

Group 1 type: repeatedcv

Observations: 2840

Number of groups: 1

Observations per group: 2840

Positive: Yes

Negative: No

Group: Group 1

Positive: 93

Negative: 2747

***Performance Metrics***

Group 1 Optimal Informedness = 0.387366863557899

Group 1 AUC-ROC = 0.72

**Generate ROC curves**

res_**PCNL_Intra_Complication**<-evalm(list(**PCNL_Intra_Complication**_rf, **PCNL_Intra_Complication**_rpart, **PCNL_Intra_Complication**_xgboost, **PCNL_Intra_Complication**_logit, **PCNL_Intra_Complication**_nn, **PCNL_Intra_Complication**_bayes), gnames=c('Random Forest', 'Partitioning', 'Extreme Gradient Boosting', 'Logistic Regression', 'Neural Network', 'Bayesian Generalised Linear Model'))

**#Deep Neural Net**

PCNL_Intra_Complication_train_predictors3<-as.matrix(PCNL_Intra_Complication_train_predictors1)

PCNL_Intra_Complication_train_outcome<-ifelse(PCNL_Intra_Complication_train1$Intra_complications=="Yes", 1,0)

PCNL_Intra_Complication_test_predictors3<-as.matrix(PCNL_Intra_Complication_test_predictors1)

PCNL_Intra_Complication_test_outcome<-ifelse(PCNL_Intra_Complication_test1$Intra_complications=="Yes", 1,0)

library(keras)

library(tensorflow)

library(pROC)

Intra_Complication_keras<-keras_model_sequential()

Intra_Complication_keras %>%

layer_dense(units = 32, input_shape = c(43)) %>%

layer_activation('relu') %>%

layer_dense(units = 1) %>%

layer_activation('sigmoid')

Intra_Complication_keras %>% compile(loss='binary_crossentropy', optimizer='rmsprop', metrics='accuracy')

Intra_Complication_keras %>% fit(PCNL_Intra_Complication_train_predictors3, PCNL_Intra_Complication_train_outcome, epochs=200, batch_size=5, validation_split=0.2, verbose=2)

Intra_Complication_keras_results<-Intra_Complication_keras %>% evaluate(PCNL_Intra_Complication_test_predictors3, PCNL_Intra_Complication_test_outcome, verbose=1)

Intra_Complication_keras_prediction<- Intra_Complication_keras%>%predict(PCNL_Intra_Complication_test_predictors3, batch_size=32, verbose=1)

Intra_Complicationn_keras_prediction1<-round(Intra_Complication_keras_prediction, digits=0)

Intra_Complication_keras_tb<- table(PCNL_Intra_Complication_test_outcome, Intra_Complication_keras_prediction1)

Intra_Complication_keras_roc<- roc(PCNL_Intra_Complication_test_outcome, Intra_Complication_keras_prediction1)

auc(Intra_Complication_keras_roc)

confusionMatrix(Intra_Complication_keras_tb)

plot(Intra_Complication_keras_roc)

```
Area under the curve: 0.5
Confusion Matrix and Statistics

     0    1
0 1171    0
1   47    0

               Accuracy : 0.9614
                 95% CI : (0.949, 0.9715)
    No Information Rate : 1
    P-Value [Acc > NIR] : 1

                  Kappa : 0

 Mcnemar's Test P-Value : 1.949e-11

            Sensitivity : 0.9614
            Specificity :     NA
         Pos Pred Value :     NA
         Neg Pred Value :     NA
             Prevalence : 1.0000
         Detection Rate : 0.9614
   Detection Prevalence : 0.9614
      Balanced Accuracy :     NA

       'Positive' Class : 0
```

10. **Need for ITU/HDU**

**#Create training/test sets:**

**PCNL_ITU_HDU**_1<-na.omit(**PCNL_ITU_HDU**_coded)

**PCNL_ITU_HDU**_2<-na.omit(**PCNL_ITU_HDU**_non_coded)

set.seed(1234)

**PCNL_ITU_HDU**_sample_coded<-sample(1:nrow(**PCNL_ITU_HDU**_1), size=nrow(**PCNL_ITU_HDU**_1) *0.7)

**PCNL_ITU_HDU**_train1<- **PCNL_ITU_HDU**_1[**PCNL_ITU_HDU**_sample_coded,]

**PCNL_ITU_HDU**_test1<- **PCNL_ITU_HDU**_1[-**PCNL_ITU_HDU**_sample_coded,]

**PCNL_ITU_HDU**_train_predictors1<- **PCNL_ITU_HDU**_train1[,-44]

**PCNL_ITU_HDU**_test_predictors1<- **PCNL_ITU_HDU**_test1[,-44]

**PCNL_ITU_HDU**_sample_non_coded<- sample(1:nrow(**PCNL_ITU_HDU**_2), size=nrow(**PCNL_ITU_HDU**_2) *0.7)

**PCNL_ITU_HDU**_train2<- **PCNL_ITU_HDU**_2[**PCNL_ITU_HDU**_sample_non_coded,]

**PCNL_ITU_HDU**_test2<- **PCNL_ITU_HDU**_2[-**PCNL_ITU_HDU**_sample_non_coded,]

**PCNL_ITU_HDU**_train_predictors2<- **PCNL_ITU_HDU**_train2[,-44]

**PCNL_ITU_HDU**_test_predictors2<- **PCNL_ITU_HDU**_test2[,-44]

**PCNL_ITU_HDU**_ctrl<-trainControl(method= "repeatedcv", number=10, classProbs=TRUE, savePredictions=TRUE)

**Random Forests**

**PCNL_ITU_HDU**_rf<- train(**ITU_HDU** ~., data= **PCNL_ITU_HDU**_train1, method="cforest", trControl= **PCNL_ITU_HDU**_ctrl)

**PCNL_ITU_HDU**_rf_predict<-predict(**PCNL_ITU_HDU**_rf, newdata= **PCNL_ITU_HDU**_test_predictors1)

**PCNL_ITU_HDU**_rf_tb<-table(**PCNL_ITU_HDU**_rf_predict, ref= **PCNL_ITU_HDU**_test1$**ITU_HDU**)

confusionMatrix(**PCNL_ITU_HDU**_rf_tb)

res_**PCNL_ITU_HDU**_rf<-evalm(**PCNL_ITU_HDU**_rf)

Confusion Matrix and Statistics

ref

PCNL_ITU_HDU_rf_predict	No Yes

No 1187	73

Yes	0	0

Accuracy : 0.9421

95% CI : (0.9277, 0.9543)

No Information Rate : 0.9421

P-Value [Acc > NIR] : 0.5311

Kappa : 0

Mcnemar's Test P-Value : <2e-16

Sensitivity : 1.0000

Specificity : 0.0000

Pos Pred Value : 0.9421

Neg Pred Value : NaN

Prevalence : 0.9421

Detection Rate : 0.9421

Detection Prevalence : 1.0000

Balanced Accuracy : 0.5000

'Positive' Class : No

***MLeval: Machine Learning Model Evaluation***

Input: caret train function object

Averaging probs.

Group 1 type: repeatedcv

Observations: 2940

Number of groups: 1

Observations per group: 2940

Positive: Yes

Negative: No

Group: Group 1

Positive: 128

Negative: 2812

***Performance Metrics***

Group 1 Optimal Informedness = 0.245288051209104

Group 1 AUC-ROC = 0.63

**Partitioning**

**PCNL_ITU_HDU**_rpart<-train(**ITU_HDU** ~ ., data= **PCNL_ITU_HDU**_train1, method="rpart", trControl= **PCNL_ITU_HDU**_ctrl)

**PCNL_ITU_HDU**_rpart_predict<-predict(**PCNL_ITU_HDU**_rpart, newdata= **PCNL_ITU_HDU**_test1)

**PCNL_ITU_HDU**_rpart_tb<-table(pred=factor(**PCNL_ITU_HDU**_rpart_predict), ref= **PCNL_ITU_HDU**_test1$**ITU_HDU**)

confusionMatrix(**PCNL_ITU_HDU**_rpart_tb)

res_**PCNL_ITU_HDU**_rpart<- evalm(**PCNL_ITU_HDU**_rpart)

ref

PCNL_ITU_HDU_rpart_predict	No Yes

No 1187	73

Yes	0	0

Accuracy : 0.9421

95% CI : (0.9277, 0.9543)

No Information Rate : 0.9421

P-Value [Acc > NIR] : 0.5311

Kappa : 0

Mcnemar's Test P-Value : <2e-16

Sensitivity : 1.0000

Specificity : 0.0000

Pos Pred Value : 0.9421

Neg Pred Value : NaN

Prevalence : 0.9421

Detection Rate : 0.9421

Detection Prevalence : 1.0000

Balanced Accuracy : 0.5000

'Positive' Class : No

***MLeval: Machine Learning Model Evaluation***

Input: caret train function object

Averaging probs.

Group 1 type: repeatedcv

Observations: 2940

Number of groups: 1

Observations per group: 2940

Positive: Yes

Negative: No

Group: Group 1

Positive: 128

Negative: 2812

***Performance Metrics***

Group 1 Optimal Informedness = 0.0575213371266003

Group 1 AUC-ROC = 0.54

**XGBoost**

**PCNL_ITU_HDU**_xgboost<-train(**ITU_HDU** ~., data= **PCNL_ITU_HDU**_train1, method="xgbTree", trControl= **PCNL_ITU_HDU**_ctrl)

**PCNL_ITU_HDU**_xgboost_predict<-predict(**PCNL_ITU_HDU**_xgboost, **PCNL_ITU_HDU**_test1)

**PCNL_ITU_HDU**_xgboost_tb<-table(pred=factor(**PCNL_ITU_HDU**_xgboost_predict), ref= **PCNL_ITU_HDU**_test1$**ITU_HDU**)

confusionMatrix(**PCNL_ITU_HDU**_xgboost_tb)

res_**PCNL_ITU_HDU**_xgboost<- evalm(**PCNL_ITU_HDU**_xgboost)

PCNL_ITU_HDU_xgboost_predict	No Yes

No 1187	73

Yes	0	0

Accuracy : 0.9421

95% CI : (0.9277, 0.9543)

No Information Rate : 0.9421

P-Value [Acc > NIR] : 0.5311

Kappa : 0

Mcnemar's Test P-Value : <2e-16

Sensitivity : 1.0000

Specificity : 0.0000

Pos Pred Value : 0.9421

Neg Pred Value : NaN

Prevalence : 0.9421

Detection Rate : 0.9421

Detection Prevalence : 1.0000

Balanced Accuracy : 0.5000

'Positive' Class : No

***MLeval: Machine Learning Model Evaluation***

Input: caret train function object

Averaging probs.

Group 1 type: repeatedcv

Observations: 2940

Number of groups: 1

Observations per group: 2940

Positive: Yes

Negative: No

Group: Group 1

Positive: 128

Negative: 2812

***Performance Metrics***

Group 1 Optimal Informedness = 0.176987019914651

Group 1 AUC-ROC = 0.6

**Logistic Regression**

**PCNL_ITU_HDU**_logit<-train(**ITU_HDU** ~., data= **PCNL_ITU_HDU**_train1, method="LogitBoost", trControl= **PCNL_ITU_HDU**_ctrl)

**PCNL_ITU_HDU**_logit_predict<-predict(**PCNL_ITU_HDU**_logit, **PCNL_ITU_HDU**_test1)

**PCNL_ITU_HDU**_logit_tb<-table(pred=factor(**PCNL_ITU_HDU**_logit_predict), ref= **PCNL_ITU_HDU**_test1$**ITU_HDU**)

confusionMatrix(**PCNL_ITU_HDU**_logit_tb)

res_**PCNL_ITU_HDU**_logit<- evalm(**PCNL_ITU_HDU**_logit)

***MLeval: Machine Learning Model Evaluation***

Input: caret train function object

Averaging probs.

Group 1 type: repeatedcv

Observations: 2940

Number of groups: 1

Observations per group: 2940

Positive: Yes

Negative: No

Group: Group 1

Positive: 128

Negative: 2812

***Performance Metrics***

Group 1 Optimal Informedness = 0.0497866287339972

Group 1 AUC-ROC = 0.52

Confusion Matrix and Statistics

ref

pred	No Yes

No 1186	73

Yes	1	0

Accuracy : 0.9413

95% CI : (0.9268, 0.9536)

No Information Rate : 0.9421

P-Value [Acc > NIR] : 0.5785

Kappa : -0.0016

Mcnemar's Test P-Value : <2e-16

Sensitivity : 0.9992

Specificity : 0.0000

Pos Pred Value : 0.9420

Neg Pred Value : 0.0000

Prevalence : 0.9421

Detection Rate : 0.9413

Detection Prevalence : 0.9992

Balanced Accuracy : 0.4996

'Positive' Class : No

**Neural Net**

**PCNL_ITU_HDU**_nn<-train(**ITU_HDU** ~., data= **PCNL_ITU_HDU**_train1, method="nnet", trControl= **PCNL_ITU_HDU**_ctrl)

**PCNL_ITU_HDU**_nn_predict<-predict(**PCNL_ITU_HDU**_nn, **PCNL_ITU_HDU**_test1)

**PCNL_ITU_HDU**_nn_tb<-table(pred=factor(**PCNL_ITU_HDU**_nn_predict), ref= **PCNL_ITU_HDU**_test1$**ITU_HDU**)

confusionMatrix(**PCNL_ITU_HDU**_nn_tb)

res_**PCNL_ITU_HDU**_nn<- evalm(**PCNL_ITU_HDU**_nn)

PCNL_ITU_HDU_nn_predict	No Yes

No 1187	73

Yes	0	0

Accuracy : 0.9421

95% CI : (0.9277, 0.9543)

No Information Rate : 0.9421

P-Value [Acc > NIR] : 0.5311

Kappa : 0

Mcnemar's Test P-Value : <2e-16

Sensitivity : 1.0000

Specificity : 0.0000

Pos Pred Value : 0.9421

Neg Pred Value : NaN

Prevalence : 0.9421

Detection Rate : 0.9421

Detection Prevalence : 1.0000

Balanced Accuracy : 0.5000

'Positive' Class : No

***MLeval: Machine Learning Model Evaluation***

Input: caret train function object

Averaging probs.

Group 1 type: repeatedcv

Observations: 2940

Number of groups: 1

Observations per group: 2940

Positive: Yes

Negative: No

Group: Group 1

Positive: 128

Negative: 2812

***Performance Metrics***

Group 1 Optimal Informedness = 0

Group 1 AUC-ROC = 0.49

**Bayesian Generalised Linear Model**

**PCNL_ITU_HDU**_bayes<-train(**ITU_HDU** ~., data= **PCNL_ITU_HDU**_train1, method="bayesglm", trControl= **PCNL_ITU_HDU**_ctrl)

**PCNL_ITU_HDU**_bayes_predict<-predict(**PCNL_ITU_HDU**_bayes, **PCNL_ITU_HDU**_test1)

**PCNL_ITU_HDU**_bayes_tb<-table(pred=factor(**PCNL_ITU_HDU**_bayes_predict), ref= **PCNL_ITU_HDU**_test1$**ITU_HDU**)

confusionMatrix(**PCNL_ITU_HDU**_bayes_tb)

res_**PCNL_ITU_HDU**_bayes<- evalm(**PCNL_ITU_HDU**_bayes)

PCNL_ITU_HDU_bayes_predict	No Yes

No 1187	73

Yes	0	0

Accuracy : 0.9421

95% CI : (0.9277, 0.9543)

No Information Rate : 0.9421

P-Value [Acc > NIR] : 0.5311

Kappa : 0

Mcnemar's Test P-Value : <2e-16

Sensitivity : 1.0000

Specificity : 0.0000

Pos Pred Value : 0.9421

Neg Pred Value : NaN

Prevalence : 0.9421

Detection Rate : 0.9421

Detection Prevalence : 1.0000

Balanced Accuracy : 0.5000

'Positive' Class : No

***MLeval: Machine Learning Model Evaluation***

Input: caret train function object

Averaging probs.

Group 1 type: repeatedcv

Observations: 2940

Number of groups: 1

Observations per group: 2940

Positive: Yes

Negative: No

Group: Group 1

Positive: 128

Negative: 2812

***Performance Metrics***

Group 1 Optimal Informedness = 0.0784583926031295

Group 1 AUC-ROC = 0.54

**Generate ROC curves**

res_**PCNL_ITU_HDU**<-evalm(list(**PCNL_ITU_HDU**_rf, **PCNL_ITU_HDU**_rpart, **PCNL_ITU_HDU**_xgboost, **PCNL_ITU_HDU**_logit, **PCNL_ITU_HDU**_nn, **PCNL_ITU_HDU**_bayes), gnames=c('Random Forest', 'Partitioning', 'Extreme Gradient Boosting', 'Logistic Regression', 'Neural Network', 'Bayesian Generalised Linear Model'))

**#Deep Neural Net**

PCNL_ITU_HDU_train_predictors3<-as.matrix(PCNL_ITU_HDU_train_predictors1)

PCNL_ITU_HDU_train_outcome<-ifelse(PCNL_ITU_HDU_train1$ITU_HDU=="Yes", 1,0)

PCNL_ITU_HDU_test_predictors3<-as.matrix(PCNL_ITU_HDU_test_predictors1)

PCNL_ITU_HDU_test_outcome<-ifelse(PCNL_ITU_HDU_test1$ITU_HDU=="Yes", 1,0)

library(keras)

library(tensorflow)

library(pROC)

ITU_HDU_keras<-keras_model_sequential()

ITU_HDU_keras %>%

layer_dense(units = 32, input_shape = c(43)) %>%

layer_activation('relu') %>%

layer_dense(units = 1) %>%

layer_activation('sigmoid')

ITU_HDU_keras %>% compile(loss='binary_crossentropy', optimizer='rmsprop', metrics='accuracy')

ITU_HDU_keras %>% fit(PCNL_ITU_HDU_train_predictors3, PCNL_ITU_HDU_train_outcome, epochs=200, batch_size=5, validation_split=0.2, verbose=2)

ITU_HDU_keras_results<-ITU_HDU_keras %>% evaluate(PCNL_ITU_HDU_test_predictors3, PCNL_ITU_HDU_test_outcome, verbose=1)

ITU_HDU_keras_prediction<- ITU_HDU_keras%>%predict(PCNL_ITU_HDU_test_predictors3, batch_size=32, verbose=1)

ITU_HDU_keras_prediction1<-round(ITU_HDU_keras_prediction, digits=0)

ITU_HDU_keras_tb<- table(PCNL_ITU_HDU_test_outcome, ITU_HDU_keras_prediction1)

ITU_HDU_keras_roc<- roc(PCNL_ITU_HDU_test_outcome, ITU_HDU_keras_prediction1)

auc(ITU_HDU_keras_roc)

confusionMatrix(ITU_HDU_keras_tb)

plot(ITU_HDU_keras_roc)

```
Area under the curve: 0.5
Confusion Matrix and Statistics

     0    1
0 1187    0
1   73    0

               Accuracy : 0.9421
                 95% CI : (0.9277, 0.9543)
    No Information Rate : 1
    P-Value [Acc > NIR] : 1

                  Kappa : 0

 Mcnemar's Test P-Value : <2e-16

            Sensitivity : 0.9421
            Specificity :     NA
         Pos Pred Value :     NA
         Neg Pred Value :     NA
             Prevalence : 1.0000
         Detection Rate : 0.9421
   Detection Prevalence : 0.9421
      Balanced Accuracy :     NA

       'Positive' Class : 0
```

11. **Stone Free at Follow-up**

**#Create training/test sets:**

**PCNL_SF**_1<-na.omit(**PCNL_SF**_coded)

**PCNL_SF**_2<-na.omit(**PCNL_SF**_non_coded)

set.seed(1234)

**PCNL_SF**_sample_coded<-sample(1:nrow(**PCNL_SF**_1), size=nrow(**PCNL_SF**_1) *0.7)

**PCNL_SF**_train1<- **PCNL_SF**_1[**PCNL_SF**_sample_coded,]

**PCNL_SF**_test1<- **PCNL_SF**_1[-**PCNL_SF**_sample_coded,]

**PCNL_SF**_train_predictors1<- **PCNL_SF**_train1[,-44]

**PCNL_SF**_test_predictors1<- **PCNL_SF**_test1[,-44]

**PCNL_SF**_sample_non_coded<- sample(1:nrow(**PCNL_SF**_2), size=nrow(**PCNL_SF**_2) *0.7)

**PCNL_SF**_train2<- **PCNL_SF**_2[**PCNL_SF**_sample_non_coded,]

**PCNL_SF**_test2<- **PCNL_SF**_2[-**PCNL_SF**_sample_non_coded,]

**PCNL_SF**_train_predictors2<- **PCNL_SF**_train2[,-44]

**PCNL_SF**_test_predictors2<- **PCNL_SF**_test2[,-44]

**PCNL_SF**_ctrl<-trainControl(method= "repeatedcv", number=10, classProbs=TRUE, savePredictions=TRUE)

**Random Forests**

**PCNL_SF**_rf<- train(**SF** ~., data= **PCNL_SF**_train1, method="cforest", trControl= **PCNL_SF**_ctrl)

**PCNL_SF**_rf_predict<-predict(**PCNL_SF**_rf, newdata= **PCNL_SF**_test_predictors1)

**PCNL_SF**_rf_tb<-table(**PCNL_SF**_rf_predict, ref= **PCNL_SF**_test1$**SF**)

confusionMatrix(**PCNL_SF**_rf_tb)

res_**PCNL_SF**_rf<-evalm(**PCNL_SF**_rf)

Confusion Matrix and Statistics

ref

PCNL_SF_rf_predict No Yes

No	0	0

Yes 100 228

Accuracy : 0.6951

95% CI : (0.6422, 0.7445)

No Information Rate : 0.6951

P-Value [Acc > NIR] : 0.527

Kappa : 0

Mcnemar's Test P-Value : <2e-16

Sensitivity : 0.0000

Specificity : 1.0000

Pos Pred Value : NaN

Neg Pred Value : 0.6951

Prevalence : 0.3049

Detection Rate : 0.0000

Detection Prevalence : 0.0000

Balanced Accuracy : 0.5000

'Positive' Class : No

***MLeval: Machine Learning Model Evaluation***

Input: caret train function object

Averaging probs.

Group 1 type: repeatedcv

Observations: 763

Number of groups: 1

Observations per group: 763

Positive: Yes

Negative: No

Group: Group 1

Positive: 535

Negative: 228

***Performance Metrics***

Group 1 Optimal Informedness = 0.288006230529595

Group 1 AUC-ROC = 0.69

**Partitioning**

**PCNL_SF**_rpart<-train(**SF** ~ ., data= **PCNL_SF**_train1, method="rpart", trControl= **PCNL_SF**_ctrl)

**PCNL_SF**_rpart_predict<-predict(**PCNL_SF**_rpart, newdata= **PCNL_SF**_test1)

**PCNL_SF**_rpart_tb<-table(pred=factor(**PCNL_SF**_rpart_predict), ref= **PCNL_SF**_test1$**SF**)

confusionMatrix(**PCNL_SF**_rpart_tb)

res_**PCNL_SF**_rpart<- evalm(**PCNL_SF**_rpart)

Confusion Matrix and Statistics

ref

PCNL_SF_rpart_predict No Yes

No	0	0

Yes 100 228

Accuracy : 0.6951

95% CI : (0.6422, 0.7445)

No Information Rate : 0.6951

P-Value [Acc > NIR] : 0.527

Kappa : 0

Mcnemar's Test P-Value : <2e-16

Sensitivity : 0.0000

Specificity : 1.0000

Pos Pred Value : NaN

Neg Pred Value : 0.6951

Prevalence : 0.3049

Detection Rate : 0.0000

Detection Prevalence : 0.0000

Balanced Accuracy : 0.5000

'Positive' Class : No

***MLeval: Machine Learning Model Evaluation***

Input: caret train function object

Averaging probs.

Group 1 type: repeatedcv

Observations: 763

Number of groups: 1

Observations per group: 763

Positive: Yes

Negative: No

Group: Group 1

Positive: 543

Negative: 220

***Performance Metrics***

Group 1 Optimal Informedness = 0.0839862715553323

Group 1 AUC-ROC = 0.55

**XGBoost**

**PCNL_SF**_xgboost<-train(**SF** ~., data= **PCNL_SF**_train1, method="xgbTree", trControl= **PCNL_SF**_ctrl)

**PCNL_SF**_xgboost_predict<-predict(**PCNL_SF**_xgboost, **PCNL_SF**_test1)

**PCNL_SF**_xgboost_tb<-table(pred=factor(**PCNL_SF**_xgboost_predict), ref= **PCNL_SF**_test1$**SF**)

confusionMatrix(**PCNL_SF**_xgboost_tb)

res_**PCNL_SF**_xgboost<- evalm(**PCNL_SF**_xgboost)

Confusion Matrix and Statistics

ref

pred	No Yes

No	22 28

Yes 86 192

Accuracy : 0.6524

95% CI : (0.5982, 0.7039)

No Information Rate : 0.6707

P-Value [Acc > NIR] : 0.7783

Kappa : 0.0885

Mcnemar's Test P-Value : 9.37e-08

Sensitivity : 0.20370

Specificity : 0.87273

Pos Pred Value : 0.44000

Neg Pred Value : 0.69065

Prevalence : 0.32927

Detection Rate : 0.06707

Detection Prevalence : 0.15244

Balanced Accuracy : 0.53822

'Positive' Class : No

***MLeval: Machine Learning Model Evaluation***

Input: caret train function object

Averaging probs.

Group 1 type: repeatedcv

Observations: 763

Number of groups: 1

Observations per group: 763

Positive: Yes

Negative: No

Group: Group 1

Positive: 543

Negative: 220

***Performance Metrics***

Group 1 Optimal Informedness = 0.307073497404989

Group 1 AUC-ROC = 0.7

**Logistic Regression**

**PCNL_SF**_logit<-train(**SF** ~., data= **PCNL_SF**_train1, method="LogitBoost", trControl= **PCNL_SF**_ctrl)

**PCNL_SF**_logit_predict<-predict(**PCNL_SF**_logit, **PCNL_SF**_test1)

**PCNL_SF**_logit_tb<-table(pred=factor(**PCNL_SF**_logit_predict), ref= **PCNL_SF**_test1$**SF**)

confusionMatrix(**PCNL_SF**_logit_tb)

res_**PCNL_SF**_logit<- evalm(**PCNL_SF**_logit)

Confusion Matrix and Statistics

ref

pred	No Yes

No	32 49

Yes 76 171

Accuracy : 0.6189

95% CI : (0.5639, 0.6717)

No Information Rate : 0.6707

P-Value [Acc > NIR] : 0.97917

Kappa : 0.0786

Mcnemar's Test P-Value : 0.02004

Sensitivity : 0.29630

Specificity : 0.77727

Pos Pred Value : 0.39506

Neg Pred Value : 0.69231

Prevalence : 0.32927

Detection Rate : 0.09756

Detection Prevalence : 0.24695

Balanced Accuracy : 0.53678

'Positive' Class : No

***MLeval: Machine Learning Model Evaluation***

Input: caret train function object

Averaging probs.

Group 1 type: repeatedcv

Observations: 763

Number of groups: 1

Observations per group: 763

Positive: Yes

Negative: No

Group: Group 1

Positive: 543

Negative: 220

***Performance Metrics***

Group 1 Optimal Informedness = 0.174652603381885

Group 1 AUC-ROC = 0.61

**Neural Net**

**PCNL_SF**_nn<-train(**SF** ~., data= **PCNL_SF**_train1, method="nnet", trControl= **PCNL_SF**_ctrl)

**PCNL_SF**_nn_predict<-predict(**PCNL_SF**_nn, **PCNL_SF**_test1)

**PCNL_SF**_nn_tb<-table(pred=factor(**PCNL_SF**_nn_predict), ref= **PCNL_SF**_test1$**SF**)

confusionMatrix(**PCNL_SF**_nn_tb)

res_**PCNL_SF**_nn<- evalm(**PCNL_SF**_nn)

Confusion Matrix and Statistics

ref

PCNL_SF_nn_predict No Yes

No	0	0

Yes 100 228

Accuracy : 0.6951

95% CI : (0.6422, 0.7445)

No Information Rate : 0.6951

P-Value [Acc > NIR] : 0.527

Kappa : 0

Mcnemar's Test P-Value : <2e-16

Sensitivity : 0.0000

Specificity : 1.0000

Pos Pred Value : NaN

Neg Pred Value : 0.6951

Prevalence : 0.3049

Detection Rate : 0.0000

Detection Prevalence : 0.0000

Balanced Accuracy : 0.5000

'Positive' Class : No

***MLeval: Machine Learning Model Evaluation***

Input: caret train function object

Averaging probs.

Group 1 type: repeatedcv

Observations: 763

Number of groups: 1

Observations per group: 763

Positive: Yes

Negative: No

Group: Group 1

Positive: 543

Negative: 220

***Performance Metrics***

Group 1 Optimal Informedness = 0

Group 1 AUC-ROC = 0.5

**Bayesian Generalised Linear Model**

**PCNL_SF**_bayes<-train(**SF** ~., data= **PCNL_SF**_train1, method="bayesglm", trControl= **PCNL_SF**_ctrl)

**PCNL_SF**_bayes_predict<-predict(**PCNL_SF**_bayes, **PCNL_SF**_test1)

**PCNL_SF**_bayes_tb<-table(pred=factor(**PCNL_SF**_bayes_predict), ref= **PCNL_SF**_test1$**SF**)

confusionMatrix(**PCNL_SF**_bayes_tb)

res_**PCNL_SF**_bayes<- evalm(**PCNL_SF**_bayes)

Confusion Matrix and Statistics

ref

pred	No Yes

No	32 27

Yes 76 193

Accuracy : 0.686

95% CI : (0.6327, 0.7358)

No Information Rate : 0.6707

P-Value [Acc > NIR] : 0.3002

Kappa : 0.1962

Mcnemar's Test P-Value : 2.25e-06

Sensitivity : 0.29630

Specificity : 0.87727

Pos Pred Value : 0.54237

Neg Pred Value : 0.71747

Prevalence : 0.32927

Detection Rate : 0.09756

Detection Prevalence : 0.17988

Balanced Accuracy : 0.58678

'Positive' Class : No

***MLeval: Machine Learning Model Evaluation***

Input: caret train function object

Averaging probs.

Group 1 type: repeatedcv

Observations: 763

Number of groups: 1

Observations per group: 763

Positive: Yes

Negative: No

Group: Group 1

Positive: 543

Negative: 220

***Performance Metrics***

Group 1 Optimal Informedness = 0.265134773145823

Group 1 AUC-ROC = 0.67

**Generate ROC curves**

res_**PCNL_SF** <-evalm(list(**PCNL_SF**_rf, **PCNL_SF**_rpart, **PCNL_SF**_xgboost, **PCNL_SF**_logit, **PCNL_SF**_nn, **PCNL_SF**_bayes), gnames=c('Random Forest', 'Partitioning', 'Extreme Gradient Boosting', 'Logistic Regression', 'Neural Network', 'Bayesian Generalised Linear Model'))

#Deep Neural Net

PCNL_SF_train_predictors3<-as.matrix(PCNL_SF_train_predictors1)

PCNL_SF_train_outcome<-ifelse(PCNL_SF_train1$SF=="Yes", 1,0)

PCNL_SF_test_predictors3<-as.matrix(PCNL_SF_test_predictors1)

PCNL_SF_test_outcome<-ifelse(PCNL_SF_test1$SF=="Yes", 1,0)

library(keras)

library(tensorflow)

library(pROC)

SF_keras<-keras_model_sequential()

SF_keras %>%

layer_dense(units = 32, input_shape = c(43)) %>%

layer_activation('relu') %>%

layer_dense(units = 1) %>%

layer_activation('sigmoid')

SF_keras %>% compile(loss='binary_crossentropy', optimizer='rmsprop', metrics='accuracy')

SF_keras %>% fit(PCNL_SF_train_predictors3, PCNL_SF_train_outcome, epochs=200, batch_size=5, validation_split=0.2, verbose=2)

SF_keras_results<-SF_keras %>% evaluate(PCNL_SF_test_predictors3, PCNL_SF_test_outcome, verbose=1)

SF_keras_prediction<- SF_keras%>%predict(PCNL_SF_test_predictors3, batch_size=32, verbose=1)

SF_keras_prediction1<-round(SF_keras_prediction, digits=0)

SF_keras_tb<- table(PCNL_SF_test_outcome, SF_keras_prediction1)

SF_keras_roc<- roc(PCNL_SF_test_outcome, SF_keras_prediction1)

auc(SF_keras_roc)

confusionMatrix(SF_keras_tb)

plot(SF_keras_roc)

```
Area under the curve: 0.6153
Confusion Matrix and Statistics

                    SF_keras_prediction1
PCNL_SF_test_outcome   0   1
                   0  74  34
                   1 100 120

               Accuracy : 0.5915
                 95% CI : (0.5361, 0.6451)
    No Information Rate : 0.5305
    P-Value [Acc > NIR] : 0.01525

                  Kappa : 0.1996

 Mcnemar's Test P-Value : 1.964e-08

            Sensitivity : 0.4253
            Specificity : 0.7792
         Pos Pred Value : 0.6852
         Neg Pred Value : 0.5455
             Prevalence : 0.5305
         Detection Rate : 0.2256
   Detection Prevalence : 0.3293
      Balanced Accuracy : 0.6023

       'Positive' Class : 0
```

12. **Need for Adjuvant Treatment**

**#Create training/test sets:**

**PCNL_Adjuvant**_1<-na.omit(**PCNL_Adjuvant**_coded)

**PCNL_Adjuvant**_2<-na.omit(**PCNL_Adjuvant**_non_coded)

set.seed(1234)

**PCNL_Adjuvant**_sample_coded<-sample(1:nrow(**PCNL_Adjuvant**_1), size=nrow(**PCNL_Adjuvant**_1) *0.7)

**PCNL_Adjuvant**_train1<- **PCNL_Adjuvant**_1[**PCNL_Adjuvant**_sample_coded,]

**PCNL_Adjuvant**_test1<- **PCNL_Adjuvant**_1[-**PCNL_Adjuvant**_sample_coded,]

**PCNL_Adjuvant**_train_predictors1<- **PCNL_Adjuvant**_train1[,-44]

**PCNL_Adjuvant**_test_predictors1<- **PCNL_Adjuvant**_test1[,-44]

**PCNL_Adjuvant**_sample_non_coded<- sample(1:nrow(**PCNL_Adjuvant**_2), size=nrow(**PCNL_Adjuvant**_2) *0.7)

**PCNL_Adjuvant**_train2<- **PCNL_Adjuvant**_2[**PCNL_Adjuvant**_sample_non_coded,]

**PCNL_Adjuvant**_test2<- **PCNL_Adjuvant**_2[-**PCNL_Adjuvant**_sample_non_coded,]

**PCNL_Adjuvant**_train_predictors2<- **PCNL_Adjuvant**_train2[,-44]

**PCNL_Adjuvant**_test_predictors2<- **PCNL_Adjuvant**_test2[,-44]

**PCNL_Adjuvant**_ctrl<-trainControl(method= "repeatedcv", number=10, classProbs=TRUE, savePredictions=TRUE)

**Random Forests**

**PCNL_Adjuvant**_rf<- train(**Adjuvant** ~., data= **PCNL_Adjuvant**_train1, method="cforest", trControl= **PCNL_Adjuvant**_ctrl)

**PCNL_Adjuvant**_rf_predict<-predict(**PCNL_Adjuvant**_rf, newdata= **PCNL_Adjuvant**_test_predictors1)

**PCNL_Adjuvant**_rf_tb<-table(**PCNL_Adjuvant**_rf_predict, ref= **PCNL_Adjuvant**_test1$**Adjuvant**)

confusionMatrix(**PCNL_Adjuvant**_rf_tb)

res_**PCNL_Adjuvant**_rf<-evalm(**PCNL_Adjuvant**_rf)

Confusion Matrix and Statistics

ref

PCNL_Adjuvant_rf_predict No Yes

No 270 58

Yes	2	1

Accuracy : 0.8187

95% CI : (0.7729, 0.8587)

No Information Rate : 0.8218

P-Value [Acc > NIR] : 0.591

Kappa : 0.0153

Mcnemar's Test P-Value : 1.243e-12

Sensitivity : 0.99265

Specificity : 0.01695

Pos Pred Value : 0.82317

Neg Pred Value : 0.33333

Prevalence : 0.82175

Detection Rate : 0.81571

Detection Prevalence : 0.99094

Balanced Accuracy : 0.50480

'Positive' Class : No

***MLeval: Machine Learning Model Evaluation***

Input: caret train function object

Averaging probs.

Group 1 type: repeatedcv

Observations: 771

Number of groups: 1

Observations per group: 771

Positive: Yes

Negative: No

Group: Group 1

Positive: 126

Negative: 645

***Performance Metrics***

Group 1 Optimal Informedness = 0.301624215577704

Group 1 AUC-ROC = 0.69

**Partitioning**

**PCNL_Adjuvant**_rpart<-train(**Adjuvant** ~ ., data= **PCNL_Adjuvant**_train1, method="rpart", trControl= **PCNL_Adjuvant**_ctrl)

**PCNL_Adjuvant**_rpart_predict<-predict(**PCNL_Adjuvant**_rpart, newdata= **PCNL_Adjuvant**_test1)

**PCNL_Adjuvant**_rpart_tb<-table(pred=factor(**PCNL_Adjuvant**_rpart_predict), ref= **PCNL_Adjuvant**_test1$**Adjuvant**)

confusionMatrix(**PCNL_Adjuvant**_rpart_tb)

res_**PCNL_Adjuvant**_rpart<- evalm(**PCNL_Adjuvant**_rpart)

Confusion Matrix and Statistics

ref

pred	No Yes

No 267 52

Yes	5	7

Accuracy : 0.8278

95% CI : (0.7827, 0.8669)

No Information Rate : 0.8218

P-Value [Acc > NIR] : 0.4205

Kappa : 0.1457

Mcnemar's Test P-Value : 1.109e-09

Sensitivity : 0.9816

Specificity : 0.1186

Pos Pred Value : 0.8370

Neg Pred Value : 0.5833

Prevalence : 0.8218

Detection Rate : 0.8066

Detection Prevalence : 0.9637

Balanced Accuracy : 0.5501

'Positive' Class : No

***MLeval: Machine Learning Model Evaluation***

Input: caret train function object

Averaging probs.

Group 1 type: repeatedcv

Observations: 771

Number of groups: 1

Observations per group: 771

Positive: Yes

Negative: No

Group: Group 1

Positive: 126

Negative: 645

***Performance Metrics***

Group 1 Optimal Informedness = 0.201919527500923

Group 1 AUC-ROC = 0.61

**XGBoost**

**PCNL_Adjuvant**_xgboost<-train(**Adjuvant** ~., data= **PCNL_Adjuvant**_train1, method="xgbTree", trControl= **PCNL_Adjuvant**_ctrl)

**PCNL_Adjuvant**_xgboost_predict<-predict(**PCNL_Adjuvant**_xgboost, **PCNL_Adjuvant**_test1)

**PCNL_Adjuvant**_xgboost_tb<-table(pred=factor(**PCNL_Adjuvant**_xgboost_predict), ref= **PCNL_Adjuvant**_test1$**Adjuvant**)

confusionMatrix(**PCNL_Adjuvant**_xgboost_tb)

res_**PCNL_Adjuvant**_xgboost<- evalm(**PCNL_Adjuvant**_xgboost)

Confusion Matrix and Statistics

ref

pred	No Yes

No 271 58

Yes	1	1

Accuracy : 0.8218

95% CI : (0.7762, 0.8615)

No Information Rate : 0.8218

P-Value [Acc > NIR] : 0.5347

Kappa : 0.0213

Mcnemar's Test P-Value : 3.086e-13

Sensitivity : 0.99632

Specificity : 0.01695

Pos Pred Value : 0.82371

Neg Pred Value : 0.50000

Prevalence : 0.82175

Detection Rate : 0.81873

Detection Prevalence : 0.99396

Balanced Accuracy : 0.50664

'Positive' Class : No

***MLeval: Machine Learning Model Evaluation***

Input: caret train function object

Averaging probs.

Group 1 type: repeatedcv

Observations: 771

Number of groups: 1

Observations per group: 771

Positive: Yes

Negative: No

Group: Group 1

Positive: 126

Negative: 645

***Performance Metrics***

Group 1 Optimal Informedness = 0.298560354374308

Group 1 AUC-ROC = 0.67

**Logistic Regression**

**PCNL_Adjuvant**_logit<-train(**Adjuvant** ~., data= **PCNL_Adjuvant**_train1, method="LogitBoost", trControl= **PCNL_Adjuvant**_ctrl)

**PCNL_Adjuvant**_logit_predict<-predict(**PCNL_Adjuvant**_logit, **PCNL_Adjuvant**_test1)

**PCNL_Adjuvant**_logit_tb<-table(pred=factor(**PCNL_Adjuvant**_logit_predict), ref= **PCNL_Adjuvant**_test1$**Adjuvant**)

confusionMatrix(**PCNL_Adjuvant**_logit_tb)

res_**PCNL_Adjuvant**_logit<- evalm(**PCNL_Adjuvant**_logit)

Confusion Matrix and Statistics

ref

pred	No Yes

No 255 51

Yes 17	8

Accuracy : 0.7946

95% CI : (0.747, 0.8368)

No Information Rate : 0.8218

P-Value [Acc > NIR] : 0.9119

Kappa : 0.0944

Mcnemar's Test P-Value : 6.285e-05

Sensitivity : 0.9375

Specificity : 0.1356

Pos Pred Value : 0.8333

Neg Pred Value : 0.3200

Prevalence : 0.8218

Detection Rate : 0.7704

Detection Prevalence : 0.9245

Balanced Accuracy : 0.5365

'Positive' Class : No

***MLeval: Machine Learning Model Evaluation***

Input: caret train function object

Averaging probs.

Group 1 type: repeatedcv

Observations: 771

Number of groups: 1

Observations per group: 771

Positive: Yes

Negative: No

Group: Group 1

Positive: 126

Negative: 645

***Performance Metrics***

Group 1 Optimal Informedness = 0.127537836840163

Group 1 AUC-ROC = 0.59

**Neural Net**

**PCNL_Adjuvant**_nn<-train(**Adjuvant** ~., data= **PCNL_Adjuvant**_train1, method="nnet", trControl= **PCNL_Adjuvant**_ctrl)

**PCNL_Adjuvant**_nn_predict<-predict(**PCNL_Adjuvant**_nn, **PCNL_Adjuvant**_test1)

**PCNL_Adjuvant**_nn_tb<-table(pred=factor(**PCNL_Adjuvant**_nn_predict), ref= **PCNL_Adjuvant**_test1$**Adjuvant**)

confusionMatrix(**PCNL_Adjuvant**_nn_tb)

res_**PCNL_Adjuvant**_nn<- evalm(**PCNL_Adjuvant**_nn)

**#Predicts all as ‘No’**

Confusion Matrix and Statistics

No Yes

No 272 59

Yes	0	0

Accuracy : 0.8218

95% CI : (0.7762, 0.8615)

No Information Rate : 0.8218

P-Value [Acc > NIR] : 0.5347

Kappa : 0

Mcnemar's Test P-Value : 4.321e-14

Sensitivity : 1.0000

Specificity : 0.0000

Pos Pred Value : 0.8218

Neg Pred Value : NaN

Prevalence : 0.8218

Detection Rate : 0.8218

Detection Prevalence : 1.0000

Balanced Accuracy : 0.5000

'Positive' Class : No

***MLeval: Machine Learning Model Evaluation***

Input: caret train function object

Averaging probs.

Group 1 type: repeatedcv

Observations: 771

Number of groups: 1

Observations per group: 771

Positive: Yes

Negative: No

Group: Group 1

Positive: 126

Negative: 645

***Performance Metrics***

Group 1 Optimal Informedness = 0

Group 1 AUC-ROC = 0.49

**Bayesian Generalised Linear Model**

**PCNL_Adjuvant**_bayes<-train(**Adjuvant** ~., data= **PCNL_Adjuvant**_train1, method="bayesglm", trControl= **PCNL_Adjuvant**_ctrl)

**PCNL_Adjuvant**_bayes_predict<-predict(**PCNL_Adjuvant**_bayes, **PCNL_Adjuvant**_test1)

**PCNL_Adjuvant**_bayes_tb<-table(pred=factor(**PCNL_Adjuvant**_bayes_predict), ref= **PCNL_Adjuvant**_test1$**Adjuvant**)

confusionMatrix(**PCNL_Adjuvant**_bayes_tb)

res_**PCNL_Adjuvant**_bayes<- evalm(**PCNL_Adjuvant**_bayes)

Confusion Matrix and Statistics

ref

pred	No Yes

No 266 51

Yes	6	8

Accuracy : 0.8278

95% CI : (0.7827, 0.8669)

No Information Rate : 0.8218

P-Value [Acc > NIR] : 0.4205

Kappa : 0.1619

Mcnemar's Test P-Value : 5.611e-09

Sensitivity : 0.9779

Specificity : 0.1356

Pos Pred Value : 0.8391

Neg Pred Value : 0.5714

Prevalence : 0.8218

Detection Rate : 0.8036

Detection Prevalence : 0.9577

Balanced Accuracy : 0.5568

'Positive' Class : No

***MLeval: Machine Learning Model Evaluation***

Input: caret train function object

Averaging probs.

Group 1 type: repeatedcv

Observations: 771

Number of groups: 1

Observations per group: 771

Positive: Yes

Negative: No

Group: Group 1

Positive: 126

Negative: 645

***Performance Metrics***

Group 1 Optimal Informedness = 0.270874861572536

Group 1 AUC-ROC = 0.67

**Generate ROC curves**

res_**PCNL_Adjuvant**<-evalm(list(**PCNL_Adjuvant**_rf, **PCNL_Adjuvant**_rpart, **PCNL_Adjuvant**_xgboost, **PCNL_Adjuvant**_logit, **PCNL_Adjuvant**_nn, **PCNL_Adjuvant**_bayes), gnames=c('Random Forest', 'Partitioning', 'Extreme Gradient Boosting', 'Logistic Regression', 'Neural Network', 'Bayesian Generalised Linear Model'))

**#Deep Neural Net**

PCNL_Adjuvant_train_predictors3<-as.matrix(PCNL_Adjuvant_train_predictors1)

PCNL_Adjuvant_train_outcome<-ifelse(PCNL_Adjuvant_train1$Adjuvant=="Yes", 1,0)

PCNL_Adjuvant_test_predictors3<-as.matrix(PCNL_Adjuvant_test_predictors1)

PCNL_Adjuvant_test_outcome<-ifelse(PCNL_Adjuvant_test1$Adjuvant=="Yes", 1,0)

library(keras)

library(tensorflow)

library(pROC)

Adjuvant_keras<-keras_model_sequential()

Adjuvant_keras %>%

layer_dense(units = 32, input_shape = c(43)) %>%

layer_activation('relu') %>%

layer_dense(units = 1) %>%

layer_activation('sigmoid')

Adjuvant_keras %>% compile(loss='binary_crossentropy', optimizer='rmsprop', metrics='accuracy')

Adjuvant_keras %>% fit(PCNL_Adjuvant_train_predictors3, PCNL_Adjuvant_train_outcome, epochs=200, batch_size=5, validation_split=0.2, verbose=2)

Adjuvant_keras_results<-Adjuvant_keras %>% evaluate(PCNL_Adjuvant_test_predictors3, PCNL_Adjuvant_test_outcome, verbose=1)

Adjuvant_keras_prediction<- Adjuvant_keras%>%predict(PCNL_Adjuvant_test_predictors3, batch_size=32, verbose=1)

Adjuvant_keras_prediction1<-round(Adjuvant_keras_prediction, digits=0)

Adjuvant_keras_tb<- table(PCNL_Adjuvant_test_outcome, Adjuvant_keras_prediction1)

Adjuvant_keras_roc<- roc(PCNL_Adjuvant_test_outcome, Adjuvant_keras_prediction1)

auc(Adjuvant_keras_roc)

confusionMatrix(Adjuvant_keras_tb)

plot(Adjuvant_keras_roc)

```
Area under the curve: 0.5302
Confusion Matrix and Statistics

                          Adjuvant_keras_prediction1
PCNL_Adjuvant_test_outcome   0   1
                         0 270   2
                         1  55   4

               Accuracy : 0.8278
                 95% CI : (0.7827, 0.8669)
    No Information Rate : 0.9819
    P-Value [Acc > NIR] : 1

                  Kappa : 0.0932

 Mcnemar's Test P-Value : 5.675e-12

            Sensitivity : 0.8308
            Specificity : 0.6667
         Pos Pred Value : 0.9926
         Neg Pred Value : 0.0678
             Prevalence : 0.9819
         Detection Rate : 0.8157
   Detection Prevalence : 0.8218
      Balanced Accuracy : 0.7487

       'Positive' Class : 0
```

13. **Post-operative Stay**

##Bins=> 0 days, 1 day, 2 days, ≥3 days##

#Create training/test sets:

PCNL_Stay_coded_1<-na.omit(PCNL_Stay_coded)

set.seed(1234)

PCNL_Stay_sample_coded<-sample(1:nrow(PCNL_Stay_coded_1), size=nrow(PCNL_Stay_coded_1) *0.7)

PCNL_Stay_train1<- PCNL_Stay_coded_1[PCNL_Stay_sample_coded,]

PCNL_Stay_test1<- PCNL_Stay_coded_1[-PCNL_Stay_sample_coded,]

PCNL_Stay_train_predictors1<- PCNL_Stay_train1[,-44]

PCNL_Stay_test_predictors1<- PCNL_Stay_test1[,-44]

PCNL_Stay_train1$Stay1<-as.factor(PCNL_Stay_train1$Stay1)

PCNL_Stay_test1$Stay1<-as.factor(PCNL_Stay_test1$Stay1)

PCNL_Stay_ctrl<-trainControl(method= "repeatedcv", number=10, classProbs=TRUE, savePredictions=TRUE)

#Sort data

PCNL_Post_Stay_1<-na.omit(PCNL_Stay_coded)

set.seed(1234)

PCNL_Post_Stay_sample_coded<-sample(1:nrow(PCNL_Post_Stay_1), size=nrow(PCNL_Post_Stay_1) *0.7)

PCNL_Post_Stay_train1<- PCNL_Post_Stay_1[PCNL_Post_Stay_sample_coded,]

PCNL_Post_Stay_test1<- PCNL_Post_Stay_1[-PCNL_Post_Stay_sample_coded,]

PCNL_Post_Stay_train_predictors1<- PCNL_Post_Stay_train1[,-44]

PCNL_Post_Stay_test_predictors1<- PCNL_Post_Stay_test1[,-44]

PCNL_Post_Stay_train_predictors3<-as.matrix(PCNL_Post_Stay_train_predictors1)

PCNL_Post_Stay_train_outcome1<-as.matrix(PCNL_Post_Stay_train1$Stay1)

PCNL_Post_Stay_train_outcome<-to_categorical(PCNL_Post_Stay_train_outcome1)

PCNL_Post_Stay_test_predictors3<-as.matrix(PCNL_Post_Stay_test_predictors1)

PCNL_Post_Stay_test_outcome1<-as.matrix(PCNL_Post_Stay_test1$Stay1)

PCNL_Post_Stay_test_outcome<-to_categorical(PCNL_Post_Stay_test_outcome1)

#Random Forests

PCNL_Stay_rf<- train(Stay1 ~., data= PCNL_Stay_train1, method="cforest", trControl= PCNL_Stay_ctrl)

PCNL_Stay_rf_predict<-predict(PCNL_Stay_rf, newdata=PCNL_Stay_test_predictors1)

PCNL_Stay_rf_tb<-table(PCNL_Stay_rf_predict, ref=PCNL_Stay_test1$Stay1)

confusionMatrix(PCNL_Stay_rf_tb)

res_PCNL_Stay_rf<-evalm(PCNL_Stay_rf)

AUC=0.79

```
Confusion Matrix and Statistics

                    ref
PCNL_Stay_rf_predict One Three Two Zero
               One   189    37  53    8
               Three  59   664 227    3
               Two    22    42  22    0
               Zero    0     0   0    0

Overall Statistics

               Accuracy : 0.6599
                 95% CI : (0.6337, 0.6854)
    No Information Rate : 0.5603
    P-Value [Acc > NIR] : 9.196e-14

                  Kappa : 0.3683

 Mcnemar's Test P-Value : NA

Statistics by Class:

                     Class: One Class: Three Class: Two Class: Zero
Sensitivity              0.7000       0.8937    0.07285    0.000000
Specificity              0.9072       0.5043    0.93750    1.000000
Pos Pred Value           0.6585       0.6967    0.25581         NaN
Neg Pred Value           0.9220       0.7882    0.77419    0.991704
Prevalence               0.2036       0.5603    0.22775    0.008296
Detection Rate           0.1425       0.5008    0.01659    0.000000
Detection Prevalence     0.2164       0.7187    0.06486    0.000000
Balanced Accuracy        0.8036       0.6990    0.50517    0.500000
***MLeval: Machine Learning Model Evaluation***
Input: caret train function object
Averaging probs.
Group 1 type: repeatedcv
Observations: 3092
Number of groups: 1
Observations per group: 3092
Positive: Three
Negative: One
Group: Group 1
Positive: 1739
Negative: 616
***Performance Metrics***
```

#Partitioning

PCNL_Stay_rpart<-train(Stay1 ~ ., data= PCNL_Stay_train1, method="rpart", trControl= PCNL_Stay_ctrl)

PCNL_Stay_rpart_predict<-predict(PCNL_Stay_rpart, newdata= PCNL_Stay_test1)

PCNL_Stay_rpart_tb<-table(pred=factor(PCNL_Stay_rpart_predict), ref= PCNL_Stay_test1$Stay1)

confusionMatrix(PCNL_Stay_rpart_tb)

res_PCNL_Stay_rpart<- evalm(PCNL_Stay_rpart)

AUC=0.68

```
Confusion Matrix and Statistics

      One Three Two Zero
One   198    68  56    8
Three  72   675 246    3
Two     0     0   0    0
Zero    0     0   0    0

Overall Statistics

               Accuracy : 0.6584
                 95% CI : (0.6321, 0.6839)
    No Information Rate : 0.5603
    P-Value [Acc > NIR] : 2.141e-13

                  Kappa : 0.3535

 Mcnemar's Test P-Value : NA

Statistics by Class:

                     Class: One Class: Three Class: Two Class: Zero
Sensitivity              0.7333       0.9085     0.0000    0.000000
Specificity              0.8750       0.4494     1.0000    1.000000
Pos Pred Value           0.6000       0.6777        NaN         NaN
Neg Pred Value           0.9277       0.7939     0.7722    0.991704
Prevalence               0.2036       0.5603     0.2278    0.008296
Detection Rate           0.1493       0.5090     0.0000    0.000000
Detection Prevalence     0.2489       0.7511     0.0000    0.000000
Balanced Accuracy        0.8042       0.6789     0.5000    0.500000
***MLeval: Machine Learning Model Evaluation***
Input: caret train function object
Averaging probs.
Group 1 type: repeatedcv
Observations: 3092
Number of groups: 1
Observations per group: 3092
Positive: Three
Negative: One
Group: Group 1
Positive: 1739
Negative: 616
***Performance Metrics***
```

#XGBoost

PCNL_Stay_xgboost<-train(Stay1 ~., data= PCNL_Stay_train1, method="xgbTree", trControl= PCNL_Stay_ctrl)

PCNL_Stay_xgboost_predict<-predict(PCNL_Stay_xgboost, PCNL_Stay_test1)

PCNL_Stay_xgboost_tb<-table(pred=factor(PCNL_Stay_xgboost_predict), ref= PCNL_Stay_test1$Stay1)

confusionMatrix(PCNL_Stay_xgboost_tb)

res_PCNL_Stay_xgboost<- evalm(PCNL_Stay_xgboost)

AUC=0.78

```
Confusion Matrix and Statistics

      One Three Two Zero
One   203    41  57    8
Three  65   697 245    3
Two     2     5   0    0
Zero    0     0   0    0

Overall Statistics

               Accuracy : 0.6787
                 95% CI : (0.6528, 0.7038)
    No Information Rate : 0.5603
    P-Value [Acc > NIR] : < 2.2e-16

                  Kappa : 0.3875

 Mcnemar's Test P-Value : NA

Statistics by Class:

                     Class: One Class: Three Class: Two Class: Zero
Sensitivity              0.7519       0.9381   0.000000    0.000000
Specificity              0.8996       0.4631   0.993164    1.000000
Pos Pred Value           0.6570       0.6901   0.000000         NaN
Neg Pred Value           0.9341       0.8544   0.771039    0.991704
Prevalence               0.2036       0.5603   0.227753    0.008296
Detection Rate           0.1531       0.5256   0.000000    0.000000
Detection Prevalence     0.2330       0.7617   0.005279    0.000000
Balanced Accuracy        0.8257       0.7006   0.496582    0.500000
***MLeval: Machine Learning Model Evaluation***
Input: caret train function object
Averaging probs.
Group 1 type: repeatedcv
Observations: 3092
Number of groups: 1
Observations per group: 3092
Positive: Three
Negative: One
Group: Group 1
Positive: 1739
Negative: 616
***Performance Metrics***
```

#Logistic Regression

PCNL_Stay_logit<-train(Stay1 ~., data= PCNL_Stay_train1, method="LogitBoost", trControl= PCNL_Stay_ctrl)

PCNL_Stay_logit_predict<-predict(PCNL_Stay_logit, PCNL_Stay_test1)

PCNL_Stay_logit_tb<-table(pred=factor(PCNL_Stay_logit_predict), ref= PCNL_Stay_test1$Stay1)

confusionMatrix(PCNL_Stay_logit_tb)

res_PCNL_Stay_logit<- evalm(PCNL_Stay_logit)

AUC=0.75

```
Confusion Matrix and Statistics

      One Three Two Zero
One   190    62  53    8
Three  57   648 231    3
Two     0     8   2    0
Zero    0     0   0    0

Overall Statistics

               Accuracy : 0.6656
                 95% CI : (0.6388, 0.6916)
    No Information Rate : 0.5689
    P-Value [Acc > NIR] : 1.359e-12

                  Kappa : 0.3647

 Mcnemar's Test P-Value : NA

Statistics by Class:

                     Class: One Class: Three Class: Two Class: Zero
Sensitivity              0.7692       0.9025   0.006993    0.000000
Specificity              0.8788       0.4651   0.991803    1.000000
Pos Pred Value           0.6070       0.6901   0.200000         NaN
Neg Pred Value           0.9399       0.7833   0.773163    0.991284
Prevalence               0.1957       0.5689   0.226624    0.008716
Detection Rate           0.1506       0.5135   0.001585    0.000000
Detection Prevalence     0.2480       0.7441   0.007924    0.000000
Balanced Accuracy        0.8240       0.6838   0.499398    0.500000
***MLeval: Machine Learning Model Evaluation***
Input: caret train function object
Averaging probs.
Group 1 type: repeatedcv
Observations: 3092
Number of groups: 1
Observations per group: 3092
Positive: Three
Negative: One
Group: Group 1
Positive: 1739
Negative: 616
***Performance Metrics***
```

#Neural Net

PCNL_Stay_nn<-train(Stay1 ~., data= PCNL_Stay_train1, method="nnet", trControl= PCNL_Stay_ctrl)

PCNL_Stay_nn_predict<-predict(PCNL_Stay_nn, PCNL_Stay_test1)

PCNL_Stay_nn_tb<-table(pred=factor(PCNL_Stay_nn_predict), ref= PCNL_Stay_test1$Stay1)

confusionMatrix(PCNL_Stay_nn_tb)

res_PCNL_Stay_nn<- evalm(PCNL_Stay_nn)

AUC=0.75

```
Confusion Matrix and Statistics

      One Three Two Zero
One   194    41  56    8
Three  76   702 246    3
Two     0     0   0    0
Zero    0     0   0    0

Overall Statistics

               Accuracy : 0.6757
                 95% CI : (0.6498, 0.7009)
    No Information Rate : 0.5603
    P-Value [Acc > NIR] : < 2.2e-16

                  Kappa : 0.3765

 Mcnemar's Test P-Value : NA

Statistics by Class:

                     Class: One Class: Three Class: Two Class: Zero
Sensitivity              0.7185       0.9448     0.0000    0.000000
Specificity              0.9006       0.4425     1.0000    1.000000
Pos Pred Value           0.6488       0.6835        NaN         NaN
Neg Pred Value           0.9260       0.8629     0.7722    0.991704
Prevalence               0.2036       0.5603     0.2278    0.008296
Detection Rate           0.1463       0.5294     0.0000    0.000000
Detection Prevalence     0.2255       0.7745     0.0000    0.000000
Balanced Accuracy        0.8095       0.6937     0.5000    0.500000
***MLeval: Machine Learning Model Evaluation***
Input: caret train function object
Averaging probs.
Group 1 type: repeatedcv
Observations: 3092
Number of groups: 1
Observations per group: 3092
Positive: Three
Negative: One
Group: Group 1
Positive: 1739
Negative: 616
***Performance Metrics***
```

#Bayesian Generalised Linear Model

PCNL_Stay_bayes<-train(Stay1 ~., data= PCNL_Stay_train1, method="bayesglm", trControl= PCNL_Stay_ctrl)

PCNL_Stay_bayes_predict<-predict(PCNL_Stay_bayes, PCNL_Stay_test1)

PCNL_Stay_bayes_tb<-table(pred=factor(PCNL_Stay_bayes_predict), ref= PCNL_Stay_test1$Stay1)

confusionMatrix(PCNL_Stay_bayes_tb)

res_PCNL_Stay_bayes<- evalm(PCNL_Stay_bayes)

AUC=0.73

```
Confusion Matrix and Statistics

      One Three Two Zero
One     2     1   0    0
Three 268   741 302   11
Two     0     1   0    0
Zero    0     0   0    0

Overall Statistics

               Accuracy : 0.5603
                 95% CI : (0.5331, 0.5873)
    No Information Rate : 0.5603
    P-Value [Acc > NIR] : 0.5115

                  Kappa : 0.0024

 Mcnemar's Test P-Value : NA

Statistics by Class:

                     Class: One Class: Three Class: Two Class: Zero
Sensitivity            0.007407     0.997308  0.0000000    0.000000
Specificity            0.999053     0.003431  0.9990234    1.000000
Pos Pred Value         0.666667     0.560514  0.0000000         NaN
Neg Pred Value         0.797430     0.500000  0.7720755    0.991704
Prevalence             0.203620     0.560332  0.2277526    0.008296
Detection Rate         0.001508     0.558824  0.0000000    0.000000
Detection Prevalence   0.002262     0.996983  0.0007541    0.000000
Balanced Accuracy      0.503230     0.500369  0.4995117    0.500000
***MLeval: Machine Learning Model Evaluation***
Input: caret train function object
Averaging probs.
Group 1 type: repeatedcv
Observations: 3092
Number of groups: 1
Observations per group: 3092
Positive: Three
Negative: One
Group: Group 1
Positive: 1739
Negative: 616
***Performance Metrics***
```

#Generate ROC curves

res_PCNL_Stay<-evalm(list(PCNL_Stay_rf, PCNL_Stay_rpart, PCNL_Stay_xgboost, PCNL_Stay_logit, PCNL_Stay_nn, PCNL_Stay_bayes), gnames=c('Random Forest', 'Partitioning', 'Extreme Gradient Boosting', 'Logistic Regression', 'Neural Network', 'Bayesian Generalised Linear Model'))

**#Deep NN**

Post_Stay_keras<-keras_model_sequential()

Post_Stay_keras %>%

layer_dense(units = 32, input_shape = c(43)) %>%

layer_activation('relu') %>%

layer_dense(units = 4) %>%

layer_activation('softmax')

Post_Stay_keras %>% compile(loss='categorical_crossentropy', optimizer='rmsprop', metrics='accuracy')

Post_Stay_keras %>% fit(PCNL_Post_Stay_train_predictors3, PCNL_Post_Stay_train_outcome, epochs=200, batch_size=5, validation_split=0.2, verbose=2)

Post_Stay_keras_results<-Post_Stay_keras %>% evaluate(PCNL_Post_Stay_test_predictors3, PCNL_Post_Stay_test_outcome, verbose=1)

Post_Stay_keras_prediction<- Post_Stay_keras%>%predict(PCNL_Post_Stay_test_predictors3, batch_size=32, verbose=1)

Post_Stay_keras_prediction1<-round(Post_Stay_keras_prediction, digits=0)

Post_Stay_keras_tb<- table(PCNL_Post_Stay_test_outcome, Post_Stay_keras_prediction1)

Post_Stay_keras_roc<- roc(PCNL_Post_Stay_test_outcome, Post_Stay_keras_prediction1)

auc(Post_Stay_keras_roc)

confusionMatrix(Post_Stay_keras_tb)

plot(Post_Stay_keras_roc)

```
Area under the curve: 0.6225
Confusion Matrix and Statistics

                           Post_Stay_keras_prediction1
PCNL_Post_Stay_test_outcome    0    1
                          0 3255  723
                          1  760  566

               Accuracy : 0.7204
                 95% CI : (0.7081, 0.7324)
    No Information Rate : 0.757
    P-Value [Acc > NIR] : 1.0000

                  Kappa : 0.2474

 Mcnemar's Test P-Value : 0.3499

            Sensitivity : 0.8107
            Specificity : 0.4391
         Pos Pred Value : 0.8183
         Neg Pred Value : 0.4268
             Prevalence : 0.7570
         Detection Rate : 0.6137
   Detection Prevalence : 0.7500
      Balanced Accuracy : 0.6249

       'Positive' Class : 0
```

Post_Stay_keras_tb1<-table(PCNL_Post_Stay_test_outcome$V1, Post_Stay_keras_prediction1$V1)

Post_Stay_keras_roc1<-roc(PCNL_Post_Stay_test_outcome$V1, Post_Stay_keras_prediction1$V1)

auc(Post_Stay_keras_roc1)

confusionMatrix(Post_Stay_keras_tb1)

Area under the curve: 0.5

Confusion Matrix and Statistics

0	1

0 1309	0

1	17	0

Accuracy : 0.9872

95% CI : (0.9796, 0.9925)

No Information Rate : 1

P-Value [Acc > NIR] : 1.0000000

Kappa : 0

Mcnemar's Test P-Value : 0.0001042

Sensitivity : 0.9872

Specificity : NA

Pos Pred Value : NA

Neg Pred Value : NA

Prevalence : 1.0000

Detection Rate : 0.9872

Detection Prevalence : 0.9872

Balanced Accuracy : NA

'Positive' Class : 0

Post_Stay_keras_tb2<-table(PCNL_Post_Stay_test_outcome$V2, Post_Stay_keras_prediction1$V2)

Post_Stay_keras_roc2<-roc(PCNL_Post_Stay_test_outcome$V2, Post_Stay_keras_prediction1$V2)

auc(Post_Stay_keras_roc2)

confusionMatrix(Post_Stay_keras_tb2)

Area under the curve: 0.5613

Confusion Matrix and Statistics

0	1

0 1031	30

1 225	40

Accuracy : 0.8077

95% CI : (0.7854, 0.8286)

No Information Rate : 0.9472

P-Value [Acc > NIR] : 1

Kappa : 0.1694

Mcnemar's Test P-Value : <2e-16

Sensitivity : 0.8209

Specificity : 0.5714

Pos Pred Value : 0.9717

Neg Pred Value : 0.1509

Prevalence : 0.9472

Detection Rate : 0.7775

Detection Prevalence : 0.8002

Balanced Accuracy : 0.6961

'Positive' Class : 0

Post_Stay_keras_tb3<-table(PCNL_Post_Stay_test_outcome$V3, Post_Stay_keras_prediction1$V3)

Post_Stay_keras_roc3<-roc(PCNL_Post_Stay_test_outcome$V3, Post_Stay_keras_prediction1$V3)

auc(Post_Stay_keras_roc3)

confusionMatrix(Post_Stay_keras_tb3)

Area under the curve: 0.5

Confusion Matrix and Statistics

0	1

0 1009	0

1 317	0

Accuracy : 0.7609

95% CI : (0.737, 0.7837)

No Information Rate : 1

P-Value [Acc > NIR] : 1

Kappa : 0

Mcnemar's Test P-Value : <2e-16

Sensitivity : 0.7609

Specificity : NA

Pos Pred Value : NA

Neg Pred Value : NA

Prevalence : 1.0000

Detection Rate : 0.7609

Detection Prevalence : 0.7609

Balanced Accuracy : NA

'Positive' Class : 0

Post_Stay_keras_tb4<-table(PCNL_Post_Stay_test_outcome$V4, Post_Stay_keras_prediction1$V4)

Post_Stay_keras_roc4<-roc(PCNL_Post_Stay_test_outcome$V4, Post_Stay_keras_prediction1$V4)

auc(Post_Stay_keras_roc4)

confusionMatrix(Post_Stay_keras_tb4)

Area under the curve: 0.5388

Confusion Matrix and Statistics

0	1

0 58 541

1 14 713

Accuracy : 0.5814

95% CI : (0.5544, 0.6082)

No Information Rate : 0.9457

P-Value [Acc > NIR] : 1

Kappa : 0.0841

Mcnemar's Test P-Value : <2e-16

Sensitivity : 0.80556

Specificity : 0.56858

Pos Pred Value : 0.09683

Neg Pred Value : 0.98074

Prevalence : 0.05430

Detection Rate : 0.04374

Detection Prevalence : 0.45173

Balanced Accuracy : 0.68707

'Positive' Class : 0

14. **Clavien Dindo Classification**

##Bins = 1=Nil, 2=I, 3=IIa, 4=IIb, 5=IIIa, 6=IIIb, 7=IVa, 8=V (no IVb complications in dataset)

#Deep NN

PCNL_Clavien_coded3<-na.omit(PCNL_Clavien_coded2)

set.seed(1234)

PCNL_Clavien_sample_coded<-sample(1:nrow(PCNL_Clavien_coded3), size=nrow(PCNL_Clavien_coded3) *0.7)

PCNL_Clavien_train1<- PCNL_Clavien_coded3[PCNL_Clavien_sample_coded,]

PCNL_Clavien_test1<- PCNL_Clavien_coded3[-PCNL_Clavien_sample_coded,]

PCNL_Clavien_train_predictors1<- PCNL_Clavien_train1[,-44]

PCNL_Clavien_test_predictors1<- PCNL_Clavien_test1[,-44]

PCNL_Clavien_train_predictors3<-as.matrix(PCNL_Clavien_train_predictors1)

PCNL_Clavien_train_outcome1<-as.matrix(PCNL_Clavien_train1$Clavien2)

PCNL_Clavien_train_outcome<-to_categorical(PCNL_Clavien_train_outcome1)

PCNL_Clavien_test_predictors3<-as.matrix(PCNL_Clavien_test_predictors1)

PCNL_Clavien_test_outcome1<-as.matrix(PCNL_Clavien_test1$Clavien2)

PCNL_Clavien_test_outcome<-to_categorical(PCNL_Clavien_test_outcome1)

#Create training/test sets:

PCNL_Clavien_coded_1<-na.omit(PCNL_Clavien_coded)

set.seed(1234)

PCNL_Clavien_sample_coded<-sample(1:nrow(PCNL_Clavien_coded_1), size=nrow(PCNL_Clavien_coded_1) *0.7)

PCNL_Clavien_train1<- PCNL_Clavien_coded_1[PCNL_Clavien_sample_coded,]

PCNL_Clavien_test1<- PCNL_Clavien_coded_1[-PCNL_Clavien_sample_coded,]

PCNL_Clavien_train_predictors1<- PCNL_Clavien_train1[,-44]

PCNL_Clavien_test_predictors1<- PCNL_Clavien_test1[,-44]

PCNL_Clavien_ctrl<-trainControl(method= "repeatedcv", number=10, classProbs=TRUE, savePredictions=TRUE)

**#Random Forests**

PCNL_Clavien_rf<- train(Clavien ~., data= PCNL_Clavien_train1, method="cforest", trControl= PCNL_Clavien_ctrl)

PCNL_Clavien_rf_predict<-predict(PCNL_Clavien_rf, newdata=PCNL_Clavien_test_predictors1)

PCNL_Clavien_rf_tb<-table(PCNL_Clavien_rf_predict, ref=PCNL_Clavien_test1$Clavien)

confusionMatrix(PCNL_Clavien_rf_tb)

res_File_Name_rf<-evalm(PCNL_Clavien_rf)

AUC=0.90

```
Confusion Matrix and Statistics

                       ref
PCNL_Clavien_rf_predict    I   II IIIa IIIb  IVa    V Zero
                   I      26    7    1    1    1    0   10
                   II      8   32    7    2    2    1   19
                   IIIa    0    0    5    0    0    0    0
                   IIIb    0    0    0    2    0    0    0
                   IVa     0    0    0    0    0    0    0
                   V       0    0    0    0    0    0    0
                   Zero   53   62   14   12    2    1 1058

Overall Statistics

               Accuracy : 0.8469
                 95% CI : (0.8264, 0.8659)
    No Information Rate : 0.8198
    P-Value [Acc > NIR] : 0.004926

                  Kappa : 0.3887

 Mcnemar's Test P-Value : NA

Statistics by Class:

                     Class: I Class: II Class: IIIa Class: IIIb Class: IVa Class: V Class: Zero
Sensitivity           0.29885   0.31683    0.185185    0.117647   0.000000 0.000000      0.9733
Specificity           0.98386   0.96816    1.000000    1.000000   1.000000 1.000000      0.3975
Pos Pred Value        0.56522   0.45070    1.000000    1.000000        NaN      NaN      0.8802
Neg Pred Value        0.95234   0.94502    0.983346    0.988671   0.996229 0.998492      0.7661
Prevalence            0.06561   0.07617    0.020362    0.012821   0.003771 0.001508      0.8198
Detection Rate        0.01961   0.02413    0.003771    0.001508   0.000000 0.000000      0.7979
Detection Prevalence  0.03469   0.05354    0.003771    0.001508   0.000000 0.000000      0.9065
Balanced Accuracy     0.64135   0.64250    0.592593    0.558824   0.500000 0.500000      0.6854
***MLeval: Machine Learning Model Evaluation***
Input: caret train function object
Averaging probs.
Group 1 type: repeatedcv
Observations: 3092
Number of groups: 1
Observations per group: 3092
Positive: II
Negative: I
Group: Group 1
Positive: 242
Negative: 224
***Performance Metrics***
```

**#Partitioning**

PCNL_Clavien_rpart<-train(Clavien ~ ., data= PCNL_Clavien_train1, method="rpart", trControl= PCNL_Clavien_ctrl)

PCNL_Clavien_rpart_predict<-predict(PCNL_Clavien_rpart, newdata= PCNL_Clavien_test1)

PCNL_Clavien_rpart_tb<-table(pred=factor(PCNL_Clavien_rpart_predict), ref= PCNL_Clavien_test1$Clavien)

confusionMatrix(PCNL_Clavien_rpart_tb)

res_PCNL_Clavien_rpart<- evalm(PCNL_Clavien_rpart)

AUC=0.66

```
Confusion Matrix and Statistics

        I   II IIIa IIIb  IVa    V Zero
I       6    1    1    0    0    0    0
II      7   16    3    0    1    1    4
IIIa    0    0    0    0    0    0    0
IIIb    0    0    0    0    0    0    0
IVa     0    0    0    0    0    0    0
V       0    0    0    0    0    0    0
Zero   74   84   23   17    4    1 1083

Overall Statistics

               Accuracy : 0.8333
                 95% CI : (0.8122, 0.853)
    No Information Rate : 0.8198
    P-Value [Acc > NIR] : 0.1048

                  Kappa : 0.1779

 Mcnemar's Test P-Value : NA

Statistics by Class:

                     Class: I Class: II Class: IIIa Class: IIIb Class: IVa Class: V Class: Zero
Sensitivity          0.068966   0.15842     0.00000     0.00000   0.000000 0.000000      0.9963
Specificity          0.998386   0.98694     1.00000     1.00000   1.000000 1.000000      0.1506
Pos Pred Value       0.750000   0.50000         NaN         NaN        NaN      NaN      0.8421
Neg Pred Value       0.938543   0.93431     0.97964     0.98718   0.996229 0.998492      0.9000
Prevalence           0.065611   0.07617     0.02036     0.01282   0.003771 0.001508      0.8198
Detection Rate       0.004525   0.01207     0.00000     0.00000   0.000000 0.000000      0.8167
Detection Prevalence 0.006033   0.02413     0.00000     0.00000   0.000000 0.000000      0.9698
Balanced Accuracy    0.533676   0.57268     0.50000     0.50000   0.500000 0.500000      0.5735
***MLeval: Machine Learning Model Evaluation***
Input: caret train function object
Averaging probs.
Group 1 type: repeatedcv
Observations: 3092
Number of groups: 1
Observations per group: 3092
Positive: II
Negative: I
Group: Group 1
Positive: 242
Negative: 224
***Performance Metrics***
```

**#XGBoost**

PCNL_Clavien_xgboost<-train(Clavien ~., data= PCNL_Clavien_train1, method="xgbTree", trControl= PCNL_Clavien_ctrl)

PCNL_Clavien_xgboost_predict<-predict(PCNL_Clavien_xgboost, PCNL_Clavien_test1)

PCNL_Clavien_xgboost_tb<-table(pred=factor(PCNL_Clavien_xgboost_predict), ref= PCNL_Clavien_test1$Clavien)

confusionMatrix(PCNL_Clavien_xgboost_tb)

res_PCNL_Clavien_xgboost<- evalm(PCNL_Clavien_xgboost)

AUC=0.88

```
Confusion Matrix and Statistics

        I   II IIIa IIIb  IVa    V Zero
I      17    4    0    1    0    0    2
II      9   31    6    1    2    1   11
IIIa    0    0    2    1    0    0    0
IIIb    0    1    0    0    0    0    0
IVa     0    0    0    0    0    0    0
V       0    0    0    0    0    0    0
Zero   61   65   19   14    3    1 1074

Overall Statistics

               Accuracy : 0.8477
                 95% CI : (0.8272, 0.8666)
    No Information Rate : 0.8198
    P-Value [Acc > NIR] : 0.003959

                  Kappa : 0.3391

 Mcnemar's Test P-Value : NA

Statistics by Class:

                     Class: I Class: II Class: IIIa Class: IIIb Class: IVa Class: V Class: Zero
Sensitivity           0.19540   0.30693    0.074074   0.0000000   0.000000 0.000000      0.9880
Specificity           0.99435   0.97551    0.999230   0.9992361   1.000000 1.000000      0.3180
Pos Pred Value        0.70833   0.50820    0.666667   0.0000000        NaN      NaN      0.8682
Neg Pred Value        0.94624   0.94466    0.981104   0.9871698   0.996229 0.998492      0.8539
Prevalence            0.06561   0.07617    0.020362   0.0128205   0.003771 0.001508      0.8198
Detection Rate        0.01282   0.02338    0.001508   0.0000000   0.000000 0.000000      0.8100
Detection Prevalence  0.01810   0.04600    0.002262   0.0007541   0.000000 0.000000      0.9329
Balanced Accuracy     0.59488   0.64122    0.536652   0.4996180   0.500000 0.500000      0.6530
***MLeval: Machine Learning Model Evaluation***
Input: caret train function object
Averaging probs.
Group 1 type: repeatedcv
Observations: 3092
Number of groups: 1
Observations per group: 3092
Positive: II
Negative: I
Group: Group 1
Positive: 242
Negative: 224
***Performance Metrics***
```

**#Logistic Regression**

PCNL_Clavien_logit<-train(Clavien ~., data= PCNL_Clavien_train1, method="LogitBoost", trControl= PCNL_Clavien_ctrl)

PCNL_Clavien_logit_predict<-predict(PCNL_Clavien_logit, PCNL_Clavien_test1)

PCNL_Clavien_logit_tb<-table(pred=factor(PCNL_Clavien_logit_predict), ref= PCNL_Clavien_test1$Clavien)

confusionMatrix(PCNL_Clavien_logit_tb)

res_File_Name_logit<- evalm(PCNL_Clavien_logit)

AUC=0.82

```
Confusion Matrix and Statistics

        I   II IIIa IIIb  IVa    V Zero
I       0    0    0    0    0    0    3
II      6   17    4    0    3    0   10
IIIa    0    0    0    0    0    0    0
IIIb    0    0    0    0    0    0    0
IVa     0    0    0    0    0    0    0
V       0    0    0    0    0    0    0
Zero   53   61   13   13    2    1 1022

Overall Statistics

               Accuracy : 0.8601
                 95% CI : (0.8392, 0.8792)
    No Information Rate : 0.8568
    P-Value [Acc > NIR] : 0.3903

                  Kappa : 0.184

 Mcnemar's Test P-Value : NA

Statistics by Class:

                     Class: I Class: II Class: IIIa Class: IIIb Class: IVa  Class: V Class: Zero
Sensitivity          0.000000   0.21795     0.00000     0.00000   0.000000 0.0000000      0.9874
Specificity          0.997389   0.97965     1.00000     1.00000   1.000000 1.0000000      0.1734
Pos Pred Value       0.000000   0.42500         NaN         NaN        NaN       NaN      0.8773
Neg Pred Value       0.951037   0.94777     0.98593     0.98924   0.995861 0.9991722      0.6977
Prevalence           0.048841   0.06457     0.01407     0.01076   0.004139 0.0008278      0.8568
Detection Rate       0.000000   0.01407     0.00000     0.00000   0.000000 0.0000000      0.8460
Detection Prevalence 0.002483   0.03311     0.00000     0.00000   0.000000 0.0000000      0.9644
Balanced Accuracy    0.498695   0.59880     0.50000     0.50000   0.500000 0.5000000      0.5804
***MLeval: Machine Learning Model Evaluation***
Input: caret train function object
Averaging probs.
Group 1 type: repeatedcv
Observations: 3092
Number of groups: 1
Observations per group: 3092
Positive: II
Negative: I
Group: Group 1
Positive: 242
Negative: 224
***Performance Metrics***
```

**#Neural Net**

PCNL_Clavien_nn<-train(Clavien ~., data= PCNL_Clavien_train1, method="nnet", trControl= PCNL_Clavien_ctrl)

PCNL_Clavien_nn_predict<-predict(PCNL_Clavien_nn, PCNL_Clavien_test1)

PCNL_Clavien_nn_tb<-table(pred=factor(PCNL_Clavien_nn_predict), ref= PCNL_Clavien_test1$Clavien)

confusionMatrix(PCNL_Clavien_nn_tb)

res_PCNL_Clavien_nn<- evalm(PCNL_Clavien_nn)

AUC=0.5

```
Confusion Matrix and Statistics

        I   II IIIa IIIb  IVa    V Zero
I       0    0    0    0    0    0    0
II      0    0    0    0    0    0    0
IIIa    0    0    0    0    0    0    0
IIIb    0    0    0    0    0    0    0
IVa     0    0    0    0    0    0    0
V       0    0    0    0    0    0    0
Zero   87  101   27   17    5    2 1087

Overall Statistics

               Accuracy : 0.8198
                 95% CI : (0.798, 0.8401)
    No Information Rate : 0.8198
    P-Value [Acc > NIR] : 0.5173

                  Kappa : 0

 Mcnemar's Test P-Value : NA

Statistics by Class:

                     Class: I Class: II Class: IIIa Class: IIIb Class: IVa Class: V Class: Zero
Sensitivity           0.00000   0.00000     0.00000     0.00000   0.000000 0.000000      1.0000
Specificity           1.00000   1.00000     1.00000     1.00000   1.000000 1.000000      0.0000
Pos Pred Value            NaN       NaN         NaN         NaN        NaN      NaN      0.8198
Neg Pred Value        0.93439   0.92383     0.97964     0.98718   0.996229 0.998492         NaN
Prevalence            0.06561   0.07617     0.02036     0.01282   0.003771 0.001508      0.8198
Detection Rate        0.00000   0.00000     0.00000     0.00000   0.000000 0.000000      0.8198
Detection Prevalence  0.00000   0.00000     0.00000     0.00000   0.000000 0.000000      1.0000
Balanced Accuracy     0.50000   0.50000     0.50000     0.50000   0.500000 0.500000      0.5000
***MLeval: Machine Learning Model Evaluation***
Input: caret train function object
Averaging probs.
Group 1 type: repeatedcv
Observations: 3092
Number of groups: 1
Observations per group: 3092
Positive: II
Negative: I
Group: Group 1
Positive: 242
Negative: 224
***Performance Metrics***
```

**#Bayesian Generalised Linear Model**

PCNL_Clavien_bayes<-train(Clavien ~., data= PCNL_Clavien_train1, method="bayesglm", trControl= PCNL_Clavien_ctrl)

PCNL_Clavien_bayes_predict<-predict(PCNL_Clavien_bayes, PCNL_Clavien_test1)

PCNL_Clavien_bayes_tb<-table(pred=factor(PCNL_Clavien_bayes_predict), ref= PCNL_Clavien_test1$Clavien)

confusionMatrix(PCNL_Clavien_bayes_tb)

res_PCNL_Clavien_bayes<- evalm(PCNL_Clavien_bayes)

AUC=0.79

```
Confusion Matrix and Statistics

        I   II IIIa IIIb  IVa    V Zero
I       0    0    0    0    0    0    0
II      0    0    0    0    0    0    0
IIIa    0    0    0    0    0    0    0
IIIb    0    0    0    0    0    0    0
IVa     0    0    0    0    0    0    0
V       0    0    0    0    0    0    0
Zero   87  101   27   17    5    2 1087

Overall Statistics

               Accuracy : 0.8198
                 95% CI : (0.798, 0.8401)
    No Information Rate : 0.8198
    P-Value [Acc > NIR] : 0.5173

                  Kappa : 0

 Mcnemar's Test P-Value : NA

Statistics by Class:

                     Class: I Class: II Class: IIIa Class: IIIb Class: IVa Class: V Class: Zero
Sensitivity           0.00000   0.00000     0.00000     0.00000   0.000000 0.000000      1.0000
Specificity           1.00000   1.00000     1.00000     1.00000   1.000000 1.000000      0.0000
Pos Pred Value            NaN       NaN         NaN         NaN        NaN      NaN      0.8198
Neg Pred Value        0.93439   0.92383     0.97964     0.98718   0.996229 0.998492         NaN
Prevalence            0.06561   0.07617     0.02036     0.01282   0.003771 0.001508      0.8198
Detection Rate        0.00000   0.00000     0.00000     0.00000   0.000000 0.000000      0.8198
Detection Prevalence  0.00000   0.00000     0.00000     0.00000   0.000000 0.000000      1.0000
Balanced Accuracy     0.50000   0.50000     0.50000     0.50000   0.500000 0.500000      0.5000
***MLeval: Machine Learning Model Evaluation***
Input: caret train function object
Averaging probs.
Group 1 type: repeatedcv
Observations: 3092
Number of groups: 1
Observations per group: 3092
Positive: II
Negative: I
Group: Group 1
Positive: 242
Negative: 224
***Performance Metrics***
```

**#Generate ROC curves**

res_PCNL_Clavien<-evalm(list(PCNL_Clavien_rf, PCNL_Clavien_rpart, PCNL_Clavien_xgboost, PCNL_Clavien_logit, PCNL_Clavien_nn, PCNL_Clavien_bayes), gnames=c('Random Forest', 'Partitioning', 'Extreme Gradient Boosting', 'Logistic Regression', 'Neural Network', 'Bayesian Generalised Linear Model'))

**#Deep NN**

Clavien_keras<-keras_model_sequential()

Clavien_keras %>%

layer_dense(units = 32, input_shape = c(43)) %>%

layer_activation('relu') %>%

layer_dense(units = 8) %>%

layer_activation('softmax')

Clavien_keras %>% compile(loss='categorical_crossentropy', optimizer='rmsprop', metrics='accuracy')

Clavien_keras %>% fit(PCNL_Clavien_train_predictors3, PCNL_Clavien_train_outcome, epochs=200, batch_size=5, validation_split=0.2, verbose=2)

Clavien_keras_results<-Clavien_keras %>% evaluate(PCNL_Clavien_test_predictors3, PCNL_Clavien_test_outcome, verbose=1)

Clavien_keras_prediction<- Clavien_keras%>%predict(PCNL_Clavien_test_predictors3, batch_size=32, verbose=1)

Clavien_keras_prediction1<-round(Clavien_keras_prediction, digits=0)

Clavien_keras_tb<- table(PCNL_Clavien_test_outcome, Clavien_keras_prediction1)

Clavien_keras_roc<- roc(PCNL_Clavien_test_outcome, Clavien_keras_prediction1)

auc(Clavien_keras_roc)

confusionMatrix(Clavien_keras_tb)

plot(Clavien_keras_roc)

```
Area under the curve: 0.8891
Confusion Matrix and Statistics

                         Clavien_keras_prediction1
PCNL_Clavien_test_outcome    0    1
                        0 9037  245
                        1  259 1067

               Accuracy : 0.9525
                 95% CI : (0.9483, 0.9565)
    No Information Rate : 0.8763
    P-Value [Acc > NIR] : <2e-16

                  Kappa : 0.7818

 Mcnemar's Test P-Value : 0.5625

            Sensitivity : 0.9721
            Specificity : 0.8133
         Pos Pred Value : 0.9736
         Neg Pred Value : 0.8047
             Prevalence : 0.8763
         Detection Rate : 0.8519
   Detection Prevalence : 0.8750
      Balanced Accuracy : 0.8927

       'Positive' Class : 0
```

Clavien_keras_tb1<- table(PCNL_Clavien_test_outcome$V1, Clavien_keras_prediction1$V1)

Clavien_keras_roc1<- roc(PCNL_Clavien_test_outcome$V1, Clavien_keras_prediction1$V1)

auc(Clavien_keras_roc1)

confusionMatrix(Clavien_keras_tb1)

Area under the curve: 0.5036

Confusion Matrix and Statistics

0	1

0	2 243

1	1 1080

Accuracy : 0.816

95% CI : (0.7941, 0.8365)

No Information Rate : 0.9977

P-Value [Acc > NIR] : 1

Kappa : 0.0117

Mcnemar's Test P-Value : <2e-16

Sensitivity : 0.666667

Specificity : 0.816327

Pos Pred Value : 0.008163

Neg Pred Value : 0.999075

Prevalence : 0.002262

Detection Rate : 0.001508

Detection Prevalence : 0.184766

Balanced Accuracy : 0.741497

'Positive' Class : 0

Clavien_keras_tb2<- table(PCNL_Clavien_test_outcome$V2, Clavien_keras_prediction1$V2)

Clavien_keras_roc2<- roc(PCNL_Clavien_test_outcome$V2, Clavien_keras_prediction1$V2)

auc(Clavien_keras_roc2)

confusionMatrix(Clavien_keras_tb2)

Area under the curve: 0.4992

Confusion Matrix and Statistics

0	1

0 1237	2

1	87	0

Accuracy : 0.9329

95% CI : (0.9181, 0.9458)

No Information Rate : 0.9985

P-Value [Acc > NIR] : 1

Kappa : -0.003

Mcnemar's Test P-Value : <2e-16

Sensitivity : 0.9343

Specificity : 0.0000

Pos Pred Value : 0.9984

Neg Pred Value : 0.0000

Prevalence : 0.9985

Detection Rate : 0.9329

Detection Prevalence : 0.9344

Balanced Accuracy : 0.4671

'Positive' Class : 0

Clavien_keras_tb3<- table(PCNL_Clavien_test_outcome$V3, Clavien_keras_prediction1$V3)

Clavien_keras_roc3<- roc(PCNL_Clavien_test_outcome$V3, Clavien_keras_prediction1$V3)

auc(Clavien_keras_roc3)

confusionMatrix(Clavien_keras_tb3)

Area under the curve: 0.5

Confusion Matrix and Statistics

0	1

0 1216	0

1 110	0

Accuracy : 0.917

95% CI : (0.9009, 0.9313)

No Information Rate : 1

P-Value [Acc > NIR] : 1

Kappa : 0

Mcnemar's Test P-Value : <2e-16

Sensitivity : 0.917

Specificity : NA

Pos Pred Value : NA

Neg Pred Value : NA

Prevalence : 1.000

Detection Rate : 0.917

Detection Prevalence : 0.917

Balanced Accuracy : NA

'Positive' Class : 0

Clavien_keras_tb4<- table(PCNL_Clavien_test_outcome$V4, Clavien_keras_prediction1$V4)

Clavien_keras_roc4<- roc(PCNL_Clavien_test_outcome$V4, Clavien_keras_prediction1$V4)

auc(Clavien_keras_roc4)

confusionMatrix(Clavien_keras_tb4)

Area under the curve: 0.5

Confusion Matrix and Statistics

0	1

0 1301	0

1	25	0

Accuracy : 0.9811

95% CI : (0.9723, 0.9878)

No Information Rate : 1

P-Value [Acc > NIR] : 1

Kappa : 0

Mcnemar's Test P-Value : 1.587e-06

Sensitivity : 0.9811

Specificity : NA

Pos Pred Value : NA

Neg Pred Value : NA

Prevalence : 1.0000

Detection Rate : 0.9811

Detection Prevalence : 0.9811

Balanced Accuracy : NA

'Positive' Class : 0

Clavien_keras_tb5<- table(PCNL_Clavien_test_outcome$V5, Clavien_keras_prediction1$V5)

Clavien_keras_roc5<- roc(PCNL_Clavien_test_outcome$V5, Clavien_keras_prediction1$V5)

auc(Clavien_keras_roc5)

confusionMatrix(Clavien_keras_tb5)

Area under the curve: 0.5

Confusion Matrix and Statistics

0	1

0 1308	0

1	18	0

Accuracy : 0.9864

95% CI : (0.9786, 0.9919)

No Information Rate : 1

P-Value [Acc > NIR] : 1

Kappa : 0

Mcnemar's Test P-Value : 6.151e-05

Sensitivity : 0.9864

Specificity : NA

Pos Pred Value : NA

Neg Pred Value : NA

Prevalence : 1.0000

Detection Rate : 0.9864

Detection Prevalence : 0.9864

Balanced Accuracy : NA

'Positive' Class : 0

Clavien_keras_tb6<- table(PCNL_Clavien_test_outcome$V6, Clavien_keras_prediction1$V6)

Clavien_keras_roc6<- roc(PCNL_Clavien_test_outcome$V6, Clavien_keras_prediction1$V6)

auc(Clavien_keras_roc6)

confusionMatrix(Clavien_keras_tb6)

Area under the curve: 0.5

Confusion Matrix and Statistics

0	1

0 1323	0

1	3	0

Accuracy : 0.9977

95% CI : (0.9934, 0.9995)

No Information Rate : 1

P-Value [Acc > NIR] : 1.0000

Kappa : 0

Mcnemar's Test P-Value : 0.2482

Sensitivity : 0.9977

Specificity : NA

Pos Pred Value : NA

Neg Pred Value : NA

Prevalence : 1.0000

Detection Rate : 0.9977

Detection Prevalence : 0.9977

Balanced Accuracy : NA

'Positive' Class : 0

Clavien_keras_tb7<- table(PCNL_Clavien_test_outcome$V7, Clavien_keras_prediction1$V7)

Clavien_keras_roc7<- roc(PCNL_Clavien_test_outcome$V7, Clavien_keras_prediction1$V7)

auc(Clavien_keras_roc7)

confusionMatrix(Clavien_keras_tb7)

Area under the curve: 0.5

Confusion Matrix and Statistics

0	1

0 1325	0

1	1	0

Accuracy : 0.9992

95% CI : (0.9958, 1)

No Information Rate : 1

P-Value [Acc > NIR] : 1

Kappa : 0

Mcnemar's Test P-Value : 1

Sensitivity : 0.9992

Specificity : NA

Pos Pred Value : NA

Neg Pred Value : NA

Prevalence : 1.0000

Detection Rate : 0.9992

Detection Prevalence : 0.9992

Balanced Accuracy : NA

'Positive' Class : 0

Clavien_keras_tb8<- table(PCNL_Clavien_test_outcome$V8, Clavien_keras_prediction1$V8)

Clavien_keras_roc8<- roc(PCNL_Clavien_test_outcome$V8, Clavien_keras_prediction1$V8)

auc(Clavien_keras_roc8)

confusionMatrix(Clavien_keras_tb8)

Area under the curve: 0.5

Confusion Matrix and Statistics

0	1

0 1325	0

1	1	0

Accuracy : 0.9992

95% CI : (0.9958, 1)

No Information Rate : 1

P-Value [Acc > NIR] : 1

Kappa : 0

Mcnemar's Test P-Value : 1

Sensitivity : 0.9992

Specificity : NA

Pos Pred Value : NA

Neg Pred Value : NA

Prevalence : 1.0000

Detection Rate : 0.9992

Detection Prevalence : 0.9992

Balanced Accuracy : NA

'Positive' Class : 0

15. **Truncated Multiple output DL model**

##Outcomes: Transfusion, Infection + SF

#Sort Data for well predicted outcomes - Infection + Transfusion. Add in SF

PCNL_multiple_output_data_new<-na.omit(cbind(PCNL_factors_coded, Transfusion_coded3, Post_infection_coded3, SF_coded3))

PCNL_multiple_output_sample_new<-sample(1:nrow(PCNL_multiple_output_data_new), size=nrow(PCNL_multiple_output_data_new) *0.7)

PCNL_multiple_output_train_new<-PCNL_multiple_output_data_new[PCNL_multiple_output_sample_new,]

PCNL_multiple_output_test_new<-PCNL_multiple_output_data_new[-PCNL_multiple_output_sample_new,]

PCNL_multiple_output_train_predictors_new<-PCNL_multiple_output_train_new[,-44:-46]

PCNL_multiple_output_test_predictors_new<-PCNL_multiple_output_test_new[,-44:-46]

PCNL_multiple_output_train_outcomes_new<-PCNL_multiple_output_train_new[,-1:-43]

PCNL_multiple_output_test_outcomes_new<-PCNL_multiple_output_test_new[,-1:-43]

PCNL_multiple_output_train_predictors_new1<-as.matrix(PCNL_multiple_output_train_predictors_new)

PCNL_multiple_output_test_predictors_new1<-as.matrix(PCNL_multiple_output_test_predictors_new)

Transfusion_train_new<-as.matrix(PCNL_multiple_output_train_new$Transfusion_coded3)

Infection_train_new<-as.matrix(PCNL_multiple_output_train_new$Post_infection_coded3)

SF_train_new<-as.matrix(PCNL_multiple_output_train_new$SF_coded3)

Transfusion_test_new<-as.matrix(PCNL_multiple_output_test_new$Transfusion_coded3)

Infection_test_new<-as.matrix(PCNL_multiple_output_test_new$Post_infection_coded3)

SF_test_new<-as.matrix(PCNL_multiple_output_test_new$SF_coded3)

#Build model

library(keras)

library(tensorflow)

library(caret)

library(pROC)

PCNL_base_model_new<-layer_input(shape=c(43)) %>%

layer_dense(units=128, activation = "relu") %>%

layer_dense(units=128, activation = "relu") %>%

layer_dense(units=43, activation = "relu")

Transfusion_output_new<-PCNL_base_model_new %>%

layer_dense(units=1, activation="sigmoid")

Post_infection_output_new<-PCNL_base_model_new %>%

layer_dense(units=1, activation="sigmoid")

SF_output_new<-PCNL_base_model_new %>%

layer_dense(units=1, activation="sigmoid")

PCNL_multi_model_new<-keras_model(PCNL_base_model_new, list(Transfusion_output_new, Post_infection_output_new, SF_output_new))

PCNL_multi_model_new%>%compile(optimizer="rmsprop", loss=list("binary_crossentropy", "binary_crossentropy", "binary_crossentropy"), metrics="accuracy")

New_combined_model<-PCNL_multi_model_new%>%fit(PCNL_multiple_output_train_predictors_new1, list(Transfusion_train_new, Infection_train_new, SF_train_new), validation_split=0.2, epoch=200, batch_size=32, verbose=1)

New_results<-PCNL_multi_model_new%>%evaluate(PCNL_multiple_output_test_predictors_new1, list(Transfusion_test_new,Infection_test_new, SF_test_new),verbose=1)

New_combined_model_predictions<-PCNL_multi_model_new%>%predict(PCNL_multiple_output_test_predictors_new1,batch_size=32, verbose=1)

#Test model

New_combined_model_predictions1<-as.data.frame(New_combined_model_predictions)

#Transfusion

Transfusion_prediction_new<-New_combined_model_predictions1$structure.c.0.00262477993965149..0.0120222270488739..0.00148430466651917..

Transfusion_prediction_new1<-round(Transfusion_prediction_new, digits=0)

Transfusion_prediction_new1_tb<-table(Transfusion_test_new, Transfusion_prediction_new1)

confusionMatrix(Transfusion_prediction_new1_tb)

Transfusion_prediction_new1_roc<-roc(Transfusion_test_new, Transfusion_prediction_new1)

auc(Transfusion_prediction_new1_roc)

plot(Transfusion_prediction_new1_roc, col=5, lty=2)

```
Area under the curve: 0.8496

Confusion Matrix and Statistics

                    Transfusion_prediction_new1
Transfusion_test_new    0    1
                   0 1275    1
                   1   15   35

               Accuracy : 0.9879
                 95% CI : (0.9805, 0.9931)
    No Information Rate : 0.9729
    P-Value [Acc > NIR] : 0.0001309

                  Kappa : 0.8079

 Mcnemar's Test P-Value : 0.0011541

            Sensitivity : 0.9884
            Specificity : 0.9722
         Pos Pred Value : 0.9992
         Neg Pred Value : 0.7000
             Prevalence : 0.9729
         Detection Rate : 0.9615
   Detection Prevalence : 0.9623
      Balanced Accuracy : 0.9803

       'Positive' Class : 0
```

#Infection

Infection_predictions_new<-New_combined_model_predictions1$structure.c.0.290845274925232..0.150248736143112..0.0557928681373596..

Infection_predictions_new1<-round(Infection_predictions_new, digits=0)

Infection_prediction_new1_tb<-table(Infection_test_new, Infection_predictions_new1)

confusionMatrix(Infection_prediction_new1_tb)

Infection_prediction_new1_roc<-roc(Infection_test_new, Infection_predictions_new1)

auc(Infection_prediction_new1_roc)

plot(Infection_prediction_new1_roc, col=5, lty=2)

```
Area under the curve: 0.9048

Confusion Matrix and Statistics

                  Infection_predictions_new1
Infection_test_new   0   1
                 0 936  79
                 1  35 276

               Accuracy : 0.914
                 95% CI : (0.8976, 0.9286)
    No Information Rate : 0.7323
    P-Value [Acc > NIR] : < 2.2e-16

                  Kappa : 0.7718

 Mcnemar's Test P-Value : 5.642e-05

            Sensitivity : 0.9640
            Specificity : 0.7775
         Pos Pred Value : 0.9222
         Neg Pred Value : 0.8875
             Prevalence : 0.7323
         Detection Rate : 0.7059
   Detection Prevalence : 0.7655
      Balanced Accuracy : 0.8707

       'Positive' Class : 0
```

#SF

SF_predictions_new<-New_combined_model_predictions1$structure.c.0.271894574165344..0.0481393933296204..0.3408282995224..

SF_predictions_new1<-round(SF_predictions_new, digits=0)

SF_prediction_new1_tb<-table(SF_test_new, SF_predictions_new1)

confusionMatrix(SF_prediction_new1_tb)

SF_prediction_new1_roc<-roc(SF_test_new, SF_predictions_new1)

auc(SF_prediction_new1_roc)

plot(SF_prediction_new1_roc, col=5, lty=2)

```
Area under the curve: 0.5572

Confusion Matrix and Statistics

           SF_predictions_new1
SF_test_new    0    1
          0 1074   44
          1  176   32

               Accuracy : 0.8341
                 95% CI : (0.813, 0.8537)
    No Information Rate : 0.9427
    P-Value [Acc > NIR] : 1

                  Kappa : 0.1544

 Mcnemar's Test P-Value : <2e-16

            Sensitivity : 0.8592
            Specificity : 0.4211
         Pos Pred Value : 0.9606
         Neg Pred Value : 0.1538
             Prevalence : 0.9427
         Detection Rate : 0.8100
   Detection Prevalence : 0.8431
      Balanced Accuracy : 0.6401

       'Positive' Class : 0
```

16. ROC Curves for deep learning models

```{r Combine ROC Curves - Immediate Clearance}

#Immediate clearance

plot(Multi_Immediate_clearance_roc, col=4, lty=1, main=("Immediate Clearance on Fluoroscopy - Deep Neural Networks"), legend(x="right",legend=c("Blue: Multiple Output model", "AUC=0.533", "Green: Single Output model", "AUC=0.587")))

plot(immediate_clearance_keras_roc, col=3, lty=1, add=TRUE)

```

```{r Combine ROC Curves - Visceral Injury}

#Visceral Injury

plot(Multi_Visc_Injury_roc,col=4, lty=1, main="Visceral Injury on Fluoroscopy - Deep Neural Networks", legend(x="right", legend=c("Blue: Multiple Output model", " AUC=0.500", "Green: Single Output model", " AUC=0.500")))

plot(Visc_injury_keras_roc, col=3, lty=1, add=TRUE)

```

```{r Combine ROC Curves - Survival}

#Survival

#No Multi-output model ROC due to insufficient samples

plot(Survival_keras_roc,col=4, lty=1, main="Survival - Deep Neural Networks", legend(x="right", legend=c("Multiple Output model - N/A", "Blue: Single Output model", "AUC=0.499")))

```

```{r Combine ROC Curves - Transfusion}

#Transfusion

plot(Multi_Transfusion_roc,col=4, lty=1, main="Transfusion - Deep Neural Networks")

plot(Multi_Transfusion_roc,col=4, lty=1, main="Transfusion - Deep Neural Networks", legend(x="right", legend=c("Blue: Original Multiple Output model", " AUC=0.769", "Green: Single Output model", " AUC=0.874", "Purple: Truncated Multiple Output Model", " AUC=0.850")))

plot(Transfusion_keras_roc, col=3, lty=1, add=TRUE)

plot(Transfusion_prediction_new1_roc, col=6, lty=2, add=TRUE)

```

```{r Combine ROC Curves - Post-operative Infection}

#Post-operative Infection

plot(Multi_Post_Infection_roc,col=4, lty=1, main="Post-operative Infection - Deep Neural Networks")

plot(Multi_Post_Infection_roc,col=4, lty=1, main="Post-operative Infection - Deep Neural Networks", legend(x="right", legend=c("Blue: Original Multiple Output model", " AUC=0.905", "Green: Single Output model", " AUC=0.924","Purple: Truncated Multiple Output Model", " AUC=0.905")))

plot(Post_Infection_keras_roc, col=3, lty=1, add=TRUE)

plot(Infection_prediction_new1_roc, col=6, lty=2, add=TRUE)

```

```{r Combine ROC Curves - Intra-Operative Complications}

#Intra-operative complications

plot(Multi_Intra_Complications_roc,col=4, lty=1, main="Intra-Operative Complications - Deep Neural Networks")

plot(Multi_Intra_Complications_roc,col=4, lty=1, main="Intra-Operative Complications - Deep Neural Networks", legend(x="right", legend=c("Blue: Multiple Output model", " AUC=0.500", "Green: Single Output model", " AUC=0.500")))

plot(Intra_Complication_keras_roc, col=3, lty=1, add=TRUE)

```

```{r Combine ROC Curves - Need for ITU/HDU }

#Need for ITU/HDU

plot(Multi_ITU_HDU_roc, col=4, lty=1, main="Need for ITU/HDU - Deep Neural Networks")

plot(Multi_ITU_HDU_roc, col=1, lty=1, main="Need for ITU/HDU - Deep Neural Networks", legend(x="right", legend=c("Blue: Multiple Output model", " AUC=0.500", "Green: Single Output model", " AUC=0.500")))

plot(ITU_HDU_keras_roc, col=3, lty=1, add=TRUE)

```

```{r Combine ROC Curves - Stone Free Status at Follow-up}

#Stone Free status at follow-up

plot(Multi_SF_roc, col=4, lty=1, main="Stone Free Status at Follow-up - Deep Neural Networks")

plot(Multi_SF_roc, col=4, lty=1, main="Stone Free Status at Follow-up - Deep Neural Networks", legend(x="bottomright", legend=c("Blue: Original Multiple Output model", " AUC=0.599", "Green: Single Output model", " AUC=0.615", "Purple: Truncated Multiple Output Model", " AUC=0.557")))

plot(SF_keras_roc, col=3, lty=1, add=TRUE)

plot(SF_prediction_new1_roc , col=6, lty=2, add=TRUE)

```

```{r Combine ROC Curves - Need for Adjuvant Treatment}

#Need for Adjuvant treatment

plot(Multi_Adjuvant_roc, col=4, lty=1, main="Need for Adjuvant Treatment - Deep Neural Networks")

plot(Multi_Adjuvant_roc, col=4, lty=1, main="Need for Adjuvant Treatment - Deep Neural Networks", legend(x="right", legend=c("Blue: Multiple Output model", " AUC=0.500", "Green: Single Output model", " AUC=0.530")))

plot(Adjuvant_keras_roc, col=3, lty=1, add=TRUE)

```

```{r Combine ROC Curves - Post-Operative Stay - Overall Predictions}

#Post-operative stay (coded)

###Post-op stay - overall

plot(Multi_Post_op_stay_roc, col=4, lty=1, main="Post-Operative Stay - Overall Predictions - Deep Neural Networks")

plot(Multi_Post_op_stay_roc, col=4, lty=1, main="Post-Operative Stay - Overall Predictions - Deep Neural Networks", legend(x="right", legend=c("Blue: Multiple Output model", " AUC=0.756", "Green: Single Output model", " AUC=0.623")))

plot(Post_Stay_keras_roc, col=3, lty=1, add=TRUE)

```

```{r Combine ROC Curves - Post-op stay - individual predictions}

###Post-op stay - individual predictions

plot(Multi_Post_op_stay_roc1, col=1, lty=1, main="Post-Operative Stay - Individual Predictions - Deep Neural Networks", legend(x="right", legend=c("Multiple Output model", "AUC=", "Single Output model", "AUC=")))

plot(Multi_Post_op_stay_roc2, col=2, lty=1, add=TRUE)

plot(Multi_Post_op_stay_roc3, col=3, lty=1, add=TRUE)

plot(Multi_Post_op_stay_roc4, col=4, lty=1, add=TRUE)

```

```{r Combine ROC Curves - CD classification - overall predictions}

#Clavien grade of complication

###Clavien - overall

plot(Multi_Clavien_roc, col=4, lty=1, main="Clavien Dindo Classification - Overall Predictions - Deep Neural Networks")

plot(Multi_Clavien_roc, col=4, lty=1, main="Clavien Dindo Classification - Overall Predictions - Deep Neural Networks", legend(x="right", legend=c("Blue: Multiple Output model", " AUC=0.892", "Green: Single Output model", " AUC=0.889")))

plot(Clavien_keras_roc, col=3, lty=1, add=TRUE)

```

```{r Combine ROC Curves - CD classification - individual predictions}

###Clavien - individual predictions

plot(Multi_Clavien_roc1, col=1, lty=1, main="Clavien Dindo Classification - Overall Predictions - Deep Neural Networks", legend(x="right", legend=c("Multiple Output model", "AUC=0.533", "Single Output model", "AUC=")))

plot(Multi_Clavien_roc2, col=2, lty=1, add=TRUE)

plot(Multi_Clavien_roc3, col=3, lty=1, add=TRUE)

plot(Multi_Clavien_roc4, col=4, lty=1, add=TRUE)

plot(Multi_Clavien_roc5, col=5, lty=1, add=TRUE)

plot(Multi_Clavien_roc6, col=6, lty=1, add=TRUE)

plot(Multi_Clavien_roc8, col=7, lty=1, add=TRUE)

```
