## Supplementary Tables for "Use of Internally Validated Machine and Deep Learning Models to Predict Outcomes of Percutaneous Nephrolithotomy using data from the BAUS PCNL audit"

| **Variable** | | **Overall dataset (n=4418)** | **Training set (n=3092)*** | **Test set (n=1326)*** | **P (95% CI – if available) [Training vs Test set]** |
| --- | --- | --- | --- | --- | --- |
| **Age, mean±SD** | | 56.5 ± 19.4 | 56.3 ± 19.4 | 56.8 ± 19.4 | 0.44 (-1.74 to 0.76) |
| **Female, n (%)** | | 2074 (46%) | 1461 (47%) | 613 (46%) | 0.55 |
| **BMI, median (IQR)** | | 28.4 (25.0, 33.0) | 28.3 (25.0, 33.0) | 28.5 (24.7, 33.1) | 0.73 |
| **Pre-Operative Haemoglobin (g/L), mean±SD** | | 135.6 ± 16.6 | 135.6 ± 16.7 | 135.7 ± 16.4 | 0.77 (-1.22 to 0.90) |
| **Side, n (%)** | **Right** | 2,391 (54%) | 1666 (54%) | 725 (55%) | 0.86 |
|  | **Left** | 1,956 (44%) | 1377 (45%) | 579 (44%) |  |
|  | **Bilateral** | 71 (2%) | 49 (2%) | 22 (2%) |  |
| **Charlson Score, median (IQR)** | | 1 (1, 2) | 0 (0, 1) | 0 (0, 1) | 0.82 |
| **Age Related Charlson Score, median (IQR)** | | 0 (0, 3) | 0 (0, 3) | 0 (0, 3) | 0.94 |
| **Number Of Tracts Planned, median (IQR)** | | 1 (1, 2) | 1 (1, 2) | 1 (1, 2) | 0.75 |
| **Previous UTI, n (%)** | **Ia - one or two oral antibiotic courses** | 743 (17%) | 525 (17%) | 218 (16%) | 0.60 |
|  | **Ib - three or more oral antibiotic courses** | 628 (14%) | 429 (14%) | 199 (15%) |  |
|  | **IIa - an inpatient admission for IV antibiotics** | 190 (4%) | 136 (4%) | 54 (4%) |  |
|  | **IIb - two or more inpatient admissions for IV antibiotics** | 100 (2%) | 65 (2%) | 35 (3%) |  |
|  | **IIIa - a JJ stent or nephrostomy tube for drainage of infected system** | 364 (8%) | 258 (8%) | 106 (8%) |  |
|  | **IIIb - as above; with patient remaining in hospital until definitive procedure performed** | 22 (0.5%) | 16 (1%) | 6 (0.5%) |  |
|  | **IVa - an ITU admission for sepsis for IV antibiotics and close monitoring** | 46 (1%) | 28 (1%) | 18 (1%) |  |
|  | **IVb - an ITU admission for sepsis requiring inotropic support** | 40 (1%) | 32 (1%) | 8 (1%) |  |
|  | **None** | 2285 (52%) | 1603 (52%) | 682 (51%) |  |
| **Previous UTI, n (%)** | | 2133 (48%) | 1489 (48%) | 644 (48%) | 0.83 |
| **Prior Antibiotics, n (%)** | **None** | 3204 (73%) | 2278 (73%) | 936 (71%) | 0.56 |
|  | **Prophylactic** | 164 (4%) | 110 (4%) | 54 (4%) |  |
|  | **Oral course (1-3 days)** | 205 (5%) | 138 (4%) | 67 (5%) |  |
|  | **Oral course (>3 days)** | 559 (13%) | 384 (12%) | 175 (13%) |  |
|  | **IV pre-induction** | 13 (0.5%) | 9 (0.5%) | 4 (0.5%) |  |
|  | **IV course** | 273 (6%) | 183 (6%) | 90 (7%) |  |
| **Prior Antibiotics, n (%)** | | 1214 (27%) | 814 (26%) | 390 (29%) | 0.75 |
| **Pre-Operative MSU, n (%)** | | 3993 (90%) | 2808 (91%) | 1185 (89%) | 0.15 |
| **Pre-Operative MSU Result, n (%)** | **No growth** | 2685 (61%) | 1921 (62%) | 764 (58%) | 0.07 |
|  | **Not Done** | 493 (11%) | 327 (11%) | 166 (13%) |  |
|  | **Heavy mixed growth untreated** | 246 (6%) | 170 (5%) | 76 (6%) |  |
|  | **Heavy mixed growth treated pre-operatively** | 128 (3%) | 85 (3%) | 43 (3%) |  |
|  | **UTI untreated** | 80 (2%) | 59 (2%) | 21 (2%) |  |
|  | **UTI treated pre-operatively** | 786 (18%) | 530 (17%) | 256 (19%) |  |
| **Pre-operative Imaging, n (%)** | **CTU** | 1006 (23%) | 689 (22%) | 317 (24%) | 0.41 |
|  | **IVU** | 12 (0.3%) | 8 (0.3%) | 4 (0.3%) |  |
|  | **NCCT** | 3118 (71%) | 2204 (71%) | 914 (69%) |  |
|  | **US** | 16 (0.4%) | 13 (0.4%) | 3 (0.2%) |  |
|  | **XR** | 245 (6%) | 166 (5%) | 79 (6%) |  |
|  | **XR + US** | 21 (0.5%) | 12 (0.4%) | 9 (0.7%) |  |
| **Pre-Op eGFR, n (%)** | **<15** | 18 (0.4%) | 13 (0.4%) | 5 (0.4%) | 0.43 |
|  | **15-29** | 131 (3%) | 100 (3%) | 31 (2%) |  |
|  | **30-44** | 295 (7%) | 203 (7%) | 92 (7%) |  |
|  | **45-49** | 540 (12%) | 367 (12%) | 173 (13%) |  |
|  | **≥60** | 3434 (78%) | 2409 (78%) | 1025 (77%) |  |
| **Abx on Induction, n (%)** | | 4259 (96%) | 2978 (96%) | 1281 (97%) | 0.70 |
| **Type of Anaesthesia, n (%)** | **General** | 4411 (99.8%) | 3088 (99.9%) | 1323 (99.8%) | 0.43 |
|  | **Spinal** | 6 (0.1%) | 4 (0.1%) | 2 (0.2%) |  |
|  | **Local** | 1 (0.02%) | 0 | 1 (0.08%) |  |
| **IR Back-Up Available, n (%)** | | 3490 (79%) | 2446 (79%) | 1044 (79%) | 0.81 |
| **Relook Nephroscopy, n (%)** | | 8 (0.2%) | 5 (0.2%) | 3 (0.2%) | 0.70 |
| **Stone Dimensions, n (%)** | **0-1cm** | 177 (4%) | 130 (4%) | 47 (4%) | 0.09 |
|  | **1-2cm** | 1160 (27%) | 805 (26%) | 355 (27%) |  |
|  | **>2cm** | 302 (7%) | 195 (6%) | 107 (8%) |  |
|  | **2-4cm** | 1881 (43%) | 1344 (43%) | 537 (40%) |  |
|  | **>4cm** | 898 (20%) | 618 (20%) | 280 (21%) |  |
| **No. Stones, n (%)** | **Single** | 602 (86%) | 423 (86%) | 179 (87%) | 0.91 |
|  | **Multiple** | 3816 (14%) | 2669 (14%) | 1147 (13%) |  |
| **Stone Location, n (%)** | **Calyceal Diverticular** | 49 (1%) | 32 (1%) | 17 (1%) | 0.52 |
|  | **Lower Calyx** | 864 (20%) | 605 (20%) | 259 (20%) |  |
|  | **Middle Calyx** | 148 (3%) | 102 (3%) | 46 (3%) |  |
|  | **Upper Calyx** | 236 (5%) | 169 (5%) | 67 (5%) |  |
|  | **Upper ureteric** | 141 (3%) | 94 (3%) | 47 (3%) |  |
|  | **Other ureteric** | 33 (1%) | 22 (1%) | 11 (1%) |  |
|  | **Pelvic** | 1463 (33%) | 1046 (34%) | 417 (31%) |  |
|  | **Partial Staghorn** | 822 (19%) | 581 (19%) | 241 (18%) |  |
|  | **Complete Staghorn** | 662 (15%) | 441 (14%) | 221 (17%) |  |
| **GSS (Stone Complexity)** | **I** | 338 (8%) | 254 (8%) | 84 (6%) | 0.21 |
|  | **II** | 1908 (43%) | 1339 (43%) | 569 (43%) |  |
|  | **III** | 1439 (33%) | 1005 (33%) | 434 (33%) |  |
|  | **IV** | 733 (17%) | 494 (16%) | 239 (18%) |  |
| **Max Hounsfield Units on CTKUB, n (%)** | **Not Recorded** | 2020 (46%) | 1413 (46%) | 607 (46%) | 0.22 |
|  | **<500 HU** | 177 (4%) | 123 (4%) | 54 (4%) |  |
|  | **501-1000 HU** | 834 (19%) | 590 (19%) | 244 (18%) |  |
|  | **1001-1500 HU** | 1065 (24%) | 745 (24%) | 320 (24%) |  |
|  | **>1500 HU** | 322 (7%) | 221 (7%) | 101 (8%) |  |
| **Pre-Existing PCN, n (%)** | | 491 (11%) | 349 (11%) | 142 (11%) | 0.61 |
| **Puncture Performed By, n (%)** | **Urologist** | 1723 (39%) | 1209 (39%) | 514 (39%) | 0.86 |
|  | **Radiologist** | 2695 (61%) | 1883 (61%) | 812 (61%) |  |
| **Puncture Site, n (%)** | **Interpolar** | 568 (13%) | 397 (13%) | 171 (13%) | 0.44 |
|  | **Lower Pole** | 2167 (49%) | 1535 (50%) | 632 (48%) |  |
|  | **Upper Pole** | 1683 (38%) | 1160 (38%) | 523 (39%) |  |
| **Image Guidance, n (%)** | **Fluoroscopy & Ultrasound** | 2556 (58%) | 1779 (58%) | 777 (59%) | 0.66 |
|  | **Fluoroscopy alone** | 1638 (37%) | 1149 (37%) | 489 (37%) |  |
|  | **Ultrasound only** | 217 (5%) | 158 (5%) | 49 (4%) |  |
|  | **CT only** | 7 (0%) | 6 (0%) | 1 (0%) |  |
| **Patient Position, n (%)** | **Prone** | 2857 (65%) | 2001 (65%) | 856 (65%) | 0.20 |
|  | **Supine** | 1515 (34%) | 1061 (34%) | 454 (34%) |  |
|  | **Other** | 46 (1%) | 30 (1%) | 16 (1%) |  |
| **Tract Placement, n (%)** | **Subcostal** | 3949 (89%) | 2765 (89%) | 1184 (89%) | 0.94 |
|  | **Supracostal** | 469 (11%) | 327 (11%) | 142 (11%) |  |
| **Size Outer Diameter Amplatz Sheath** | **<6Fr** | 2 (0 %) | 0 | 2 (0%) | 0.32 |
|  | **6-10Fr** | 8 (0%) | 5 (0%) | 3 (0%) |  |
|  | **11-14Fr** | 70 (2%) | 48 (2%) | 22 (2%) |  |
|  | **15-18Fr** | 412 (9%) | 294 (10%) | 118 (9%) |  |
|  | **19-24Fr** | 528 (12%) | 382 (12%) | 146 (11%) |  |
|  | **25-26Fr** | 283 (6%) | 196 (6%) | 87 (7%) |  |
|  | **27-30Fr** | 3115 (71%) | 2167 (70%) | 948 (71%) |  |
| **Dilators Used, n (%)** | **Balloon** | 2802 (63%) | 1956 (63%) | 846 (64%) | 0.89 |
|  | **Metal** | 1479 (33%) | 1038 (34%) | 441 (33%) |  |
|  | **Teflon** | 137 (3%) | 98 (3%) | 39 (3%) |  |
| **Predicted Difficulty, n (%)** | **None** | 3042 (77%) | 2393 (77%) | 1009 (76%) | 0.80 |
|  | **Renal Anatomy** | 902 (20%) | 632 (20%) | 270 (20%) |  |
|  | **Patient Anatomy** | 114 (3%) | 67 (2%) | 47 (4%) |  |
| **Accessory Procedures, n (%)** | **None** | 1041 (24%) | 731 (24%) | 310 (23%) | 0.46 |
|  | **Stent insertion only** | 685 (16%) | 483 (16%) | 202 (15%) |  |
|  | **Ureteric catheter only** | 188 (4%) | 128 (4%) | 60 (5%) |  |
|  | **Stent insertion & Ureteric catheter** | 17 (0%) | 10 (0%) | 7 (1%) |  |
|  | **Flexible Renoscopy** | 1661 (38%) | 1153 (37%) | 508 (38%) |  |
|  | **Flexible ureteroscopy** | 335 (8%) | 226 (7%) | 109 (8%) |  |
|  | **Flexible Renoscopy and Flexible Ureteroscopy** | 491 (11%) | 361 (12%) | 130 (10%) |  |
| **Nephrostomy Drain in Situ, n (%)** | | 3047 (69%) | 2133 (69%) | 914 (69%) | 0.99 |
| **Stone Extraction/Fragmentation Technique, n (%)** | **Ultrasonic lithotripter** | 1508 (34%) | 1074 (35%) | 434 (33%) | 0.59 |
|  | **Lithoclast** | 1047 (24%) | 726 (23%) | 321 (24%) |  |
|  | **Laser** | 969 (22%) | 677 (22%) | 292 (22%) |  |
|  | **Lift out** | 894 (20%) | 615 (20%) | 279 (21%) |  |

Table 1. All predictor variables for overall dataset, training and test sets. P-values relate to training vs test sets

| **Immediate Clearance** | **ML (all Single Outcome)** | | | | | | **Deep NN** | |
| --- | --- | --- | --- | --- | --- | --- | --- | --- |
|  | **Random Forests** | **Partitioning** | **XGBoost** | **Logistic Regression** | **Neural Network** | **Bayesian Generalised Linear Model** | **Single Outcome** | **Multiple Outcome** |
| **AUC** | 0.73 | 0.63 | 0.75 | 0.63 | 0.74 | 0.75 | 0.59 | 0.53 |
| **Overall accuracy (95% CI)** | 0.77 (0.74-0.79) | 0.71 (0.69-0.74) | 0.77 (0.75-0.80) | 0.76 (0.73-0.78) | 0.75 (0.73-0.78) | 0.77 (0.75-0.80) | 0.77 (0.75-0.79) | 0.77 (0.75-0.79) |
| **Sensitivity** | 0.27 | 0.22 | 0.27 | 0.12 | 0.23 | 0.29 | 0.57 | 0.6 |
| **Specificity** | 0.94 | 0.89 | 0.94 | 0.97 | 0.93 | 0.94 | 0.79 | 0.78 |
| **PPV** | 0.58 | 0.43 | 0.61 | 0.56 | 0.52 | 0.60 | 0.23 | 0.084 |
| **NPV** | 0.79 | 0.76 | 0.79 | 0.77 | 0.78 | 0.80 | 0.95 | 0.98 |
| **N with outcome/Total N (Training group)** | 2360/3092 | | | | | | | |
| **N with outcome/Total N (Test group)** | 1015/1326 | | | | | | | |

Table 2. Diagnostic accuracy statistics for immediate clearance.

| **Visceral Injury** | **ML (all Single Outcome)** | | | | | | **Deep NN** | |
| --- | --- | --- | --- | --- | --- | --- | --- | --- |
|  | **Random Forests** | **Partitioning** | **XGBoost** | **Logistic Regression** | **Neural Network** | **Bayesian Generalised Linear Model** | **Single Outcome** | **Multiple Outcome** |
| **AUC** | 0.72 | 0.46 | 0.66 | 0.56 | 0.47 | 0.77 | 0.50 | 0.5 |
| **Overall accuracy (95% CI)** | 0.99 (0.99-1.00) | 0.99 (0.99-1.00) | 0.99 (0.99-1.00) | 0.99 (0.99-1.00) | 0.99 (0.99-1.00) | 0.99 (0.99-1.00) | 0.99 (0.99-1.00) | 0.99 (0.99-1.00) |
| **Sensitivity** | 1.00 | 1.00 | 1.00 | 1.00 | 1.00 | 1.00 | 0.99 | 0.99 |
| **Specificity** | 0.00 | 0.00 | 0.00 | 0.00 | 0.00 | 0.00 | N/A | N/A |
| **PPV** | 0.99 | 0.99 | 0.99 | 0.99 | 0.99 | 0.99 | N/A | N/A |
| **NPV** | N/A | N/A | N/A | N/A | N/A | N/A | N/A | N/A |
| **N with outcome/Total N (Train group)** | 11/3092 | | | | | | | |
| **N with outcome/Total N (Test group)** | 5/1326 | | | | | | | |

Table 3. Diagnostic accuracy statistics for Visceral Injury

| **Survival** | **ML (all Single Outcome)** | | | | | | **Deep NN** |
| --- | --- | --- | --- | --- | --- | --- | --- |
|  | **Random Forests** | **Partitioning** | **XGBoost** | **Logistic Regression** | **Neural Network** | **Bayesian Generalised Linear Model** | **Single Outcome** |
| **AUC** | 0.72 | 0.46 | 0.55 | 0.55 | 0.46 | 0.77 | 0.5 |
| **Overall accuracy (95% CI)** | 0.99 (0.99-1.00) | 0.99 (0.99-1.00) | 0.99 (0.99-1.00) | 0.99 (0.99-1.00) | 0.99 (0.99-1.00) | 0.99 (0.99-1.00) | 0.99 (0.99-1.00) |
| **Sensitivity** | 1 | 1 | 1 | 1 | 1 | 1 | 1 |
| **Specificity** | 0 | 0 | 0 | 0 | 0 | 0 | 0 |
| **PPV** | 0.99 | 0.99 | 0.99 | 0.99 | 0.99 | 0.99 | 0.99 |
| **NPV** | N/A | N/A | N/A | N/A | N/A | N/A | 0 |
| **N with outcome/Total N (Training group)** | 11/3092 | | | | | | |
| **N with outcome/Total N (Test group)** | 1/1326 | | | | | | |

Table 4. Diagnostic accuracy statistics for Survival

| **Intra-operative Complication** | **ML (all Single Outcome)** | | | | | | **Deep NN** | |
| --- | --- | --- | --- | --- | --- | --- | --- | --- |
|  | **Random Forests** | **Partitioning** | **XGBoost** | **Logistic Regression** | **Neural Network** | **Bayesian Generalised Linear Model** | **Single Outcome** | **Multiple Outcome** |
| **AUC** | 0.72 | 0.56 | 0.69 | 0.58 | 0.60 | 0.72 | 0.50 | 0.50 |
| **Overall accuracy (95% CI)** | 0.96 (0.95-0.97) | 0.96 (0.95-0.97) | 0.96 (0.95-0.97) | 0.96 (0.95-0.97) | 0.96 (0.95-0.97) | 0.96 (0.95-0.97) | 0.96 (0.95-0.97) | 0.97 (0.95-0.97) |
| **Sensitivity** | 1.00 | 1.00 | 1.00 | 1.00 | 1.00 | 1.00 | 0.96 | 0.97 |
| **Specificity** | 0.00 | 0.00 | 0.00 | 0.04 | 0.00 | 0.02 | N/A | N/A |
| **PPV** | 0.96 | 0.96 | 0.96 | 0.96 | 0.96 | 0.96 | N/A | N/A |
| **NPV** | N/A | N/A | N/A | 0.50 | N/A | 1.00 | N/A | N/A |
| **N with outcome/Total N (Training group)** | 93/2840 | | | | | | | |
| **N with outcome/Total N (Test group)** | 47/1218 | | | | | | | |

Table 5. Diagnostic accuracy statistics for intra-operative complication

| **Need for HDU/ITU** | **ML (all Single Outcome)** | | | | | | **Deep NN** | |
| --- | --- | --- | --- | --- | --- | --- | --- | --- |
|  | **Random Forests** | **Partitioning** | **XGBoost** | **Logistic Regression** | **Neural Network** | **Bayesian Generalised Linear Model** | **Single Outcome** | **Multiple Outcome** |
| **AUC** | 0.63 | 0.54 | 0.60 | 0.52 | 0.49 | 0.54 | 0.50 | 0.50 |
| **Overall accuracy (95% CI)** | 0.94 (0.93-0.95) | 0.94 (0.93-0.95) | 0.94 (0.93-0.95) | 0.94 (0.93-0.95) | 0.94 (0.93-0.95) | 0.94 (0.93-0.95) | 0.94 (0.93-0.95) | 0.95 (0.94-0.96) |
| **Sensitivity** | 1.00 | 1.00 | 1.00 | 1.00 | 1.00 | 1.00 | 0.94 | 0.95 |
| **Specificity** | 0.00 | 0.00 | 0.00 | 0.00 | 0.00 | 0.00 | N/A | N/A |
| **PPV** | 0.94 | 0.94 | 0.94 | 0.94 | 0.94 | 0.94 | N/A | N/A |
| **NPV** | N/A | N/A | N/A | 0.00 | N/A | N/A | N/A | N/A |
| **N with outcome/Total N (Training group)** | 128/2940 | | | | | | | |
| **N with outcome/Total N (Test group)** | 73/1260 | | | | | | | |

Table 6. Diagnostic accuracy statistics for need for ITU/HDU

| **Stone Free at Follow-up** | **ML (all Single Outcome)** | | | | | | **Deep NN** | | |
| --- | --- | --- | --- | --- | --- | --- | --- | --- | --- |
|  | **Random Forests** | **Partitioning** | **XGBoost** | **Logistic Regression** | **Neural Network** | **Bayesian Generalised Linear Model** | **Single Outcome** | **Multiple Outcome** | **Truncated Multiple Outcome** |
| **AUC** | 0.69 | 0.55 | 0.70 | 0.61 | 0.50 | 0.67 | 0.62 | 0.60 | 0.56 |
| **Overall accuracy (95% CI)** | 0.70 (0.64-0.74) | 0.70 (0.64-0.74) | 0.65 (0.60-0.70) | 0.62 (0.56-0.67) | 0.70 (0.64-0.74) | 0.69 (0.63-0.74) | 0.59 (0.54-0.65) | 0.44 (0.41-0.46) | 0.83 (0.81-0.85) |
| **Sensitivity** | 0.00 | 0.00 | 0.2 | 0.30 | 0.00 | 0.30 | 0.43 | 0.92 | 0.86 |
| **Specificity** | 1.00 | 1.00 | 0.87 | 0.78 | 1.00 | 0.88 | 0.78 | 0.21 | 0.42 |
| **PPV** | N/A | N/A | 0.44 | 0.40 | N/A | 0.54 | 0.69 | 0.35 | 0.96 |
| **NPV** | 0.70 | 0.70 | 0.69 | 0.69 | 0.70 | 0.72 | 0.55 | 0.85 | 0.15 |
| **N with outcome/Total N (Training group)** | 535/778 | | | | | | | | |
| **N with outcome/Total N (Test group)** | 228/328 | | | | | | | | |

Table 7. Diagnostic accuracy statistics for Stone free at follow-up

| **Need for Adjuvant treatment** | **ML (all Single Outcome)** | | | | | | **Deep NN** | |
| --- | --- | --- | --- | --- | --- | --- | --- | --- |
|  | **Random Forests** | **Partitioning** | **XGBoost** | **Logistic Regression** | **Neural Network** | **Bayesian Generalised Linear Model** | **Single Outcome** | **Multiple Outcome** |
| **AUC** | 0.69 | 0.61 | 0.67 | 0.59 | 0.49 | 0.67 | 0.53 | 0.5 |
| **Overall accuracy (95% CI)** | 0.82 (0.77-0.86) | 0.83 (0.78-0.87) | 0.82 (0.78-0.86) | 0.79 (0.75-0.84) | 0.82 (0.78-0.86) | 0.83 (0.78-0.87) | 0.83 (0.78-0.87) | 0.96 (0.95-0.97) |
| **Sensitivity** | 0.99 | 0.98 | 1.00 | 0.94 | 1.00 | 0.98 | 0.83 | 0.96 |
| **Specificity** | 0.02 | 0.12 | 0.02 | 0.14 | 0.00 | 0.14 | 0.67 | N/A |
| **PPV** | 0.82 | 0.84 | 0.82 | 0.83 | 0.82 | 0.84 | 0.99 | N/A |
| **NPV** | 0.33 | 0.58 | 0.50 | 0.32 | N/A | 0.57 | 0.07 | N/A |
| **N with outcome/Total N (Training group)** | 126/771 | | | | | | | |
| **N with outcome/Total N (Test group)** | 59/331 | | | | | | | |

Table 8. Diagnostic accuracy statistics for need for adjuvant treatment

| **Post-operative stay** | | **ML (all Single Outcome)** | | | | | | **Deep NN** | |
| --- | --- | --- | --- | --- | --- | --- | --- | --- | --- |
|  |  | **Random Forests** | **Partitioning** | **XGBoost** | **Logistic Regression** | **Neural Network** | **Bayesian Generalised Linear Model** | **Single Outcome** | **Multiple Outcome** |
| **AUC** | | 0.79 | 0.68 | 0.78 | 0.75 | 0.75 | 0.73 | 0.62 | 0.76 |
| **Overall accuracy (95% CI)** | | 0.66 (0.63-0.69) | 0.66 (0.63-0.68) | 0.68 (0.65-0.70) | 0.67 (0.64-0.69) | 0.68 (0.65-0.70) | 0.56 (0.53-0.59) | 0.72 (0.71-0.73) | 0.82 (0.81-0.83) |
| **Daycase** | **Sensitivity** | 0.00 | 0.00 | 0.00 | 0.00 | 0.00 | 0.00 | 0.98 | 0.98 |
|  | **Specificity** | 1.00 | 1.00 | 1.00 | 1.00 | 1.00 | 1.00 | N/A | N/A |
|  | **PPV** | N/A | N/A | N/A | N/A | N/A | N/A | N/A | N/A |
|  | **NPV** | 0.99 | 0.99 | 0.99 | 0.99 | 0.99 | 0.99 | N/A | N/A |
| **1 day** | **Sensitivity** | 0.70 | 0.73 | 0.75 | 0.77 | 0.72 | 0.007 | 0.82 | 0.95 |
|  | **Specificity** | 0.91 | 0.88 | 0.90 | 0.88 | 0.90 | 1.00 | 0.57 | 0.45 |
|  | **PPV** | 0.67 | 0.60 | 0.66 | 0.60 | 0.65 | 0.67 | 0.97 | 0.75 |
|  | **NPV** | 0.92 | 0.93 | 0.93 | 0.94 | 0.93 | 0.80 | 0.15 | 0.83 |
| **2 days** | **Sensitivity** | 0.073 | 0.00 | 0.00 | 0.0070 | 0.00 | 0.00 | 0.76 | 0.80 |
|  | **Specificity** | 0.94 | 1.00 | 0.99 | 0.99 | 1.00 | 1.00 | N/A | 0.25 |
|  | **PPV** | 0.26 | N/A | 0.00 | 0.20 | N/A | 0.00 | N/A | 0.62 |
|  | **NPV** | 0.77 | 0.77 | 0.77 | 0.77 | 0.77 | 0.77 | N/A | 0.46 |
| **a days** | **Sensitivity** | 0.89 | 0.91 | 0.94 | 0.90 | 0.94 | 0.99 | 0.81 | 0.46 |
|  | **Specificity** | 0.50 | 0.45 | 0.46 | 0.47 | 0.44 | 0.003 | 0.57 | 0.88 |
|  | **PPV** | 0.70 | 0.68 | 0.69 | 0.69 | 0.68 | 0.56 | 0.10 | 0.99 |
|  | **NPV** | 0.79 | 0.79 | 0.85 | 0.78 | 0.86 | 0.50 | 0.98 | 0.08 |
| **Total N (Training group)** | | 3092 | | | | | | | |
| **Total N (Test group)** | | 1326 | | | | | | | |

Table 9. Diagnostic accuracy statistics for post-operative stay duration

| **Clavien Dindo classification** | | **ML (all Single Outcome)** | | | | | | **Deep NN** | |
| --- | --- | --- | --- | --- | --- | --- | --- | --- | --- |
|  |  | **Random Forests** | **Partitioning** | **XGBoost** | **Logistic Regression** | **Neural Network** | **Bayesian Generalised Linear Model** | **Single Outcome** | **Multiple Outcome** |
| **AUC** | | 0.90 | 0.66 | 0.88 | 0.82 | 0.50 | 0.79 | 0.89 | 0.89 |
| **Overall accuracy (95% CI)** | | 0.85 (0.83-0.87) | 0.83 (0.81-0.85) | 0.85 (0.83-0.87) | 0.86 (0.84-0.88) | 0.82 (0.80-0.84) | 0.82 (0.80-0.84) | 0.95 (0.95-0.96) | 0.95 (0.95-0.96) |
| **Zero** | **Sensitivity** | 0.97 | 1.00 | 0.99 | 0.99 | 1.00 | 1.00 | 0.67 | 0.60 |
|  | **Specificity** | 0.40 | 0.15 | 0.32 | 0.17 | 0.00 | 0.00 | 0.82 | 0.81 |
|  | **PPV** | 0.88 | 0.84 | 0.87 | 0.88 | 0.82 | 0.82 | 0.008 | 0.01 |
|  | **NPV** | 0.77 | 0.90 | 0.85 | 0.70 | N/A | N/A | 1.00 | 0.99 |
| **I** | **Sensitivity** | 0.30 | 0.07 | 0.20 | 0.00 | 0.00 | 0.00 | 0.93 | 0.92 |
|  | **Specificity** | 0.98 | 1.00 | 0.99 | 1.00 | 1.00 | 1.00 | 0.00 | N/A |
|  | **PPV** | 0.57 | 0.75 | 0.71 | 0.00 | N/A | N/A | 1.00 | N/A |
|  | **NPV** | 0.95 | 0.94 | 0.95 | 0.95 | 0.93 | 0.93 | 0.00 | N/A |
| **II** | **Sensitivity** | 0.32 | 0.16 | 0.31 | 0.22 | 0.00 | 0.00 | 0.92 | 0.93 |
|  | **Specificity** | 0.97 | 0.99 | 0.98 | 0.98 | 1.00 | 1.00 | N/A | N/A |
|  | **PPV** | 0.45 | 0.50 | 0.51 | 0.43 | N/A | N/A | N/A | N/A |
|  | **NPV** | 0.95 | 0.93 | 0.94 | 0.95 | 0.92 | 0.92 | N/A | N/A |
| **IIIa** | **Sensitivity** | 0.19 | 0.00 | 0.07 | 0.00 | 0.00 | 0.00 | 0.98 | 0.98 |
|  | **Specificity** | 1.00 | 1.00 | 1.00 | 1.00 | 1.00 | 1.00 | N/A | 0.00 |
|  | **PPV** | 1.00 | N/A | 0.67 | N/A | N/A | N/A | N/A | 1.00 |
|  | **NPV** | 0.98 | 0.97 | 0.98 | 0.99 | 0.98 | 0.98 | N/A | 0.00 |
| **IIIb** | **Sensitivity** | 0.12 | 0.00 | 0.00 | 0.00 | 0.00 | 0.00 | 0.99 | 0.98 |
|  | **Specificity** | 1.00 | 1.00 | 1.00 | 1.00 | 1.00 | 1.00 | N/A | N/A |
|  | **PPV** | 1.00 | N/A | 0.00 | N/A | N/A | N/A | N/A | N/A |
|  | **NPV** | 0.99 | 0.99 | 0.98 | 0.99 | 0.99 | 0.99 | N/A | N/A |
| **IVa** | **Sensitivity** | 0.00 | 0.00 | 0.00 | 0.00 | 0.00 | 0.00 | 1.00 | 1.00 |
|  | **Specificity** | 1.00 | 1.00 | 1.00 | 1.00 | 1.00 | 1.00 | N/A | N/A |
|  | **PPV** | N/A | N/A | N/A | N/A | N/A | N/A | N/A | N/A |
|  | **NPV** | 1.00 | 0.99 | 0.99 | 0.99 | 0.99 | 0.99 | N/A | N/A |
| **V** | **Sensitivity** | 0.00 | 0.00 | 0.00 | 0.00 | 0.00 | 0.00 | 1.00 | 1.00 |
|  | **Specificity** | 1.00 | 1.00 | 1.00 | 1.00 | 1.00 | 1.00 | N/A | N/A |
|  | **PPV** | N/A | N/A | N/A | N/A | N/A | N/A | N/A | N/A |
|  | **NPV** | 1.00 | 0.99 | 0.99 | 0.99 | 1.00 | 1.00 | N/A | N/A |
| **Total N (Training group)** | | 3092 | | | | | | | |
| **Total N (Test group)** | | 1326 | | | | | | | |

Table 10. Diagnostic accuracy statistics for Clavien Dindo complication classification
